# One year of SARS-CoV-2: Genomic characterization of COVID-19 outbreak in Qatar

**DOI:** 10.1101/2021.05.19.21257433

**Authors:** Fatiha M. Benslimane, Hebah A. Al Khatib, Ola Al-Jamal, Dana Albatesh, Sonia Boughattas, Ayeda A Ahmed, Meryem Bensaad, Shameem Younuskunju, Yasmin A Mohamoud, Mashael Al Badr, Abdalla A. Mohamed, Reham A. El-Kahlout, Tasneem Al-Hamad, Dina Elgakhlab, Fatima H. Al-Kuwari, Chadi Saad, Andrew Jeremijenko, Abdullatif Al-Khal, Muna A. Al-Maslamani, Roberto Bertollini, Einas A. Al-Kuwari, Hamad E. Al-Romaihi, Salih Al-Marri, Mohammed Al-Thani, Radja M. Badji, Hamdi Mbarek, Yasser Al-Sarraj, Joel A Malek, Said I. Ismail, Laith J. Abu-Raddad, Peter V. Coyle, Asmaa A. Al Thani, Hadi M. Yassine

## Abstract

Qatar, a state that has a diverse population consisting mainly of foreign residents, has experienced a large COVID19 outbreak. In this study, we report on 2634 SARS-CoV-2 whole-genome sequences from infected patients in Qatar between March-2020 and March-2021, representing 1.5% of all positive cases in this period. Despite the restrictions on international travel, the viruses sampled from the populace of Qatar mirrored nearly the entire global population’s genomic diversity with nine predominant viral lineages that were sustained by local transmission chains and the emergence of mutations that are likely to have originated in Qatar. We reported an increased number of mutations and deletions in B.1.1.7 and B.1.351 lineages in a short period. These findings raise the imperative need to continue the ongoing genomic surveillance that has been an integral part of the national response to monitor the SARS-CoV-2 profile and re-emergence in Qatar.

## 1. Introduction

Following its first appearance in China, the ongoing pandemic of Corona Virus Disease 19 (COVID-19) caused by severe acute respiratory syndrome coronavirus 2 (SARS-CoV-2) has urged the global community to take steps to ease its transmission [1, 2]. Qatar, which is characterized by a diverse population of 2.8 million [3, 4], has experienced a large outbreak with over >75,006 infections per million population, ranking as one of the highest countries with laboratory-confirmed rates. The first confirmed COVID-19 case in Qatar was reported on February 28 2020 in quarantined individuals with a history of travel from Iran. A large outbreak of over 300 infections was then identified as the first community cluster on March 6 2020 among manual workers and expatriate craft [5]. The country declared a suspension on entry of foreign nationals on March 17, 2020, allowing only exceptional entry to Qatari citizens and posed strict measures to control the epidemic curve. Despite these measurements, Qatar witnessed a large outbreak, with the highest confirmed cases of 2,355 per day reported on May 30, 2020. By July 2020, cases dropped dramatically to an average of 200 cases per day. As such, the country lifted the travel restriction in August 2020 and started accepting returning residence while implementing a mandatory quarantine. Restrictions inside Qatar were also eased, and the lockdown was elevated in four phases [6]. In December 2020, new variants of concerns (VOC) were detected in other countries, including the UK and South Africa [7-10]. These strains were related to increased transmissibility and posed higher public health concerns. Qatar posed strict restrictions on travelers arriving from those countries to prevent the strains’ entrance into the country. However, soon after their global detection, COVID-19 positive cases increased significantly, indicating that a second wave is hitting the country. As of March 31, 2021, 179,965 patients have tested positive, including 291 fatalities [6], although seroprevalence studies indicated a higher infection rate within the population [3, 11]. Throughout the epidemic in Qatar, we implemented a sequencing strategy to detect, monitor, and evaluate the spread of the virus, including the VOC. Such sequencing efforts were crucial towards understanding the epidemiological and clinical significance of variants, especially when it comes to estimating reinfection rates and vaccine effectiveness [12-16]. Here we report on the clusters of SARS-CoV-2 infection reported in Qatar for the past year, intending to understand the origin, evolution, transmission patterns of the virus. This is the first study of its kind from the MENA region, which will add context to the international genomic consortiums and global data towards combating the pandemic.

## 2. Methods

### 2.1. Ethics statement

The study was approved by the Institutional Review Board (IRB) committees of Qatar University (QU-IRB 1289-EA/20) and Hamad Medical Corporation (MRC-01-20-145) ethical boards. Samples collected were retrospective as such, following the national legislation and the institutional requirements, written informed consent for participants was not required for this study.

### 2.2. Clinical sample collection

Samples selection was based on epidemiological characteristics representing various cluster types and nationalities. The selection was based on investigations that were conducted by public health teams in the country on data that was fed into the national database. Data including patient’s demography (age, gender, nationality) and epidemiology (date of onset and designation) were retrieved. COVID19 positive (CT<35) respiratory specimens (nasopharyngeal swabs) of 2634 patients were retrieved from the virology laboratory at Hamad Medical Corporation (HMC), which is the main lab that provides diagnostic testing in the nation. Samples were handled in a biosafety level 3 laboratory with full personal protective equipment and adapted procedures to airborne pathogens by trained personnel as recommended by the World Health Organization [17]. Samples were selected from main clusters since the start of local transmission for a period of one year, March 10 2020 to March 29, 2021.

### 2.3. Whole-genome sequencing analysis

The RNA was extracted using NucleoSpin RNA Virus isolation kits (Macherey-Nagel) or MGI automated extraction platforms and kits (MGI, China) according to the manufacturer’s protocols. Two sequencing platforms and protocols were used, Oxford Nanopore Technology (ONT) and Illumina Miseq.

The ARTIC Network SARS-CoV-2 sequencing protocol and V3 primer amplicon set were used for sequencing SARS-CoV-2 near full genome on Oxford Nanopore’s GridION platform (https://artic.network/ncov-2019)[18]. In brief, complementary DNA was synthesized from the extracted RNA based on the CT values of the samples using SuperScript IV reverse transcriptase (Thermofisher Scientific). A multiplex PCR was then performed using Q5^®^ Hot Start High-Fidelity Master Mix (NEB) and V3 primer set to generate amplicons that are 400bp in length. Sequencing libraries were prepared using Nanopore’s ligation kits (SQK-LSK109) and native barcode kits (EXP-NBD104 and EXP-NBD114) to multiplex up to 24 samples per sequence run on an R9.4 flow cell (1). DNA sequencing and analysis on the Illumina Miseq platform were carried out as previously described [14, 16]. Briefly, libraries were constructed from viral RNA using the Paragon Genomics CleanPlex SARS-CoV-2 Panel and sequenced on the Illumina MiSeq according to the manufacturer’s recommended protocol.

### 2.4. Bioinformatics analysis

For ONT reads, MinKNOW software was used for base calling and reads with a minimum Q score of 7 were considered for downstream analysis. ARTIC network nCoV-2019 novel coronavirus bioinformatics protocol (https://artic.network/ncov-2019/ncov2019-bioinformatics-sop.html) was used for variant calling and generation of consensus sequences. In brief, guppy_barcoder was used to demultiplex the data. Only fragments that had barcodes at both ends were analyzed. Reads between 400bp and 700bp were filtered and variant calling was performed using Medaka tool in reference to the SARS-CoV-2 genome sequence (accession MN908947). The consensus was called when there is at least 30x coverage. For Illumina reads, sequences were quality/adapter trimmed with CUTADAPT, primer sequences removed with FGBIO, aligned with BWA-MEM and SNPs called with SAMTOOLS as previously described [14, 16]. SARS-CoV-2 lineages were determined based on pangolin (V.2.4.2) nomenclature [19]. For mutation analysis, the cut-off value for considering a mutation for analysis was set to a prevalence of ≥ 1%.

Phylogenetic analysis was performed for 1,452 high-coverage genomes (>90%) with collection dates ranging from March 2020 to March 2021. A Maximum Likelihood (ML) phylogeny was generated using IQTree v2.1.3 under a GTR model of nucleotide substitution with empirical codon frequencies plus the FreeRate model [20]. Independent ML phylogenies were generated for the B.1.1.7 (n=134) and B.1.351 (n=112) genomes. Global SARS-CoV-2 genomes representative of the two lineages were retrieved from GISAID (https://www.gisaid.org/) and aligned with our sequences. Global sequences were selected to include four genomes per continent per week (based on sample collection day), from December 2020 to March 2021. Models for phylogenetic tree configurations were selected based on IQ-Tree’s ModelFinder tool [21]. Phylogenetic tree of the B.1.1.7 genomes was generated under the TIM model with empirical base frequencies and invariant sites (TIM+F+I). Phylogenetic tree of the B.1.351 genomes was generated under the GTR model with empirical base frequencies, invariant sites, and the discrete Gamma model (GTR+F+G4). Phylogenetic trees were edited and visualized using Figtree software v1.4.4 (http://tree.bio.ed.ac.uk/software/figtree/). root-to-tip regression analyses of B.1.1.7 and B.1.351 genomes were performed using TempEst v1.5.3 to investigate the temporal signal of the datasets [22].

## 3. Results

### 3.1. Epidemiology of SARS-CoV-2 infection in Qatar

The population in Qatar is divided between manual and craft workers (∼60%) and the urban population. While during the first wave (April to June 2020) the virus transmission was mostly sustained in the first larger population that reached ∼65%–70% seroprevalence by September 2020, the transmission was more confound in the urban population during the second wave (started early March 2021). As such, around 70% of the samples selected for this study were from the large cluster, while 30% were from the urban cluster. An average of 3.12±8.18% of daily COVID19 positive samples were sequenced (Supplementary Figure.1.A), representing 80.26% of the days during the studied period (309 out of 385 days). The tested population was diverse as it included patients from 63 countries, with the majority being from Asia (85.81%, Supplementary Figure.1.B). The median age was 35 years and 74.86% were male (Table.1).

**Table.1.** Demographic data.

| Category | Number of cases | Frequency |
| --- | --- | --- |
| **Gender** |  |  |
| Male | 663 | 25.17% |
| Female | 1971 | 74.83% |
| **Age** |  |  |
| Infant, 0-1 yrs. | 15 | 0.57% |
| Toddler, 2-4 yrs. | 32 | 1.21% |
| Child, 5-12 yrs. | 101 | 3.83% |
| Teen, 13-19 yrs. | 107 | 4.06% |
| Adult, 20-39 yrs. | 1422 | 53.99% |
| Middle Age Adult, 40-59 yrs. | 780 | 29.61% |
| Senior Adult, 60+ | 177 | 6.72% |
| **Population** |  |  |
| Continent | Number of Countries | Frequency |
| Asia | 26 | 85.81% |
| Africa | 19 | 55.85% |
| Europe | 12 | 5.22% |
| America | 6 | 5.50% |
| Australia | 1 | 0.40% |

### 3.2. Whole-genome analysis of SARS‐CoV‐2 samples

Out of 2634 samples, 2241 (85.1%) yielded a coverage above 60% and were used for downstream analysis. Sequence coverage was significantly inversely correlated with the CT value (R^2^=0.0436 and p-value < 0.0001). All generated SARS-CoV-2 genomes in this study were deposited to the Global Initiative on Sharing All Influenza Data (GISAID). Accession IDs are listed in Supplementary Data 2.

The consensus sequences were analyzed using Pangolin COVID-19 Lineage Assigner tool. Data was then manually checked to exclude any sequences that were missing collection date, had no amino acid mutations, or their lineage assignment was not accurate in reference to the detected sequence mutations. This resulted in a total of 2013 sequences that were included in variant analysis. Nine lineages were reported in at least 1% of the samples (Supplementary Figure.2.A) with the majority falling under lineage B.1.428 (32.84%; including B.1428.1, and B.1428.2, B.1428.3), followed by B.1 (20.91%), B.1.1.7 (8.74%), B (6.61%), B.1.351 (6.06%), B.1.1 (5.17%), B.40 (3.68%), A (3.48%), and B.1.1.75 (2.14%). These lineages were circulating across both genders as well as different ages and were detected among imported, quarantined and local cases (Figure.1.A). The nine lineage were considered as highly prevalent. A further 25 lineages were detected but in less than 1% of the samples (at least two sequences). These include B.1.1.10, B.1.1.161, B.1.1.163, B.1.1.216, B.1.1.28, B.1.1.354, B.1.153, B.1.159, B.1.160.20, B.1.165, B.1.170, B.1.177, B.1.2, B.1.36, B.1.36.10, B.1.36.29, B.1.411, B.1.427, B.1.446, B.1.466.1, B.1.577, B.1.610, B.1.612, B.28, and B.36 (Supplementary Figure.2.B). Another 24 lineages were detected only once, including A.23.1, A.28, AE.4, B.1.1.171, B.1.1.243, B.1.1.263, B.1.1.53, B.1.12, B.1.128, B.1.147, B.1.160, B.1.280, B.1.289, B.1.36.17, B.1.389, B.1.441, B.1.518, B.1.533, B.1.545, B.1.565, B.1.9.5, B.23, B.4, and C.36.

**Figure 1:**
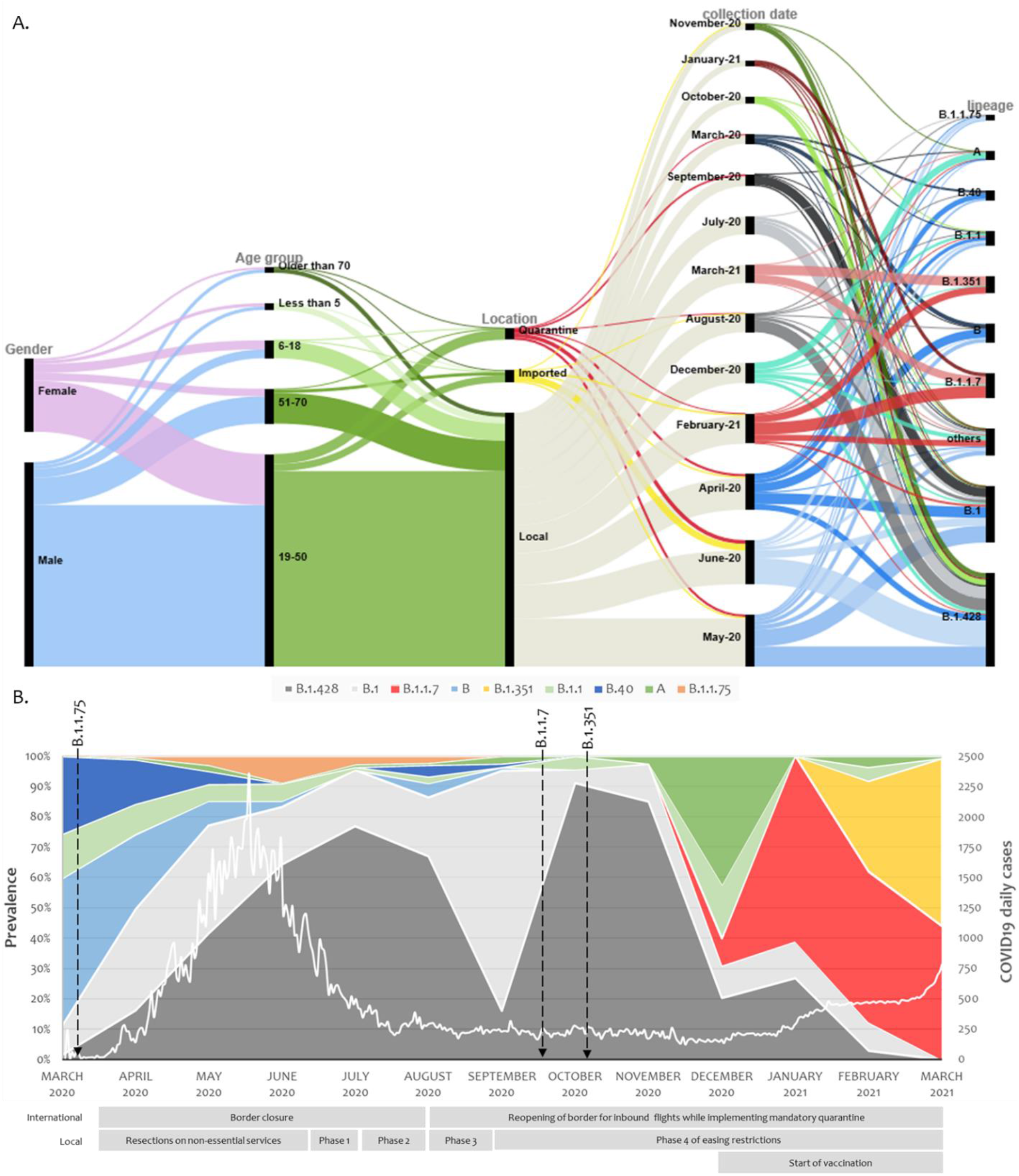
Demographics and virological characteristics of Qatar study cohort. **(A)** SARS-CoV-2 major lineages distribution over the one-year study period from March 2020 to March 2021. Arrows represent when the designated lineages were reported worldwide. The white line demonstrates the daily COVID19 positive cases in Qatar. The international and national restrictions implemented by the country is presented below the stacked area chart. **(B)** Demographics, swab collection dates and virological parameters of the study cohort are demonstrated in an alluvial diagram with the display of relatedness among features of each two neighboring nodes.

Out of the nine highly prevalent lineages, A, B, B.1, B.1.1, B.1.1.75, B.1.428 and B.40 were detected in Qatar from March 2020 and their prevalence was further enlarged with local transmission. B.1.428 lineage (including sublineages B.1.428.1, B.1.428.2 and B.1.428.3 which were first detected on 27-Apr-2020, 9-Jun-2020, and 3-Jun-2020, respectively) was of the main contributors toward the country’s first pandemic wave. As the country re-opened its boundaries and eased the local restrictions, B.1 prevalence increased in September, but the B.1.428 quickly took over again. In December, when traveling restrictions were further reduced, the prevalence of lineage A increased for a short interval, and two new lineages were imported into Qatar: B.1.1.7 on 22-Dec-20 and B.1.351 on 29-Dec-20. The imported lineage quickly succeeded in eliminating all other lineages, with B.1.1.7 and B.1.351 reaching 43.75% and 54.69%, respectively, by the end of March 2021 (Figure.1.B). The Effective Reproduction Number (Rt) was above one at the beginning of the pandemic. It decreased below one during summer and remained so until the introduction of B.1.1.7 and B.1.351 lineages in December 2020, where it reached 1.5 by the end of March 2021 (Supplementary Figure.3.A).

### 3.3. SARS-CoV-2 variant analysis

Non-synonymous mutation analysis was performed after normalizing the frequency of the variables in reference to the coverage in each region. In this study sample, 1583 mutations were detected across different gene regions; however, only the 85 mutations that had a prevalence of 1% were reported. The most prevalent mutation was D614G (84.20%) found in the Spike glycoprotein followed by P314L (83.12%) in the open reading frame 1b (ORF1b). The prevalence of these two mutations increased with the course of the pandemic, and they have been detected in all sequences from May 2020 onwards. Three other mutations were detected in over 50% of the sequences: ORF3a:Q57H (57.50%), N:T205I (53.85%), ORF1a:T265I (53.43%). They were dominant from June 2020 until November/December 2020, where their prevalence started to decrease. Twenty-one mutations and six deletions were detected in the spike glycoprotein, four of which were in the receptor binding domain (RBD); S:K417N (6.14%), S:N481K (8.63%), S:E484K (2.79%), and S:N501Y (12.14%). S:N481K was detected early in the pandemic, while S:K417N, S:E484K, and S:N501Y were detected later on in December 2020 (Figure.2).

**Figure.2.**
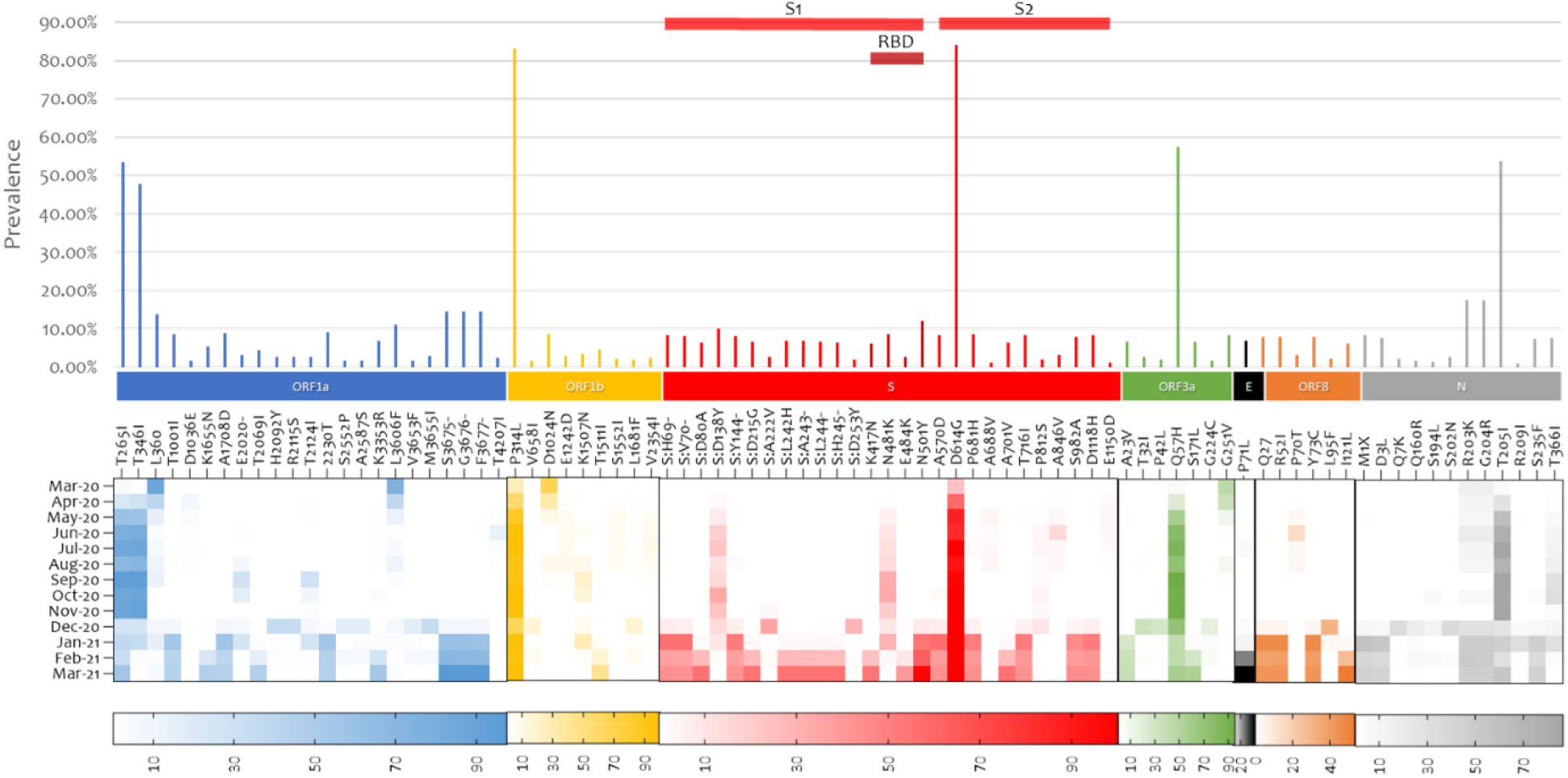
whole genome SARS-COV-2 mutation analysis. Mutations with at least 1% frequency are presented in bars and their evolution during the studied one-year period is presented in the heat map. SARS-COV-2 gene annotation is shown in colors. Abbreviation; ORF: Open reading frame, S: Spike, E: Envelope, N: Nucleocapsid.

In-depth non-synonymous mutation analysis was performed independently for each of the major variants that were highly prevalent. Mutations with a frequency of ≥1%, calculated from the total sequences detected in each variant, were considered with focus on mutations that were reported in at least 1% of the studied cohort. B.1.1.7 had the highest number of mutations (61) followed by B.1.351 (49) and B.1.428 (47) (Supplementary Figure.3.B). Some mutations were constant in the lineages while others appeared later on as the pandemic progressed. Focusing on the RBD in the spike glycoprotein, S:N481K was detected in the backbone of B.1.428 and B.1, S:K417N and S:E484K were in the backbone of B.1.1.7 and B.1.351 while S:N501Y was detected highly in B.1.1.7, B.1.351 and at a lower frequency in B.1 (Figure.3).

**Figure.3.**
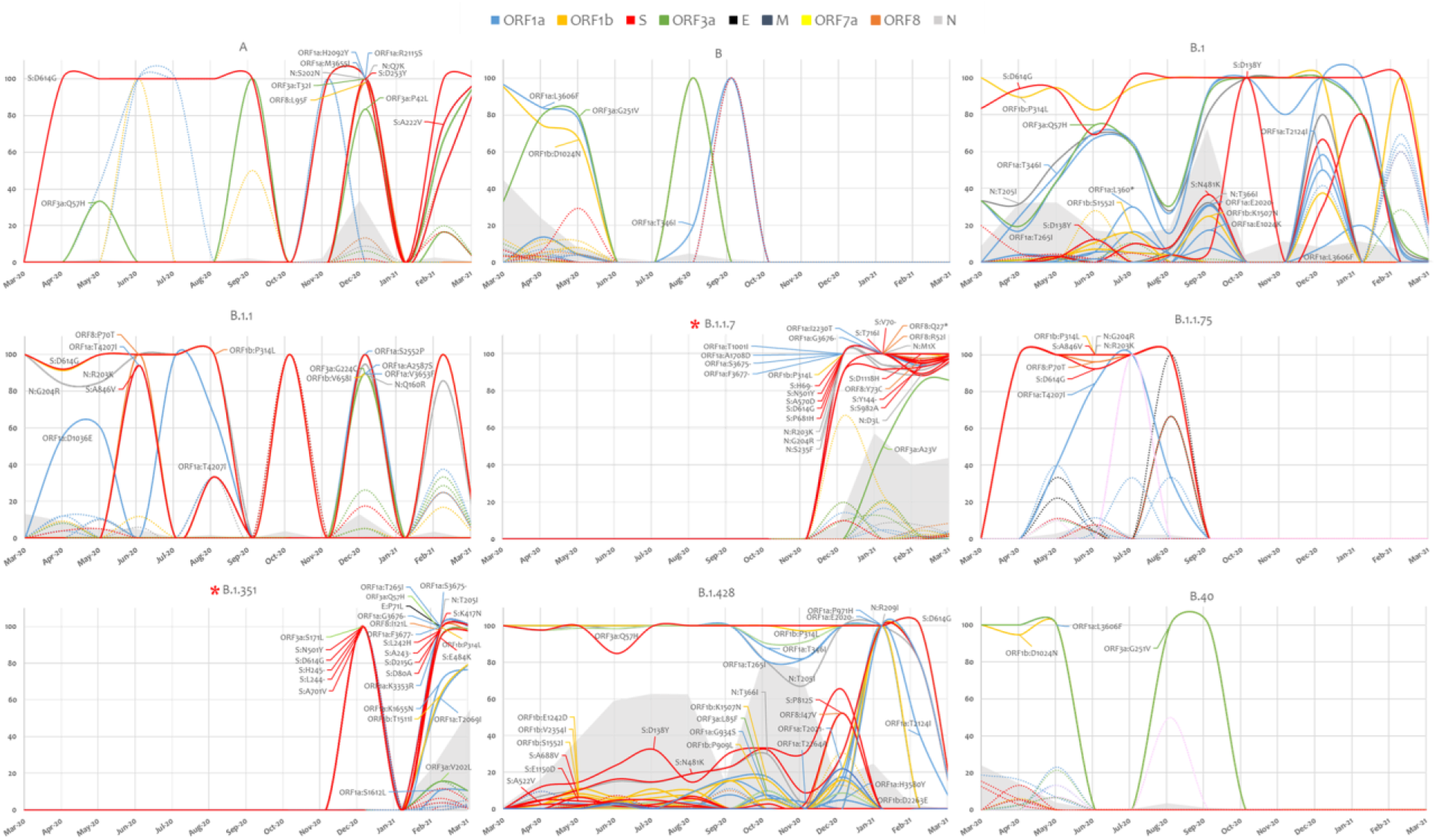
SARS-COV-2 lineage specific mutation analysis. Mutation in highly prevalent lineages are presented based on their prevalence. In solid lines, mutations that were detected at a frequency of at least 1% from the study cohort. Mutation in dotted lines did not pass the 1% cutoff in the study cohort but were present in at least 1% of the sequences in a particular lineage. The grey area represents the evolution of each lineage across the studied period. Variant of concern are identified with asterisk.

### 3.4. Phylogenetic analysis

Phylogenetic analysis of high-coverage genomes sequenced during the one-year study period revealed distinct clustering patterns (Figure.4). Based on pangolin classification, genomes clustered into nine major clusters and more than ten minor lineages. Phylogenetic analysis of the B.1.1.7 and B.1.351 genomes demonstrated unique clustering patterns regardless of the collection date. The majority of B.1.1.7 genomes (80%) clustered together in five highly supported clades (posterior probability = 1) irrespective of the sample collection date (Figure.5.A). The rest of B.1.1.7 genomes clustered with genomes from different countries, suggesting multiple introductions of B.1.1.7 in Qatar. Generated data suggested introductions from Europe, particularly from the UK, from February to March 2021. It also suggested possible introductions from the USA during February 2021, and from Hong Kong and India during March 2021. Similarly, phylogenetic analysis of the B.1.351 genomes demonstrated the clustering of local genomes in distinct clades (12 clades) (posterior probability = 1) (Figure.5.B). Notably, the B.1.351 genome of the first detected B.1.351 case in Qatar (December 2020) was in the outgroup of a highly supported clade (posterior probability = 1), clustering with B.1.351 genomes from the USA. The majority of B.1.351 genomes collected during February to March 2021 clustered in 10 clades. Only a few samples -collected in March 2021-intermixed with genomes from Greece, South Africa, and the USA (Figure.5B).

**Figure.4.**
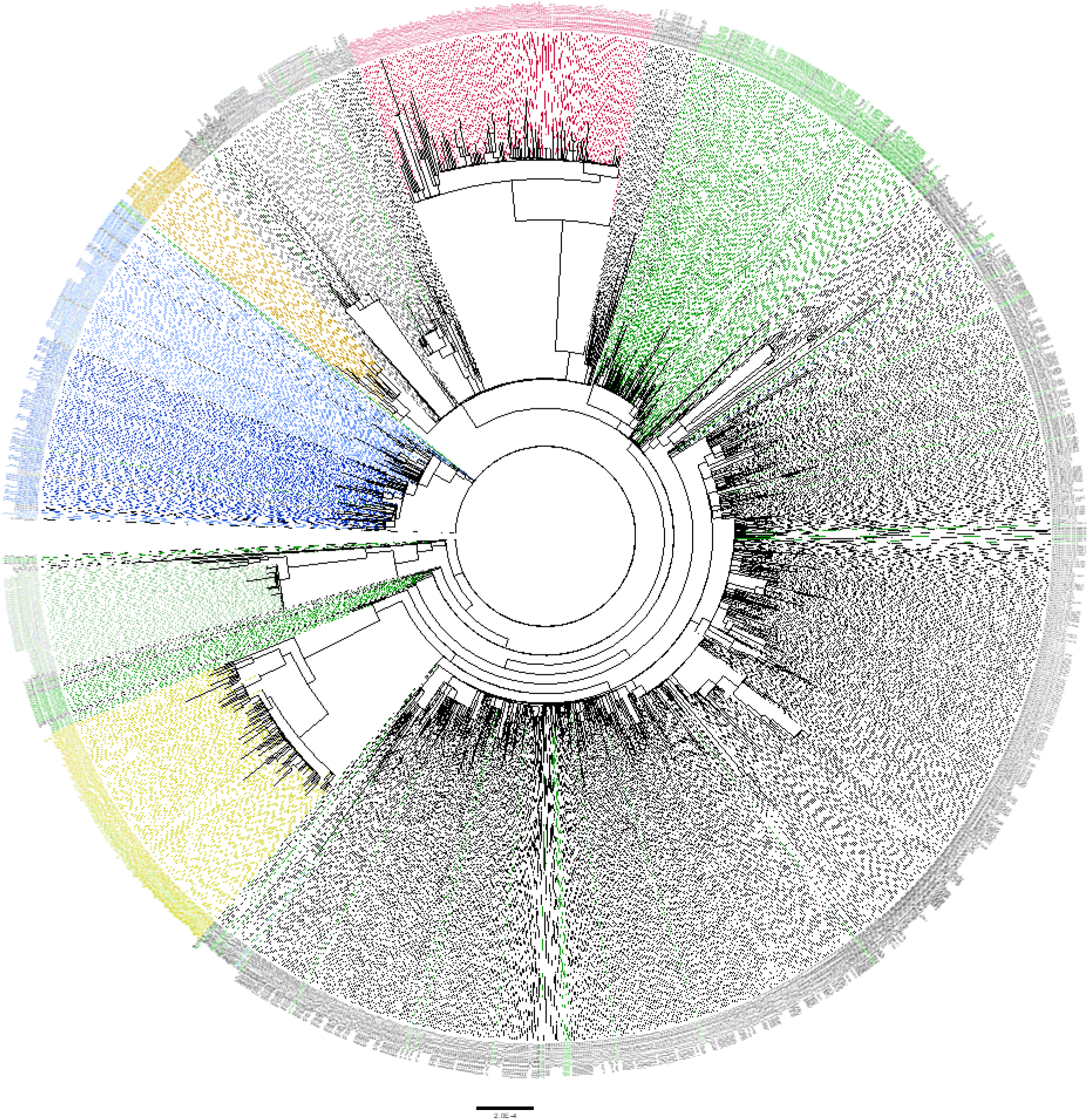
Maximum likelihood tree of high-coverage SARS-CoV-2 genomes included in this study. Phylogenetic tree was generated using 1,452 high-coverage genomes (>90%) sequenced during the period from March 2020 to March 2021. SARS-CoV-2 lineages are indicated in colors as follows: A in light green, B in light blue, B.1 in dark green, B.1.1 in grey, B.40 in in dark blue, B.1.1.75 in light brown, B.1.1.7 in bright red and B.1.351 in yellow. All other lineages are colored in black.

**Figure.5.**
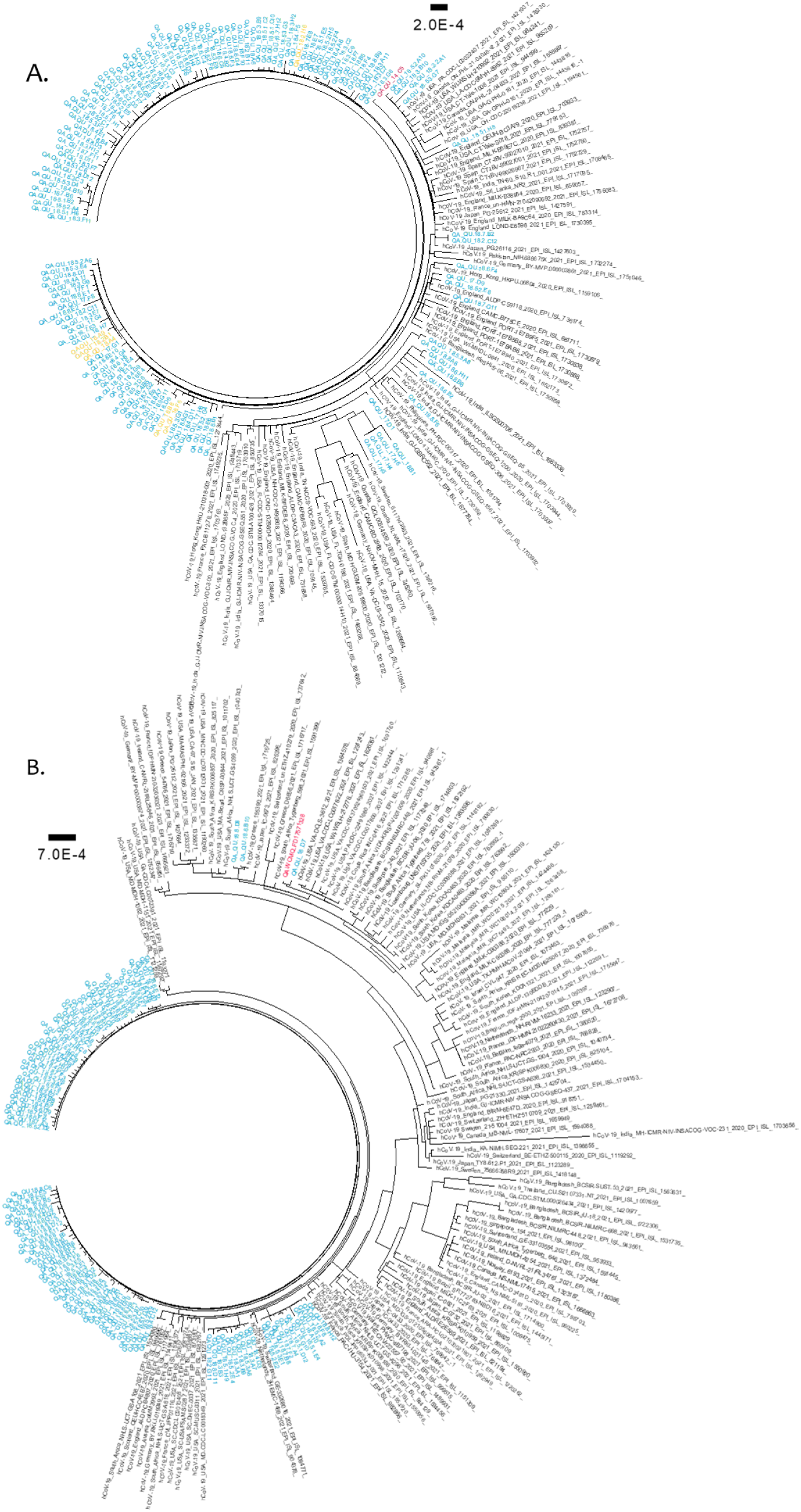
Maximum likelihood trees of SARS-CoV-2 variants of concern detected in this cohort. Phylogenetic tree of B.1.1.7 genomes **(A)** and B.1.351 genomes **(B)** were generated using 134 and 112 local sequences, respectively. Global genomes representative of the two clades with collection dates from November 2020 to March 2021 were included in the phylogenetic tress. Global genomes are indicated in black, local genomes are colored in blue, and imported samples are colored in yellow. The first genome detected locally is colored in red.

## 4. Discussion

The World Health Organization declared the COVID-19 outbreak in China a public health emergency of international concern on January 30 2020, then a worldwide pandemic on 11 March 2020 [17, 23, 24]. By that time, the virus had spread to more than 40 countries and resulted in more than 140,000 infections and more than 4000 deaths worldwide [25]. By the first week of March, the virus was reported in many countries, including eight in Middle East: Iran, Pakistan, Afghanistan, Kuwait, Bahrain, Iraq, Oman, and Qatar [26]. The first covid-19 case in Qatar was reported on February 29, from a traveler returning from Iran that was experiencing the worst outbreak in that region of the world [27]. However, the first documented case of SARS-CoV-2 community transmission was identified on March 6 among expatriate craftworkers living in high-density accommodations [3, 5]. The number of cases increased dramatically since then, reaching 781 confirmed cases by the end of March 2020 [28]. The number of cases continued to increase until it reached the peak of the first pandemic wave in mid-May 2020 [29]. Qatar reached the first pandemic peak earlier than neighboring countries such as Saudi Arabia, Iraq, Lebanon, and Jordan, but late compared to Asian and European countries [29]. The positivity rate started to decrease after May 22, reaching 17% on July 9 [3]. As of July 15, a total number of 104,984 cases had been confirmed in Qatar, a rate of 36,439 cases per million population, one of the highest rates worldwide [3]. The first wave (March-July 2020) was characterized by the circulation of seven main SARS-CoV-2 lineages; out of more than 80 lineages identified worldwide [30, 31]. Locally, the most prevalent lineages were B.1.428 (32%), B.1 (21%), and B.1.1 (5%). Worldwide, the B.1 lineage was dominating, accounting for 17% of COVID-19 cases followed by the B.1.1 (13%) [19, 30, 32]. The B.1.428, on the other hand, was less prevalent globally, accounting for less than 1% of the positive cases [30]. In August, the number of local cases declined dramatically, and accordingly, restrictions on socialization and travel were loosened [29, 33]. Despite the strict quarantine policy on traveling, more lineages were still detected locally but had limited spread. Globally, the overall situation of the pandemic seemed to be under control during September and October except for India, Brazil, and the USA [25]. During this period, B.1.1 and B.1 were the dominating lineages globally, accounting for 12% and 6% of cases, respectively. Locally, the B.1 was dominating, accounting for 48% of cases, followed by B.1.428 (32%). Both lineages were responsible for more than 80% of cases in Qatar until November 2020. B.1.428 lineage, also called the Danish lineage, emerged first in Denmark on March 3 2020 and was later on detected in Australia in, the United States in, Sweden, and Qatar [32]. Qatar reported the highest number of B.1.428 cases (62%) worldwide. Of note, the B.1.428 sub-lineage, B.1.428.1, was identified for the first time in Qatar on April 27. B.1.428.2 and B.1.428.3 sub-lineages were also detected in Qatar shortly after their emergence in neighboring countries. The highest prevalence of B.1.428 sub-lineages were reported in MENA region including Iraq, UAE, Jordan, and Tunisia [32]. They were less commonly reported in Germany, UK, and France [32]. In addition to the B.1.428 lineage, Qatar reported the highest worldwide prevalence (42%) of the Belgian lineage, B.1.1.75, which was identified for the first time in Belgium on March 17 2020 and appeared shortly in Qatar on April 5.

Despite the modest decrease in COVID-19 cases during September-November, early signs of a second wave emerged in many countries by the end of November 2020 [30]. By mid-December, European countries were already facing a second wave of SARS-CoV-2 transmission [25]. The rise in infection rates was attributed to the relaxation of lockdown measures as well as the emergence of novel SARS-CoV-2 variants [34-37]. Several genetic variants emerged in the UK, South Africa, and Brazil [38-40]. The CDC classified five of the newly identified variants, based on their impact on transmission and neutralization, as VOC. Those include B.1.1.7 (UK), B.1.351 (South Africa), P.1 (Brazil), B.1.427, and B.1.429 [41]. Three of these variants have been already detected in Qatar. The B.1.427 variant was the first to be detected on May 3. It is characterized by the spike mutation L452R, has been reported to increase transmissibility by ∼20% as well as the reduction in neutralization by convalescent and post-vaccination sera. Despite the published reports about its increased transmissibility by 20%, only sporadic cases (12 cases) were reported in Qatar from May 2020 to March 2021 [42]. Similarly, only sporadic B.1.427 cases were reported in the other 32 countries that reported this variant [30, 32]. The UK variant, B.1.1.7, was the second VOC to be detected in Qatar. The first ten cases were identified in late December from local cases. At that time, the variant was circulating in Europe and at lower frequency in the USA, Africa, and other Middle East countries [30]. As the number of B.1.1.7 cases was increasing locally and globally, another fast-spreading variant has been identified in South Africa [39]. By December 31, the South African variant, B.1.351, was detected in at least 60 countries, including Qatar [30]. However, the first community transmission case in Qatar was detected on February 1 and soon later, it took hold. By March 15, B.1.351 cases have reached 60% locally and 7% globally [30]. The prevalence of B.1.17 was evident by the S-gene target failure, using Thermo Fisher Scientific’s, USA, TaqPath COVID-19 Combo Kit platform [45], and for B.1.351, the sequencing data presented here [15]. Qatar ranked among the top 20 countries reporting B.1.351 cases, and the highest in the MENA region, despite the late appearance of the variant compared to other countries [30, 32]. The surge in B.1.351 and B.1.1.7 cases was maximized during March 2021 and was associated with a sharp increase (doubled in less than a week) in daily cases, consecutively, the Rt value increased to 1.5. Although the daily cases are still relatively lower than the first wave, the death rate, however, is significantly higher (Average death from March 1 to 20 2021 0.50±.57 as compared to 1.73±1.01 from March 21 to 31 2021, P<0.0001). This triggered the government to reimpose strict lockdown measures to control their spread in the community [29, 33]. The CDC classified these two variants as VOC after showing evidence of increased transmissibility and reduced neutralization by convalescent sera [41]. Both variants were found to increase the transmissibility by at least 50% [43, 44]. The fast-spreading variants, however, exhibited differential resistance to neutralization by convalescent sera. According to some studies, while neutralization against the U.K. variant dropped by roughly 2-fold, it dropped by 6.5- to 8.5-fold against the South Africa variant, using convalescent and post-vaccination sera [46-48]. This raised global concerns about the effectiveness of available vaccines against the two novel variants. In a recent study from Qatar, the effectiveness of the BNT162b2 vaccine was estimated at 89.5% for the B.1.1.7 variant and 75.0% for the B.1.351 variant but exhibited 100% effectiveness toward developing sever disease [15].

The vaccine effectiveness can be rendered by the accumulation of immune-escape mutation(s) in the spike protein, specifically those in the RBD. As a typical RNA virus, mutations frequently arise in the SARS-CoV-2 genome due to the error-prone replication process [49]. However, the majority of these mutations are lost as a result of the natural selection process [50]. Only mutations that confer fitness effect (increased replication, transmissibility, immune evasion) may ultimately fix in the global population of SARS-CoV-2. Here, we analyzed the mutations that emerged in each lineage over a one-year study period. Overall, sequence analysis revealed more than 99% similarity among genomes that belong to the same lineage. Fewer similarity scores were reported for B.1.351 (98%) and A (97%) genomes. Despite the relatively short circulation period of B.1.1.7 and B.1.351, additional mutations were accumulated in their genome. The number and prevalence of these mutations were limited in B.1.1.7 compared to B.1.351. Besides the lineage-defining mutations of B.1.1.7, we identified five additional high-prevalent non-S mutations; ORF1b:P314L (98.3%), N:M1X (99.4%), N:G204R (96.6%), N:R203K (96.6%), and ORF3a:A23V (74.4%), all of which are reported at high prevalence among global B.1.1.7 genomes [51]. B.1.351 genomes, on the other hand, exhibited a higher number of mutations compared to the B.1.351 representative genome. In addition to mutations that characterize B.1.351 lineage, we identified nine high-prevalent mutations; ORF1a:T265I (100.0%), ORF1a:T2069I (73.6%), ORF1a:K3353R (98.3%), ORF1b:P314L (100.0%), ORF1b:T1511I (73.9%) ORF3a:Q57H (99.2%), ORF3a:S171L (96.7%), ORF8:I121L (95.9%), and S:L242H (98.3%). Most of these mutations are reported at medium to high prevalence in global sequences [51]. Interestingly, ORF1b: P314L and ORF3a: Q57H mutations were reported locally in 70% and 98% of B.1.351 genomes, respectively, but in less than 2% of B.1.351 genomes globally [51]. Moreover, 97% of B.1.351 genomes carried three deletions in ORF1a: S3675, G3676, and F3677. On the other hand, these deletions were reported in less than 2% of global B.1.351 sequences [51] and in countries like Peru [52]. Of note, though, the three deletions are reported in more than 98% of B.1.1.7 sequences worldwide [51]. It is, therefore, striking to see these deletions at this high prevalence among B.1.351 genomes in Qatar. Fortunately, none of these new high-prevalent mutations –in B.1.1.7 and B.1.351 is located in the spike gene, the main target of vaccine-induced antibodies [47, 53] but it might influence replication and hence transmission. Future studies are needed in this regard. Only sporadic mutations were detected in the spike genes of all other lineages. Analysis of the spike mutations in other lineages revealed comparable frequencies to global data with few exceptions. The N481K, for example, was detected exclusively in Qatar and appeared in 17% and 8% of B.1.428 and B.1 genomes, respectively. The D138Y was detected locally in 10% of SARS-CoV-2 genomes as follows: 22% in B.1.428, 5% in B.1, and 2% in B.1.1. On the other hand, global records showed that this mutation is particularly found in B.1.1.7 (38%) and P.1 (38%) genomes and at lower frequency in B.1 (2%) [51]. Experimental analyses are required to understand the molecular and pathophysiological impact of these variations. Further analyses are also needed to predict the impact of accumulated mutations on viral infectivity, immune response, or disease severity. These data are essential to evaluate the virus population structure and infection dynamics.

In conclusion, this study identified SARS-CoV-2 lineages and their circulation pattern in Qatar, a highly diverse region that was heavily impacted by COVID-19. Regardless of the implementation of restriction measure, particularly on international movements, 61 SARS-COV-2 lineages were detected. Nine were widely represented including two VOCs. We identified a number of novel mutations that are likely to have originated in Qatar including N481K located in the spike protein. We also reported an increased number in the mutations and deletions in the backbone of the VOCs, B.1.1.7 and B.1.351, in a short period. This raises the imperative need to continue the ongoing genomic surveillance that has been an integral part of the national response to monitor any SARS-CoV-2 re-emergence in Qatar.

## Data Availability

All generated SARS-CoV-2 genome in this study were deposited to the Global Initiative on Sharing All Influenza Data (GISAID). Accession IDs are listed in Supplementary Data 2.

https://www.gisaid.org/

## 6. Authors contribution

F.M.B, H.A.A, J.A.M, L.J.A and H.M.Y conceptualized the idea. F.M.B and H.A.A performed the analysis that generated the graphs and drafted the manuscript. F.M.B, O.A, D.A, and S.B conducted the ONT wet lab work. H.A.A, A.A.A, M.B, S.Y, and Y.A.M conducted the Illumina wet lab work. M.B and A.A.M performed the RNA extraction using MGI platform. F.M.B performed the initial ONT reads analysis. H.A.A, F.A.A and C.S performed the initial Illumina reads analysis. F.M.B, H.A.A, O.J, R.A.E, T.A, and D.E participated in sample collection. A.J, A.K, M.A.A, R.B, E.A.A, H.E.A, S.A, M.A, L.J.A, P.V.C, A.A.A, and H.M.Y identified the samples for sequencing and extracted demographic information from databases. S.I.I, RB, HM, and YS, supported the idea by securing funds. H.M.Y reviewed the manuscript draft. All authors read the final manuscript draft.

## 8. Acknowledgement

We acknowledge the many dedicated individuals at Hamad Medical Corporation, the Ministry of Public Health, the Primary Health Care Corporation, and the Qatar Biobank for their diligent efforts and contributions to make this study possible.

## 9. Data availability

Genomic data generated in this study is available on GISAID. A list of GISAID genome accession numbers for these data and global genomes used here are provided in Supplementary Data 2.

## 11. Additional Information

Supplementary Information is available for this paper.

