## Supplementary Data 1 for "One year of SARS-CoV-2: Genomic characterization of COVID-19 outbreak in Qatar"

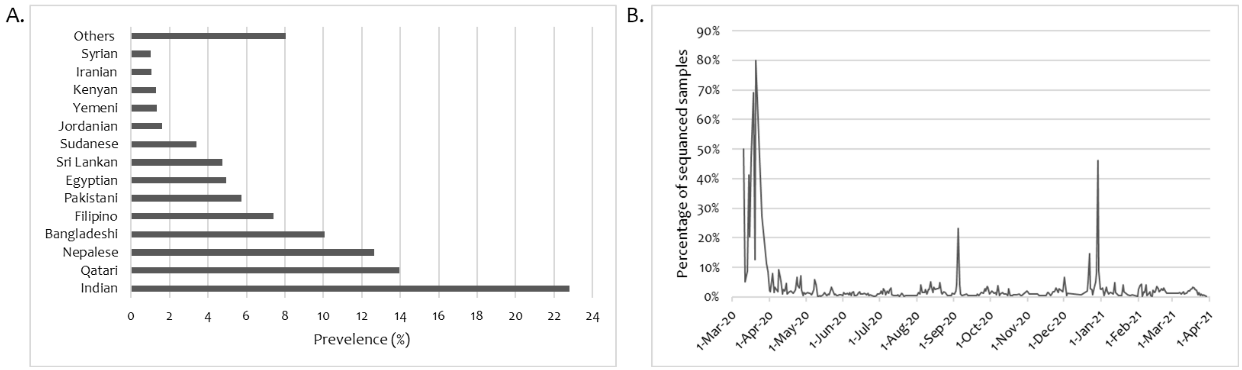


**Supplementary Figure.1. Nationalities and the frequency of sequenced samples in the studies cohort. (A)** Nationalities observed at least at 1% prevalence. Others (<1%) includes: Algerian, American, Argentinean, Australian, Bahraini, Brazilian, British, Cameroonian, Canadian, Chinese, Cuban, Cypriot, Djiboutian, Dominican, Emirati, Eritrean, Ethiopian, French, Gambian, German, Ghanaian, Indonesian, Irish, Italian, Lebanese, Malaysian, Malian, Moroccan, Myanmar, Nepalese, Nigerian, Omani, Palestinian, Portuguese, Romanian, Russian, Saudi, Serbian, Somali, South African, South Korean, Spanish, Sudanese, Swiss, Tajik, Thai, Tunisian, Turkish, Ugandan, and Ukrainian. **(B)** Percentage of sequenced samples in reference to the daily COVID19 positive cases in Qatar.

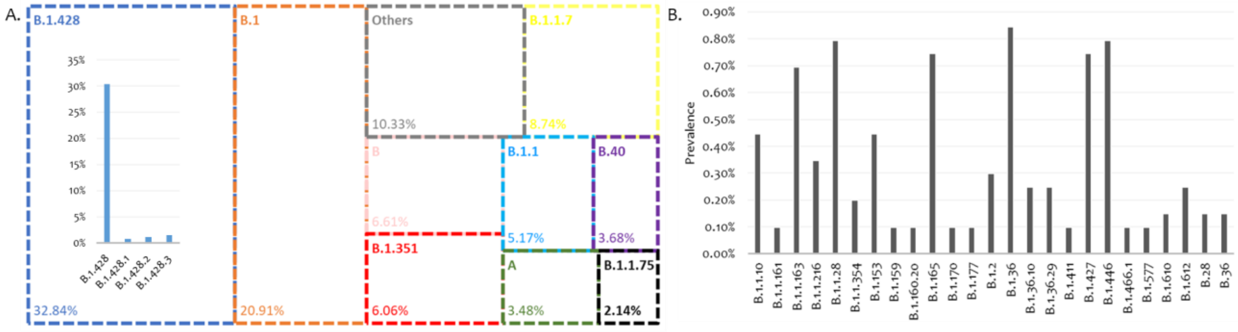


**Supplementary Figure.2. Lineages detected in Qatar's sequenced cohort. (A)** Highly prevalent lineages (>1%) and (B) Low prevalent lineages (<1% and in at least two sequences). Nomenclature is based on Pangolin tool.

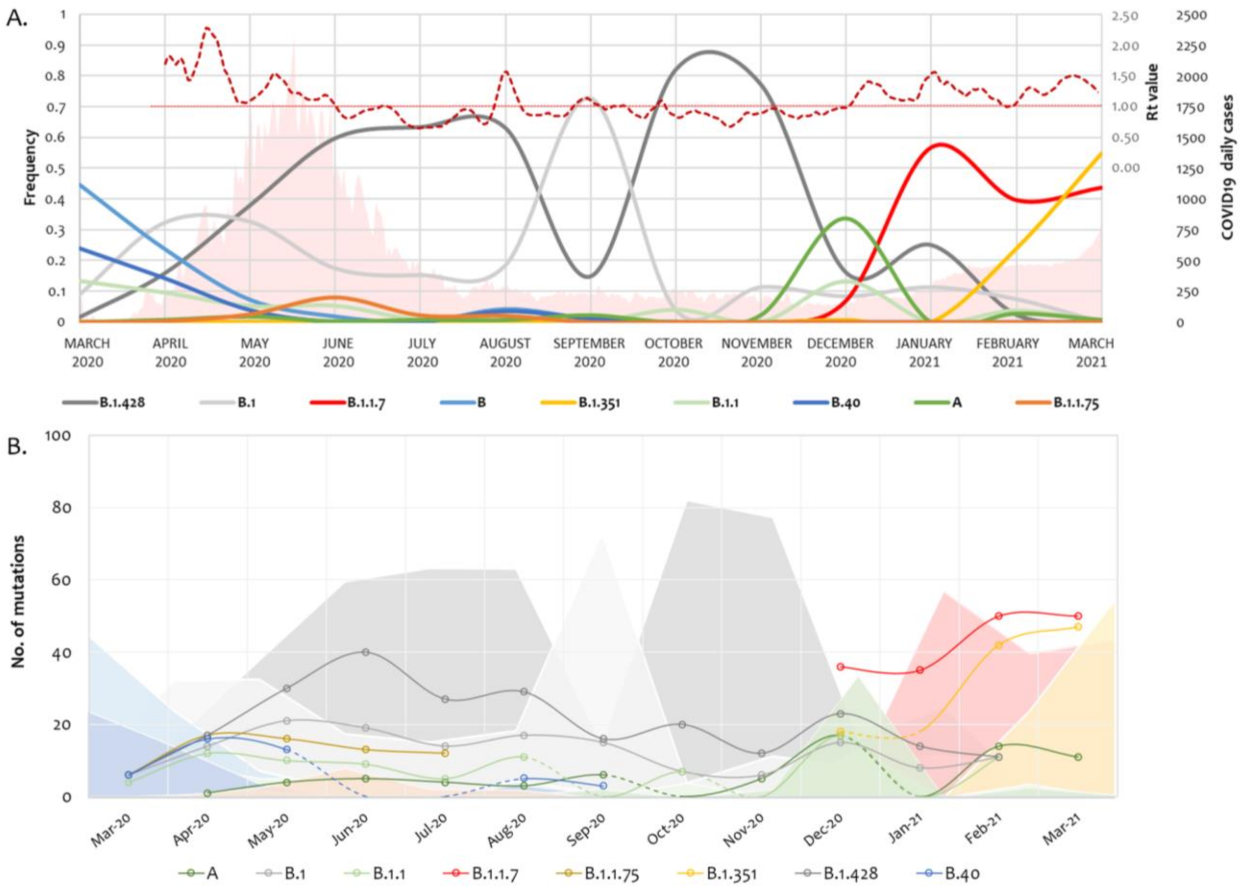


**Supplementary Figure.3. SARS-COV-2 lineages evolution over the one-year study period (March 2020-March 2021). (A)** Frequency of the highly prevalent lineages across the one year period in reference to COVID19 daily positive cases (area in light pink) and the Effective Reproduction Number (Rt, in dotted deep red line). The light red line highlights an Rt value of 1. **(B)** Accumulation of non-synonymous mutations in the genomes of major SARS-CoV-2 lineages.
