## Supplementary Data 2 for "One year of SARS-CoV-2: Genomic characterization of COVID-19 outbreak in Qatar"

| Virus name | Accession ID |
|---|---|
| hCoV-19/Qatar/QA-QU_01-11/2020 | EPI_ISL_1712659 |
| hCoV-19/Qatar/QA-QU_01-12/2020 | EPI_ISL_1712660 |
| hCoV-19/Qatar/QA-QU_01-17/2020 | EPI_ISL_1712662 |
| hCoV-19/Qatar/QA-QU_01-21/2020 | EPI_ISL_1712665 |
| hCoV-19/Qatar/QA-QU_01-23/2020 | EPI_ISL_1712667 |
| hCoV-19/Qatar/QA-QU_01-24/2020 | EPI_ISL_1712668 |
| hCoV-19/Qatar/QA-QU_01-5/2020 | EPI_ISL_1712669 |
| hCoV-19/Qatar/QA-QU_01-19/2020 | EPI_ISL_1712663 |
| hCoV-19/Qatar/QA-QU_02-2-44/2020 | EPI_ISL_1712699 |
| hCoV-19/Qatar/QA-QU_02-2-60/2020 | EPI_ISL_1712705 |
| hCoV-19/Qatar/QA-QU_02-3-36/2020 | EPI_ISL_1712717 |
| hCoV-19/Qatar/QA-QU_01-13/2020 | EPI_ISL_1712661 |
| hCoV-19/Qatar/QA-QU_01-2/2020 | EPI_ISL_1712664 |
| hCoV-19/Qatar/QA-QU_02-2-7/2020 | EPI_ISL_1712706 |
| hCoV-19/Qatar/QA-QU_02-3-14/2020 | EPI_ISL_1712710 |
| hCoV-19/Qatar/QA-QU_02-3-43/2020 | EPI_ISL_1712720 |
| hCoV-19/Qatar/QA-QU_01-22/2020 | EPI_ISL_1712666 |
| hCoV-19/Qatar/QA-QU_02-2-18/2020 | EPI_ISL_1712686 |
| hCoV-19/Qatar/QA-QU_02-2-22/2020 | EPI_ISL_1712689 |
| hCoV-19/Qatar/QA-QU_02-2-27/2020 | EPI_ISL_1712691 |
| hCoV-19/Qatar/QA-QU_02-2-35/2020 | EPI_ISL_1712695 |
| hCoV-19/Qatar/QA-QU_02-2-5/2020 | EPI_ISL_1712702 |
| hCoV-19/Qatar/QA-QU_02-2-59/2020 | EPI_ISL_1712703 |
| hCoV-19/Qatar/QA-QU_02-3-16/2020 | EPI_ISL_1712712 |
| hCoV-19/Qatar/QA-QU_02-3-33/2020 | EPI_ISL_1712715 |
| hCoV-19/Qatar/QA-QU_02-3-37/2020 | EPI_ISL_1712718 |
| hCoV-19/Qatar/QA-QU_02-2-29/2020 | EPI_ISL_1712692 |
| hCoV-19/Qatar/QA-QU_02-2-39/2020 | EPI_ISL_1712696 |
| hCoV-19/Qatar/QA-QU_02-2-4/2020 | EPI_ISL_1712697 |
| hCoV-19/Qatar/QA-QU_02-2-49/2020 | EPI_ISL_1712701 |
| hCoV-19/Qatar/QA-QU_02-2-6/2020 | EPI_ISL_1712704 |
| hCoV-19/Qatar/QA-QU_02-3-11/2020 | EPI_ISL_1712708 |
| hCoV-19/Qatar/QA-QU_02-3-28/2020 | EPI_ISL_1712713 |
| hCoV-19/Qatar/QA-QU_02-3-32/2020 | EPI_ISL_1712714 |
| hCoV-19/Qatar/QA-QU_02-3-45/2020 | EPI_ISL_1712721 |
| hCoV-19/Qatar/QA-QU_02-2-1/2020 | EPI_ISL_1712684 |
| hCoV-19/Qatar/QA-QU_02-2-10/2020 | EPI_ISL_1712685 |
| hCoV-19/Qatar/QA-QU_02-2-20/2020 | EPI_ISL_1712687 |
| hCoV-19/Qatar/QA-QU_02-2-30/2020 | EPI_ISL_1712693 |
| hCoV-19/Qatar/QA-QU_02-2-31/2020 | EPI_ISL_1712694 |
| hCoV-19/Qatar/QA-QU_02-2-40/2020 | EPI_ISL_1712698 |
| hCoV-19/Qatar/QA-QU_02-2-9/2020 | EPI_ISL_1712707 |
| hCoV-19/Qatar/QA-QU_02-3-13/2020 | EPI_ISL_1712709 |
| hCoV-19/Qatar/QA-QU_02-3-15/2020 | EPI_ISL_1712711 |
| hCoV-19/Qatar/QA-QU_02-3-42/2020 | EPI_ISL_1712719 |
| hCoV-19/Qatar/QA-QU_02-3-50/2020 | EPI_ISL_1712723 |
| hCoV-19/Qatar/QA-QU_02-3-51/2020 | EPI_ISL_1712724 |
| hCoV-19/Qatar/QA-QU_02-3-58/2020 | EPI_ISL_1712727 |
| hCoV-19/Qatar/QA-QU_02-2-21/2020 | EPI_ISL_1712688 |

| | |
|---|---|
| hCoV-19/Qatar/QA-QU_02-3-47/2020 | EPI_ISL_1712722 |
| hCoV-19/Qatar/QA-QU_02-3-52/2020 | EPI_ISL_1712725 |
| hCoV-19/Qatar/QA-QU_02-3-54/2020 | EPI_ISL_1712726 |
| hCoV-19/Qatar/QA-QU_02-3-61/2020 | EPI_ISL_1712728 |
| hCoV-19/Qatar/QA-QU_02-2-46/2020 | EPI_ISL_1712700 |
| hCoV-19/Qatar/QA-QU_01-8/2020 | EPI_ISL_1712670 |
| hCoV-19/Qatar/QA-QU_02-2-26/2020 | EPI_ISL_1712690 |
| hCoV-19/Qatar/QA-QU_02-3-34/2020 | EPI_ISL_1712716 |
| hCoV-19/Qatar/QA-QU_09-1-E3/2020 | EPI_ISL_1713096 |
| hCoV-19/Qatar/QA-QU_09-1-G8/2020 | EPI_ISL_1713115 |
| hCoV-19/Qatar/QA-QU_09-1-H2/2020 | EPI_ISL_1713117 |
| hCoV-19/Qatar/QA-QU_09-1-A7/2020 | EPI_ISL_1713078 |
| hCoV-19/Qatar/QA-QU_09-1-G1/2020 | EPI_ISL_1713110 |
| hCoV-19/Qatar/QA-WCMQ_FD18215049/2020 | EPI_ISL_1714648 |
| hCoV-19/Qatar/QA-QU_02-1-1/2020 | EPI_ISL_1712671 |
| hCoV-19/Qatar/QA-QU_02-1-7/2020 | EPI_ISL_1712683 |
| hCoV-19/Qatar/QA-QU_09-1-E10/2020 | EPI_ISL_1713094 |
| hCoV-19/Qatar/QA-QU_09-1-H8/2020 | EPI_ISL_1713121 |
| hCoV-19/Qatar/QA-QU_02-1-13/2020 | EPI_ISL_1712672 |
| hCoV-19/Qatar/QA-QU_02-1-4/2020 | EPI_ISL_1712680 |
| hCoV-19/Qatar/QA-QU_03-H9/2020 | EPI_ISL_1712786 |
| hCoV-19/Qatar/QA-QU_02-1-14/2020 | EPI_ISL_1712673 |
| hCoV-19/Qatar/QA-QU_02-1-17/2020 | EPI_ISL_1712674 |
| hCoV-19/Qatar/QA-QU_02-1-5/2020 | EPI_ISL_1712681 |
| hCoV-19/Qatar/QA-WCMQ_FD15651317/2020 | EPI_ISL_1714134 |
| hCoV-19/Qatar/QA-QU_02-1-18/2020 | EPI_ISL_1712675 |
| hCoV-19/Qatar/QA-QU_02-1-6/2020 | EPI_ISL_1712682 |
| hCoV-19/Qatar/QA-QU_02-1-23/2020 | EPI_ISL_1712677 |
| hCoV-19/Qatar/QA-QU_02-1-25/2020 | EPI_ISL_1712678 |
| hCoV-19/Qatar/QA-QU_03-A11/2020 | EPI_ISL_1712731 |
| hCoV-19/Qatar/QA-QU_09-1-A2/2020 | EPI_ISL_1713074 |
| hCoV-19/Qatar/QA-QU_09-1-A9/2020 | EPI_ISL_1713080 |
| hCoV-19/Qatar/QA-QU_09-1-F3/2020 | EPI_ISL_1713104 |
| hCoV-19/Qatar/QA-QU_09-1-F4/2020 | EPI_ISL_1713105 |
| hCoV-19/Qatar/QA-QU_09-1-F7/2020 | EPI_ISL_1713107 |
| hCoV-19/Qatar/QA-QU_09-1-F8/2020 | EPI_ISL_1713108 |
| hCoV-19/Qatar/QA-QU_09-1-H12/2020 | EPI_ISL_1713116 |
| hCoV-19/Qatar/QA-QU_09-1-H6/2020 | EPI_ISL_1713119 |
| hCoV-19/Qatar/QA-QU_02-1-2/2020 | EPI_ISL_1712676 |
| hCoV-19/Qatar/QA-QU_02-1-3/2020 | EPI_ISL_1712679 |
| hCoV-19/Qatar/QA-QU_03-B12/2020 | EPI_ISL_1712741 |
| hCoV-19/Qatar/QA-QU_09-1-A6/2020 | EPI_ISL_1713077 |
| hCoV-19/Qatar/QA-QU_09-1-F12/2020 | EPI_ISL_1713102 |
| hCoV-19/Qatar/QA-WCMQ_FD15506506/2020 | EPI_ISL_1713954 |
| hCoV-19/Qatar/QA-WCMQ_FD15506537/2020 | EPI_ISL_1713985 |
| hCoV-19/Qatar/QA-WCMQ_FD15506566/2020 | EPI_ISL_1714014 |
| hCoV-19/Qatar/QA-QU_12-4-B9/2020 | EPI_ISL_1713288 |
| hCoV-19/Qatar/QA-WCMQ_FD15506502/2020 | EPI_ISL_1713950 |
| hCoV-19/Qatar/QA-WCMQ_FD15506540/2020 | EPI_ISL_1713988 |
| hCoV-19/Qatar/QA-WCMQ_FD15506521/2020 | EPI_ISL_1713969 |

| | |
|---|---|
| hCoV-19/Qatar/QA-WCMQ_FD18272693/2020 | EPI_ISL_1714686 |
| hCoV-19/Qatar/QA-QU_09-1-G6/2020 | EPI_ISL_1713113 |
| hCoV-19/Qatar/QA-WCMQ_FD15506504/2020 | EPI_ISL_1713952 |
| hCoV-19/Qatar/QA-WCMQ_FD15506525/2020 | EPI_ISL_1713973 |
| hCoV-19/Qatar/QA-WCMQ_FD18272684/2020 | EPI_ISL_1714678 |
| hCoV-19/Qatar/QA-WCMQ_FD18365000/2020 | EPI_ISL_1714745 |
| hCoV-19/Qatar/QA-QU_09-1-C8/2020 | EPI_ISL_1713090 |
| hCoV-19/Qatar/QA-WCMQ_FD18215263/2020 | EPI_ISL_1714649 |
| hCoV-19/Qatar/QA-QU_03-H12/2020 | EPI_ISL_1712782 |
| hCoV-19/Qatar/QA-QU_09-1-A8/2020 | EPI_ISL_1713079 |
| hCoV-19/Qatar/QA-QU_09-1-C10/2020 | EPI_ISL_1713086 |
| hCoV-19/Qatar/QA-QU_09-1-E8/2020 | EPI_ISL_1713098 |
| hCoV-19/Qatar/QA-QU_09-1-F10/2020 | EPI_ISL_1713100 |
| hCoV-19/Qatar/QA-QU_09-2-E3/2020 | EPI_ISL_1713131 |
| hCoV-19/Qatar/QA-QU_09-1-B1/2020 | EPI_ISL_1713081 |
| hCoV-19/Qatar/QA-QU_09-1-D8/2020 | EPI_ISL_1713092 |
| hCoV-19/Qatar/QA-WCMQ_FD18365042/2020 | EPI_ISL_1714746 |
| hCoV-19/Qatar/QA-QU_09-1-E7/2020 | EPI_ISL_1713097 |
| hCoV-19/Qatar/QA-QU_09-1-G7/2020 | EPI_ISL_1713114 |
| hCoV-19/Qatar/QA-WCMQ_FD15506552/2020 | EPI_ISL_1714000 |
| hCoV-19/Qatar/QA-QU_09-1-B3/2020 | EPI_ISL_1713083 |
| hCoV-19/Qatar/QA-QU_09-1-C7/2020 | EPI_ISL_1713089 |
| hCoV-19/Qatar/QA-WCMQ_FD15506541/2020 | EPI_ISL_1713989 |
| hCoV-19/Qatar/QA-WCMQ_FD18270879/2020 | EPI_ISL_1714673 |
| hCoV-19/Qatar/QA-QU_09-1-G4/2020 | EPI_ISL_1713112 |
| hCoV-19/Qatar/QA-QU_09-2-B12/2020 | EPI_ISL_1713123 |
| hCoV-19/Qatar/QA-WCMQ_FD15506547/2020 | EPI_ISL_1713995 |
| hCoV-19/Qatar/QA-WCMQ_FD15506578/2020 | EPI_ISL_1714026 |
| hCoV-19/Qatar/QA-WCMQ_FD15506581/2020 | EPI_ISL_1714029 |
| hCoV-19/Qatar/QA-WCMQ_FD15506582/2020 | EPI_ISL_1714030 |
| hCoV-19/Qatar/QA-WCMQ_FD15506584/2020 | EPI_ISL_1714032 |
| hCoV-19/Qatar/QA-QU_09-1-E9/2020 | EPI_ISL_1713099 |
| hCoV-19/Qatar/QA-QU_09-1-H7/2020 | EPI_ISL_1713120 |
| hCoV-19/Qatar/QA-QU_09-2-C4/2020 | EPI_ISL_1713127 |
| hCoV-19/Qatar/QA-WCMQ_FD15506507/2020 | EPI_ISL_1713955 |
| hCoV-19/Qatar/QA-WCMQ_FD15506534/2020 | EPI_ISL_1713982 |
| hCoV-19/Qatar/QA-WCMQ_FD15506546/2020 | EPI_ISL_1713994 |
| hCoV-19/Qatar/QA-WCMQ_FD15506567/2020 | EPI_ISL_1714015 |
| hCoV-19/Qatar/QA-WCMQ_FD18272703/2020 | EPI_ISL_1714695 |
| hCoV-19/Qatar/QA-QU_09-1-A11/2020 | EPI_ISL_1713072 |
| hCoV-19/Qatar/QA-QU_09-2-C9/2020 | EPI_ISL_1713128 |
| hCoV-19/Qatar/QA-QU_09-2-H1/2020 | EPI_ISL_1713137 |
| hCoV-19/Qatar/QA-WCMQ_FD15530973/2020 | EPI_ISL_1714047 |
| hCoV-19/Qatar/QA-WCMQ_FD18272690/2020 | EPI_ISL_1714683 |
| hCoV-19/Qatar/QA-WCMQ_FD18272767/2020 | EPI_ISL_1714739 |
| hCoV-19/Qatar/QA-QU_09-1-E12/2020 | EPI_ISL_1713095 |
| hCoV-19/Qatar/QA-QU_09-1-F6/2020 | EPI_ISL_1713106 |
| hCoV-19/Qatar/QA-WCMQ_FD15506523/2020 | EPI_ISL_1713971 |
| hCoV-19/Qatar/QA-WCMQ_FD15506558/2020 | EPI_ISL_1714006 |
| hCoV-19/Qatar/QA-WCMQ_FD15506577/2020 | EPI_ISL_1714025 |

| | |
|---|---|
| hCoV-19/Qatar/QA-WCMQ_FD15533261/2020 | EPI_ISL_1714048 |
| hCoV-19/Qatar/QA-WCMQ_FD18272719/2020 | EPI_ISL_1714705 |
| hCoV-19/Qatar/QA-QU_04-B3/2020 | EPI_ISL_1712800 |
| hCoV-19/Qatar/QA-QU_09-1-B7/2020 | EPI_ISL_1713084 |
| hCoV-19/Qatar/QA-QU_09-1-G3/2020 | EPI_ISL_1713111 |
| hCoV-19/Qatar/QA-WCMQ_FD15506515/2020 | EPI_ISL_1713963 |
| hCoV-19/Qatar/QA-WCMQ_FD15506522/2020 | EPI_ISL_1713970 |
| hCoV-19/Qatar/QA-WCMQ_FD15506524/2020 | EPI_ISL_1713972 |
| hCoV-19/Qatar/QA-WCMQ_FD15506549/2020 | EPI_ISL_1713997 |
| hCoV-19/Qatar/QA-WCMQ_FD15506550/2020 | EPI_ISL_1713998 |
| hCoV-19/Qatar/QA-WCMQ_FD15506580/2020 | EPI_ISL_1714028 |
| hCoV-19/Qatar/QA-WCMQ_FD15506585/2020 | EPI_ISL_1714033 |
| hCoV-19/Qatar/QA-WCMQ_FD15506590/2020 | EPI_ISL_1714038 |
| hCoV-19/Qatar/QA-WCMQ_FD15540726/2020 | EPI_ISL_1714067 |
| hCoV-19/Qatar/QA-QU_03-B11/2020 | EPI_ISL_1712740 |
| hCoV-19/Qatar/QA-QU_09-1-A3/2020 | EPI_ISL_1713075 |
| hCoV-19/Qatar/QA-QU_09-2-E12/2020 | EPI_ISL_1713130 |
| hCoV-19/Qatar/QA-WCMQ_FD15506503/2020 | EPI_ISL_1713951 |
| hCoV-19/Qatar/QA-WCMQ_FD15506510/2020 | EPI_ISL_1713958 |
| hCoV-19/Qatar/QA-WCMQ_FD15506512/2020 | EPI_ISL_1713960 |
| hCoV-19/Qatar/QA-WCMQ_FD15506520/2020 | EPI_ISL_1713968 |
| hCoV-19/Qatar/QA-WCMQ_FD15506526/2020 | EPI_ISL_1713974 |
| hCoV-19/Qatar/QA-WCMQ_FD15506528/2020 | EPI_ISL_1713976 |
| hCoV-19/Qatar/QA-WCMQ_FD15506531/2020 | EPI_ISL_1713979 |
| hCoV-19/Qatar/QA-WCMQ_FD15506543/2020 | EPI_ISL_1713991 |
| hCoV-19/Qatar/QA-WCMQ_FD15506544/2020 | EPI_ISL_1713992 |
| hCoV-19/Qatar/QA-WCMQ_FD15506545/2020 | EPI_ISL_1713993 |
| hCoV-19/Qatar/QA-WCMQ_FD15506548/2020 | EPI_ISL_1713996 |
| hCoV-19/Qatar/QA-WCMQ_FD15506553/2020 | EPI_ISL_1714001 |
| hCoV-19/Qatar/QA-WCMQ_FD15506560/2020 | EPI_ISL_1714008 |
| hCoV-19/Qatar/QA-WCMQ_FD15506562/2020 | EPI_ISL_1714010 |
| hCoV-19/Qatar/QA-WCMQ_FD15506572/2020 | EPI_ISL_1714020 |
| hCoV-19/Qatar/QA-WCMQ_FD15506573/2020 | EPI_ISL_1714021 |
| hCoV-19/Qatar/QA-WCMQ_FD15506575/2020 | EPI_ISL_1714023 |
| hCoV-19/Qatar/QA-WCMQ_FD15506576/2020 | EPI_ISL_1714024 |
| hCoV-19/Qatar/QA-WCMQ_FD15506583/2020 | EPI_ISL_1714031 |
| hCoV-19/Qatar/QA-WCMQ_FD15506587/2020 | EPI_ISL_1714035 |
| hCoV-19/Qatar/QA-WCMQ_FD15506588/2020 | EPI_ISL_1714036 |
| hCoV-19/Qatar/QA-WCMQ_FD15540695/2020 | EPI_ISL_1714055 |
| hCoV-19/Qatar/QA-WCMQ_FD15540712/2020 | EPI_ISL_1714059 |
| hCoV-19/Qatar/QA-WCMQ_FD15540718/2020 | EPI_ISL_1714065 |
| hCoV-19/Qatar/QA-WCMQ_FD18160888/2020 | EPI_ISL_1714357 |
| hCoV-19/Qatar/QA-WCMQ_FD18216975/2020 | EPI_ISL_1714665 |
| hCoV-19/Qatar/QA-WCMQ_FD18272725/2020 | EPI_ISL_1714710 |
| hCoV-19/Qatar/QA-WCMQ_FD18272726/2020 | EPI_ISL_1714711 |
| hCoV-19/Qatar/QA-WCMQ_FD18272734/2020 | EPI_ISL_1714718 |
| hCoV-19/Qatar/QA-QU_04-B2/2020 | EPI_ISL_1712799 |
| hCoV-19/Qatar/QA-QU_09-1-B9/2020 | EPI_ISL_1713085 |
| hCoV-19/Qatar/QA-QU_09-1-E1/2020 | EPI_ISL_1713093 |
| hCoV-19/Qatar/QA-QU_09-2-B10/2020 | EPI_ISL_1713122 |

| | |
|---|---|
| hCoV-19/Qatar/QA-QU_09-2-F3/2020 | EPI_ISL_1713136 |
| hCoV-19/Qatar/QA-QU_09-2-H3/2020 | EPI_ISL_1713138 |
| hCoV-19/Qatar/QA-WCMQ_FD15506505/2020 | EPI_ISL_1713953 |
| hCoV-19/Qatar/QA-WCMQ_FD15506511/2020 | EPI_ISL_1713959 |
| hCoV-19/Qatar/QA-WCMQ_FD15506513/2020 | EPI_ISL_1713961 |
| hCoV-19/Qatar/QA-WCMQ_FD15506514/2020 | EPI_ISL_1713962 |
| hCoV-19/Qatar/QA-WCMQ_FD15506518/2020 | EPI_ISL_1713966 |
| hCoV-19/Qatar/QA-WCMQ_FD15506529/2020 | EPI_ISL_1713977 |
| hCoV-19/Qatar/QA-WCMQ_FD15506530/2020 | EPI_ISL_1713978 |
| hCoV-19/Qatar/QA-WCMQ_FD15506542/2020 | EPI_ISL_1713990 |
| hCoV-19/Qatar/QA-WCMQ_FD15506574/2020 | EPI_ISL_1714022 |
| hCoV-19/Qatar/QA-WCMQ_FD15506579/2020 | EPI_ISL_1714027 |
| hCoV-19/Qatar/QA-WCMQ_FD15506589/2020 | EPI_ISL_1714037 |
| hCoV-19/Qatar/QA-WCMQ_FD15506591/2020 | EPI_ISL_1714039 |
| hCoV-19/Qatar/QA-WCMQ_FD15506593/2020 | EPI_ISL_1714041 |
| hCoV-19/Qatar/QA-WCMQ_FD15506597/2020 | EPI_ISL_1714045 |
| hCoV-19/Qatar/QA-WCMQ_FD15617953/2020 | EPI_ISL_1714109 |
| hCoV-19/Qatar/QA-WCMQ_FD15617970/2020 | EPI_ISL_1714123 |
| hCoV-19/Qatar/QA-WCMQ_FD15617972/2020 | EPI_ISL_1714124 |
| hCoV-19/Qatar/QA-WCMQ_FD18160883/2020 | EPI_ISL_1714356 |
| hCoV-19/Qatar/QA-WCMQ_FD18272687/2020 | EPI_ISL_1714681 |
| hCoV-19/Qatar/QA-WCMQ_FD18272695/2020 | EPI_ISL_1714688 |
| hCoV-19/Qatar/QA-WCMQ_FD18272715/2020 | EPI_ISL_1714703 |
| hCoV-19/Qatar/QA-QU_12-4-B8/2020 | EPI_ISL_1713287 |
| hCoV-19/Qatar/QA-WCMQ_FD15506516/2020 | EPI_ISL_1713964 |
| hCoV-19/Qatar/QA-WCMQ_FD15506527/2020 | EPI_ISL_1713975 |
| hCoV-19/Qatar/QA-WCMQ_FD15506561/2020 | EPI_ISL_1714009 |
| hCoV-19/Qatar/QA-WCMQ_FD15617909/2020 | EPI_ISL_1714088 |
| hCoV-19/Qatar/QA-WCMQ_FD15617926/2020 | EPI_ISL_1714097 |
| hCoV-19/Qatar/QA-WCMQ_FD15617927/2020 | EPI_ISL_1714098 |
| hCoV-19/Qatar/QA-WCMQ_FD15617929/2020 | EPI_ISL_1714099 |
| hCoV-19/Qatar/QA-WCMQ_FD15617933/2020 | EPI_ISL_1714101 |
| hCoV-19/Qatar/QA-WCMQ_FD15617938/2020 | EPI_ISL_1714103 |
| hCoV-19/Qatar/QA-WCMQ_FD15617939/2020 | EPI_ISL_1714104 |
| hCoV-19/Qatar/QA-WCMQ_FD15617942/2020 | EPI_ISL_1714105 |
| hCoV-19/Qatar/QA-WCMQ_FD15617946/2020 | EPI_ISL_1714106 |
| hCoV-19/Qatar/QA-WCMQ_FD15617955/2020 | EPI_ISL_1714111 |
| hCoV-19/Qatar/QA-WCMQ_FD15617966/2020 | EPI_ISL_1714119 |
| hCoV-19/Qatar/QA-WCMQ_FD15617967/2020 | EPI_ISL_1714120 |
| hCoV-19/Qatar/QA-WCMQ_FD15617968/2020 | EPI_ISL_1714121 |
| hCoV-19/Qatar/QA-WCMQ_FD18272685/2020 | EPI_ISL_1714679 |
| hCoV-19/Qatar/QA-WCMQ_FD18272686/2020 | EPI_ISL_1714680 |
| hCoV-19/Qatar/QA-WCMQ_FD18272689/2020 | EPI_ISL_1714682 |
| hCoV-19/Qatar/QA-WCMQ_FD18272718/2020 | EPI_ISL_1714704 |
| hCoV-19/Qatar/QA-WCMQ_FD18272723/2020 | EPI_ISL_1714709 |
| hCoV-19/Qatar/QA-WCMQ_FD18272735/2020 | EPI_ISL_1714719 |
| hCoV-19/Qatar/QA-QU_09-1-B12/2020 | EPI_ISL_1713082 |
| hCoV-19/Qatar/QA-QU_09-1-F9/2020 | EPI_ISL_1713109 |
| hCoV-19/Qatar/QA-QU_12-4-A12/2020 | EPI_ISL_1713286 |
| hCoV-19/Qatar/QA-WCMQ_FD15506532/2020 | EPI_ISL_1713980 |

| | |
|---|---|
| hCoV-19/Qatar/QA-WCMQ_FD15506533/2020 | EPI_ISL_1713981 |
| hCoV-19/Qatar/QA-WCMQ_FD15506536/2020 | EPI_ISL_1713984 |
| hCoV-19/Qatar/QA-WCMQ_FD15506559/2020 | EPI_ISL_1714007 |
| hCoV-19/Qatar/QA-WCMQ_FD15506586/2020 | EPI_ISL_1714034 |
| hCoV-19/Qatar/QA-WCMQ_FD15506592/2020 | EPI_ISL_1714040 |
| hCoV-19/Qatar/QA-WCMQ_FD15506595/2020 | EPI_ISL_1714043 |
| hCoV-19/Qatar/QA-WCMQ_FD15506596/2020 | EPI_ISL_1714044 |
| hCoV-19/Qatar/QA-WCMQ_FD15540699/2020 | EPI_ISL_1714056 |
| hCoV-19/Qatar/QA-WCMQ_FD15540702/2020 | EPI_ISL_1714058 |
| hCoV-19/Qatar/QA-WCMQ_FD15540713/2020 | EPI_ISL_1714060 |
| hCoV-19/Qatar/QA-WCMQ_FD15540714/2020 | EPI_ISL_1714061 |
| hCoV-19/Qatar/QA-WCMQ_FD15540715/2020 | EPI_ISL_1714062 |
| hCoV-19/Qatar/QA-WCMQ_FD15540716/2020 | EPI_ISL_1714063 |
| hCoV-19/Qatar/QA-WCMQ_FD15617880/2020 | EPI_ISL_1714078 |
| hCoV-19/Qatar/QA-WCMQ_FD15617881/2020 | EPI_ISL_1714079 |
| hCoV-19/Qatar/QA-WCMQ_FD15617882/2020 | EPI_ISL_1714080 |
| hCoV-19/Qatar/QA-WCMQ_FD15617884/2020 | EPI_ISL_1714081 |
| hCoV-19/Qatar/QA-WCMQ_FD15617893/2020 | EPI_ISL_1714082 |
| hCoV-19/Qatar/QA-WCMQ_FD15617894/2020 | EPI_ISL_1714083 |
| hCoV-19/Qatar/QA-WCMQ_FD15617901/2020 | EPI_ISL_1714084 |
| hCoV-19/Qatar/QA-WCMQ_FD15617902/2020 | EPI_ISL_1714085 |
| hCoV-19/Qatar/QA-WCMQ_FD15617907/2020 | EPI_ISL_1714086 |
| hCoV-19/Qatar/QA-WCMQ_FD15617908/2020 | EPI_ISL_1714087 |
| hCoV-19/Qatar/QA-WCMQ_FD15617910/2020 | EPI_ISL_1714089 |
| hCoV-19/Qatar/QA-WCMQ_FD15617912/2020 | EPI_ISL_1714090 |
| hCoV-19/Qatar/QA-WCMQ_FD15617914/2020 | EPI_ISL_1714091 |
| hCoV-19/Qatar/QA-WCMQ_FD15617917/2020 | EPI_ISL_1714092 |
| hCoV-19/Qatar/QA-WCMQ_FD15617918/2020 | EPI_ISL_1714093 |
| hCoV-19/Qatar/QA-WCMQ_FD15617919/2020 | EPI_ISL_1714094 |
| hCoV-19/Qatar/QA-WCMQ_FD15617920/2020 | EPI_ISL_1714095 |
| hCoV-19/Qatar/QA-WCMQ_FD15617922/2020 | EPI_ISL_1714096 |
| hCoV-19/Qatar/QA-WCMQ_FD15617930/2020 | EPI_ISL_1714100 |
| hCoV-19/Qatar/QA-WCMQ_FD15617934/2020 | EPI_ISL_1714102 |
| hCoV-19/Qatar/QA-WCMQ_FD15617949/2020 | EPI_ISL_1714107 |
| hCoV-19/Qatar/QA-WCMQ_FD15617952/2020 | EPI_ISL_1714108 |
| hCoV-19/Qatar/QA-WCMQ_FD15617954/2020 | EPI_ISL_1714110 |
| hCoV-19/Qatar/QA-WCMQ_FD15617956/2020 | EPI_ISL_1714112 |
| hCoV-19/Qatar/QA-WCMQ_FD15617957/2020 | EPI_ISL_1714113 |
| hCoV-19/Qatar/QA-WCMQ_FD15617958/2020 | EPI_ISL_1714114 |
| hCoV-19/Qatar/QA-WCMQ_FD15617959/2020 | EPI_ISL_1714115 |
| hCoV-19/Qatar/QA-WCMQ_FD15617960/2020 | EPI_ISL_1714116 |
| hCoV-19/Qatar/QA-WCMQ_FD15617963/2020 | EPI_ISL_1714117 |
| hCoV-19/Qatar/QA-WCMQ_FD15617964/2020 | EPI_ISL_1714118 |
| hCoV-19/Qatar/QA-WCMQ_FD15617969/2020 | EPI_ISL_1714122 |
| hCoV-19/Qatar/QA-WCMQ_FD18272707/2020 | EPI_ISL_1714696 |
| hCoV-19/Qatar/QA-WCMQ_FD18272708/2020 | EPI_ISL_1714697 |
| hCoV-19/Qatar/QA-WCMQ_FD18272709/2020 | EPI_ISL_1714698 |
| hCoV-19/Qatar/QA-WCMQ_FD18272710/2020 | EPI_ISL_1714699 |
| hCoV-19/Qatar/QA-WCMQ_FD18272711/2020 | EPI_ISL_1714700 |
| hCoV-19/Qatar/QA-WCMQ_FD18272712/2020 | EPI_ISL_1714701 |

| | |
|---|---|
| hCoV-19/Qatar/QA-WCMQ_FD18272713/2020 | EPI_ISL_1714702 |
| hCoV-19/Qatar/QA-WCMQ_FD18272768/2020 | EPI_ISL_1714740 |
| hCoV-19/Qatar/QA-QU_05-J8/2020 | EPI_ISL_1712895 |
| hCoV-19/Qatar/QA-WCMQ_FD15506508/2020 | EPI_ISL_1713956 |
| hCoV-19/Qatar/QA-WCMQ_FD15506509/2020 | EPI_ISL_1713957 |
| hCoV-19/Qatar/QA-WCMQ_FD15506519/2020 | EPI_ISL_1713967 |
| hCoV-19/Qatar/QA-WCMQ_FD15506535/2020 | EPI_ISL_1713983 |
| hCoV-19/Qatar/QA-WCMQ_FD15506538/2020 | EPI_ISL_1713986 |
| hCoV-19/Qatar/QA-WCMQ_FD15506539/2020 | EPI_ISL_1713987 |
| hCoV-19/Qatar/QA-WCMQ_FD15506551/2020 | EPI_ISL_1713999 |
| hCoV-19/Qatar/QA-WCMQ_FD15506563/2020 | EPI_ISL_1714011 |
| hCoV-19/Qatar/QA-WCMQ_FD15506564/2020 | EPI_ISL_1714012 |
| hCoV-19/Qatar/QA-WCMQ_FD15506569/2020 | EPI_ISL_1714017 |
| hCoV-19/Qatar/QA-WCMQ_FD15506570/2020 | EPI_ISL_1714018 |
| hCoV-19/Qatar/QA-WCMQ_FD15506571/2020 | EPI_ISL_1714019 |
| hCoV-19/Qatar/QA-WCMQ_FD15506594/2020 | EPI_ISL_1714042 |
| hCoV-19/Qatar/QA-WCMQ_FD15540717/2020 | EPI_ISL_1714064 |
| hCoV-19/Qatar/QA-WCMQ_FD18365058/2020 | EPI_ISL_1714749 |
| hCoV-19/Qatar/QA-QU_09-1-F2/2020 | EPI_ISL_1713103 |
| hCoV-19/Qatar/QA-QU_09-1-A5/2020 | EPI_ISL_1713076 |
| hCoV-19/Qatar/QA-QU_09-1-F11/2020 | EPI_ISL_1713101 |
| hCoV-19/Qatar/QA-QU_09-1-H5/2020 | EPI_ISL_1713118 |
| hCoV-19/Qatar/QA-QU_09-2-C3/2020 | EPI_ISL_1713126 |
| hCoV-19/Qatar/QA-WCMQ_FD15506517/2020 | EPI_ISL_1713965 |
| hCoV-19/Qatar/QA-WCMQ_FD15506556/2020 | EPI_ISL_1714004 |
| hCoV-19/Qatar/QA-WCMQ_FD15506565/2020 | EPI_ISL_1714013 |
| hCoV-19/Qatar/QA-WCMQ_FD15506568/2020 | EPI_ISL_1714016 |
| hCoV-19/Qatar/QA-QU_09-1-A12/2020 | EPI_ISL_1713073 |
| hCoV-19/Qatar/QA-QU_09-2-C2/2020 | EPI_ISL_1713125 |
| hCoV-19/Qatar/QA-WCMQ_FD15506554/2020 | EPI_ISL_1714002 |
| hCoV-19/Qatar/QA-WCMQ_FD15506555/2020 | EPI_ISL_1714003 |
| hCoV-19/Qatar/QA-WCMQ_FD15506557/2020 | EPI_ISL_1714005 |
| hCoV-19/Qatar/QA-QU_04-B9/2020 | EPI_ISL_1712803 |
| hCoV-19/Qatar/QA-QU_04-H10/2020 | EPI_ISL_1712823 |
| hCoV-19/Qatar/QA-WCMQ_FD15540722/2020 | EPI_ISL_1714066 |
| hCoV-19/Qatar/QA-WCMQ_FD18160893/2020 | EPI_ISL_1714359 |
| hCoV-19/Qatar/QA-WCMQ_FD18193845/2020 | EPI_ISL_1714573 |
| hCoV-19/Qatar/QA-QU_04-D10/2020 | EPI_ISL_1712805 |
| hCoV-19/Qatar/QA-WCMQ_FD15540727/2020 | EPI_ISL_1714068 |
| hCoV-19/Qatar/QA-WCMQ_FD15651304/2020 | EPI_ISL_1714133 |
| hCoV-19/Qatar/QA-WCMQ_FD15651341/2020 | EPI_ISL_1714137 |
| hCoV-19/Qatar/QA-WCMQ_FD17438193/2020 | EPI_ISL_1714186 |
| hCoV-19/Qatar/QA-WCMQ_FD18193844/2020 | EPI_ISL_1714572 |
| hCoV-19/Qatar/QA-WCMQ_FD18272696/2020 | EPI_ISL_1714689 |
| hCoV-19/Qatar/QA-QU_09-2-E9/2020 | EPI_ISL_1713134 |
| hCoV-19/Qatar/QA-WCMQ_FD18160892/2020 | EPI_ISL_1714358 |
| hCoV-19/Qatar/QA-WCMQ_FD18160894/2020 | EPI_ISL_1714360 |
| hCoV-19/Qatar/QA-WCMQ_FD18193908/2020 | EPI_ISL_1714635 |
| hCoV-19/Qatar/QA-QU_09-1-D12/2020 | EPI_ISL_1713091 |
| hCoV-19/Qatar/QA-QU_09-2-C1/2020 | EPI_ISL_1713124 |

| | |
|---|---|
| hCoV-19/Qatar/QA-WCMQ_FD15651515/2020 | EPI_ISL_1714138 |
| hCoV-19/Qatar/QA-WCMQ_FD18160912/2020 | EPI_ISL_1714363 |
| hCoV-19/Qatar/QA-WCMQ_FD18160916/2020 | EPI_ISL_1714364 |
| hCoV-19/Qatar/QA-WCMQ_FD18193884/2020 | EPI_ISL_1714611 |
| hCoV-19/Qatar/QA-QU_05-E4/2020 | EPI_ISL_1712856 |
| hCoV-19/Qatar/QA-WCMQ_FD18193849/2020 | EPI_ISL_1714577 |
| hCoV-19/Qatar/QA-WCMQ_FD18193870/2020 | EPI_ISL_1714597 |
| hCoV-19/Qatar/QA-WCMQ_FD18193872/2020 | EPI_ISL_1714599 |
| hCoV-19/Qatar/QA-WCMQ_FD18193873/2020 | EPI_ISL_1714600 |
| hCoV-19/Qatar/QA-WCMQ_FD18193883/2020 | EPI_ISL_1714610 |
| hCoV-19/Qatar/QA-QU_03-H11/2020 | EPI_ISL_1712781 |
| hCoV-19/Qatar/QA-QU_03-H7/2020 | EPI_ISL_1712785 |
| hCoV-19/Qatar/QA-QU_04-A4/2020 | EPI_ISL_1712794 |
| hCoV-19/Qatar/QA-WCMQ_FD18193835/2020 | EPI_ISL_1714563 |
| hCoV-19/Qatar/QA-WCMQ_FD18193867/2020 | EPI_ISL_1714594 |
| hCoV-19/Qatar/QA-WCMQ_FD18270856/2020 | EPI_ISL_1714671 |
| hCoV-19/Qatar/QA-WCMQ_FD18270878/2020 | EPI_ISL_1714672 |
| hCoV-19/Qatar/QA-WCMQ_FD18270880/2020 | EPI_ISL_1714674 |
| hCoV-19/Qatar/QA-WCMQ_FD18365049/2020 | EPI_ISL_1714747 |
| hCoV-19/Qatar/QA-WCMQ_FD18365050/2020 | EPI_ISL_1714748 |
| hCoV-19/Qatar/QA-QU_03-B9/2020 | EPI_ISL_1712747 |
| hCoV-19/Qatar/QA-QU_03-D9/2020 | EPI_ISL_1712761 |
| hCoV-19/Qatar/QA-QU_03-E9/2020 | EPI_ISL_1712767 |
| hCoV-19/Qatar/QA-QU_03-F9/2020 | EPI_ISL_1712773 |
| hCoV-19/Qatar/QA-QU_03-S7/2020 | EPI_ISL_1712791 |
| hCoV-19/Qatar/QA-QU_03-S8/2020 | EPI_ISL_1712792 |
| hCoV-19/Qatar/QA-QU_09-2-F1/2020 | EPI_ISL_1713135 |
| hCoV-19/Qatar/QA-WCMQ_FD18193907/2020 | EPI_ISL_1714634 |
| hCoV-19/Qatar/QA-WCMQ_FD18272749/2020 | EPI_ISL_1714729 |
| hCoV-19/Qatar/QA-WCMQ_FD18272750/2020 | EPI_ISL_1714730 |
| hCoV-19/Qatar/QA-WCMQ_FD18272752/2020 | EPI_ISL_1714731 |
| hCoV-19/Qatar/QA-WCMQ_FD18272755/2020 | EPI_ISL_1714732 |
| hCoV-19/Qatar/QA-WCMQ_FD18272756/2020 | EPI_ISL_1714733 |
| hCoV-19/Qatar/QA-WCMQ_FD18272757/2020 | EPI_ISL_1714734 |
| hCoV-19/Qatar/QA-WCMQ_FD18272760/2020 | EPI_ISL_1714735 |
| hCoV-19/Qatar/QA-QU_03-A1/2020 | EPI_ISL_1712729 |
| hCoV-19/Qatar/QA-QU_03-A10/2020 | EPI_ISL_1712730 |
| hCoV-19/Qatar/QA-QU_03-A12/2020 | EPI_ISL_1712732 |
| hCoV-19/Qatar/QA-QU_03-A4/2020 | EPI_ISL_1712734 |
| hCoV-19/Qatar/QA-QU_03-A5/2020 | EPI_ISL_1712735 |
| hCoV-19/Qatar/QA-QU_03-A8/2020 | EPI_ISL_1712737 |
| hCoV-19/Qatar/QA-QU_03-A9/2020 | EPI_ISL_1712738 |
| hCoV-19/Qatar/QA-QU_03-B10/2020 | EPI_ISL_1712739 |
| hCoV-19/Qatar/QA-QU_03-B3/2020 | EPI_ISL_1712743 |
| hCoV-19/Qatar/QA-QU_03-B4/2020 | EPI_ISL_1712744 |
| hCoV-19/Qatar/QA-QU_03-B6/2020 | EPI_ISL_1712745 |
| hCoV-19/Qatar/QA-QU_03-B8/2020 | EPI_ISL_1712746 |
| hCoV-19/Qatar/QA-QU_03-C1/2020 | EPI_ISL_1712748 |
| hCoV-19/Qatar/QA-QU_03-C3/2020 | EPI_ISL_1712749 |
| hCoV-19/Qatar/QA-QU_03-C4/2020 | EPI_ISL_1712750 |

| | |
|---|---|
| hCoV-19/Qatar/QA-QU_03-C5/2020 | EPI_ISL_1712751 |
| hCoV-19/Qatar/QA-QU_03-C6/2020 | EPI_ISL_1712752 |
| hCoV-19/Qatar/QA-QU_03-C8/2020 | EPI_ISL_1712753 |
| hCoV-19/Qatar/QA-QU_03-D2/2020 | EPI_ISL_1712754 |
| hCoV-19/Qatar/QA-QU_03-D3/2020 | EPI_ISL_1712755 |
| hCoV-19/Qatar/QA-QU_03-D4/2020 | EPI_ISL_1712756 |
| hCoV-19/Qatar/QA-QU_03-D5/2020 | EPI_ISL_1712757 |
| hCoV-19/Qatar/QA-QU_03-D6/2020 | EPI_ISL_1712758 |
| hCoV-19/Qatar/QA-QU_03-D7/2020 | EPI_ISL_1712759 |
| hCoV-19/Qatar/QA-QU_03-D8/2020 | EPI_ISL_1712760 |
| hCoV-19/Qatar/QA-QU_03-E10/2020 | EPI_ISL_1712762 |
| hCoV-19/Qatar/QA-QU_03-E3/2020 | EPI_ISL_1712763 |
| hCoV-19/Qatar/QA-QU_03-E5/2020 | EPI_ISL_1712764 |
| hCoV-19/Qatar/QA-QU_03-E6/2020 | EPI_ISL_1712765 |
| hCoV-19/Qatar/QA-QU_03-E8/2020 | EPI_ISL_1712766 |
| hCoV-19/Qatar/QA-QU_03-F1/2020 | EPI_ISL_1712768 |
| hCoV-19/Qatar/QA-QU_03-F4/2020 | EPI_ISL_1712771 |
| hCoV-19/Qatar/QA-QU_03-F6/2020 | EPI_ISL_1712772 |
| hCoV-19/Qatar/QA-QU_03-G12/2020 | EPI_ISL_1712775 |
| hCoV-19/Qatar/QA-QU_03-G3/2020 | EPI_ISL_1712776 |
| hCoV-19/Qatar/QA-QU_03-G4/2020 | EPI_ISL_1712777 |
| hCoV-19/Qatar/QA-QU_03-G5/2020 | EPI_ISL_1712778 |
| hCoV-19/Qatar/QA-QU_03-G9/2020 | EPI_ISL_1712779 |
| hCoV-19/Qatar/QA-QU_03-H1/2020 | EPI_ISL_1712780 |
| hCoV-19/Qatar/QA-QU_03-H4/2020 | EPI_ISL_1712784 |
| hCoV-19/Qatar/QA-QU_03-S1/2020 | EPI_ISL_1712787 |
| hCoV-19/Qatar/QA-QU_03-S9/2020 | EPI_ISL_1712793 |
| hCoV-19/Qatar/QA-QU_09-2-E7/2020 | EPI_ISL_1713133 |
| hCoV-19/Qatar/QA-WCMQ_FD18272727/2020 | EPI_ISL_1714712 |
| hCoV-19/Qatar/QA-WCMQ_FD18272736/2020 | EPI_ISL_1714720 |
| hCoV-19/Qatar/QA-WCMQ_FD18272738/2020 | EPI_ISL_1714721 |
| hCoV-19/Qatar/QA-WCMQ_FD18272739/2020 | EPI_ISL_1714722 |
| hCoV-19/Qatar/QA-WCMQ_FD18272740/2020 | EPI_ISL_1714723 |
| hCoV-19/Qatar/QA-WCMQ_FD18272741/2020 | EPI_ISL_1714724 |
| hCoV-19/Qatar/QA-WCMQ_FD18272742/2020 | EPI_ISL_1714725 |
| hCoV-19/Qatar/QA-WCMQ_FD18272743/2020 | EPI_ISL_1714726 |
| hCoV-19/Qatar/QA-WCMQ_FD18272744/2020 | EPI_ISL_1714727 |
| hCoV-19/Qatar/QA-WCMQ_FD18272746/2020 | EPI_ISL_1714728 |
| hCoV-19/Qatar/QA-WCMQ_FD18272763/2020 | EPI_ISL_1714736 |
| hCoV-19/Qatar/QA-WCMQ_FD18272765/2020 | EPI_ISL_1714737 |
| hCoV-19/Qatar/QA-WCMQ_FD18272766/2020 | EPI_ISL_1714738 |
| hCoV-19/Qatar/QA-QU_03-A3/2020 | EPI_ISL_1712733 |
| hCoV-19/Qatar/QA-QU_03-A7/2020 | EPI_ISL_1712736 |
| hCoV-19/Qatar/QA-QU_03-B2/2020 | EPI_ISL_1712742 |
| hCoV-19/Qatar/QA-QU_03-F10/2020 | EPI_ISL_1712769 |
| hCoV-19/Qatar/QA-QU_03-F2/2020 | EPI_ISL_1712770 |
| hCoV-19/Qatar/QA-QU_03-G11/2020 | EPI_ISL_1712774 |
| hCoV-19/Qatar/QA-QU_03-H2/2020 | EPI_ISL_1712783 |
| hCoV-19/Qatar/QA-QU_03-S10/2020 | EPI_ISL_1712788 |
| hCoV-19/Qatar/QA-QU_03-S2/2020 | EPI_ISL_1712789 |

| | |
|---|---|
| hCoV-19/Qatar/QA-QU_03-S5/2020 | EPI_ISL_1712790 |
| hCoV-19/Qatar/QA-QU_09-2-E10/2020 | EPI_ISL_1713129 |
| hCoV-19/Qatar/QA-WCMQ_FD15481381/2020 | EPI_ISL_1713949 |
| hCoV-19/Qatar/QA-WCMQ_FD18193848/2020 | EPI_ISL_1714576 |
| hCoV-19/Qatar/QA-WCMQ_FD18272674/2020 | EPI_ISL_1714675 |
| hCoV-19/Qatar/QA-WCMQ_FD18272675/2020 | EPI_ISL_1714676 |
| hCoV-19/Qatar/QA-WCMQ_FD18272682/2020 | EPI_ISL_1714677 |
| hCoV-19/Qatar/QA-WCMQ_FD18272691/2020 | EPI_ISL_1714684 |
| hCoV-19/Qatar/QA-WCMQ_FD18272692/2020 | EPI_ISL_1714685 |
| hCoV-19/Qatar/QA-WCMQ_FD18272694/2020 | EPI_ISL_1714687 |
| hCoV-19/Qatar/QA-WCMQ_FD18272697/2020 | EPI_ISL_1714690 |
| hCoV-19/Qatar/QA-WCMQ_FD18272698/2020 | EPI_ISL_1714691 |
| hCoV-19/Qatar/QA-WCMQ_FD18272699/2020 | EPI_ISL_1714692 |
| hCoV-19/Qatar/QA-WCMQ_FD18272700/2020 | EPI_ISL_1714693 |
| hCoV-19/Qatar/QA-WCMQ_FD18272702/2020 | EPI_ISL_1714694 |
| hCoV-19/Qatar/QA-WCMQ_FD18272720/2020 | EPI_ISL_1714706 |
| hCoV-19/Qatar/QA-WCMQ_FD18272721/2020 | EPI_ISL_1714707 |
| hCoV-19/Qatar/QA-WCMQ_FD18272722/2020 | EPI_ISL_1714708 |
| hCoV-19/Qatar/QA-WCMQ_FD18272728/2020 | EPI_ISL_1714713 |
| hCoV-19/Qatar/QA-WCMQ_FD18272730/2020 | EPI_ISL_1714714 |
| hCoV-19/Qatar/QA-WCMQ_FD18272731/2020 | EPI_ISL_1714715 |
| hCoV-19/Qatar/QA-WCMQ_FD18272732/2020 | EPI_ISL_1714716 |
| hCoV-19/Qatar/QA-WCMQ_FD18272733/2020 | EPI_ISL_1714717 |
| hCoV-19/Qatar/QA-WCMQ_FD15535184/2020 | EPI_ISL_1714049 |
| hCoV-19/Qatar/QA-WCMQ_FD15381415/2020 | EPI_ISL_1713947 |
| hCoV-19/Qatar/QA-WCMQ_FD15382133/2020 | EPI_ISL_1713948 |
| hCoV-19/Qatar/QA-WCMQ_FD15540691/2020 | EPI_ISL_1714053 |
| hCoV-19/Qatar/QA-WCMQ_FD15540694/2020 | EPI_ISL_1714054 |
| hCoV-19/Qatar/QA-WCMQ_FD15540741/2020 | EPI_ISL_1714076 |
| hCoV-19/Qatar/QA-WCMQ_FD18254821/2020 | EPI_ISL_1714669 |
| hCoV-19/Qatar/QA-WCMQ_FD18254849/2020 | EPI_ISL_1714670 |
| hCoV-19/Qatar/QA-QU_05-G1/2020 | EPI_ISL_1712865 |
| hCoV-19/Qatar/QA-QU_05-I1/2020 | EPI_ISL_1712881 |
| hCoV-19/Qatar/QA-QU_05-J4/2020 | EPI_ISL_1712891 |
| hCoV-19/Qatar/QA-WCMQ_FD15540735/2020 | EPI_ISL_1714074 |
| hCoV-19/Qatar/QA-WCMQ_FD18215318/2020 | EPI_ISL_1714658 |
| hCoV-19/Qatar/QA-QU_04-B5/2020 | EPI_ISL_1712801 |
| hCoV-19/Qatar/QA-QU_04-B8/2020 | EPI_ISL_1712802 |
| hCoV-19/Qatar/QA-QU_05-A4/2020 | EPI_ISL_1712833 |
| hCoV-19/Qatar/QA-QU_05-E2/2020 | EPI_ISL_1712854 |
| hCoV-19/Qatar/QA-QU_05-F9/2020 | EPI_ISL_1712864 |
| hCoV-19/Qatar/QA-QU_05-G2/2020 | EPI_ISL_1712867 |
| hCoV-19/Qatar/QA-QU_05-H6/2020 | EPI_ISL_1712878 |
| hCoV-19/Qatar/QA-QU_05-H7/2020 | EPI_ISL_1712879 |
| hCoV-19/Qatar/QA-QU_05-I7/2020 | EPI_ISL_1712887 |
| hCoV-19/Qatar/QA-QU_05-J2/2020 | EPI_ISL_1712889 |
| hCoV-19/Qatar/QA-QU_05-J3/2020 | EPI_ISL_1712890 |
| hCoV-19/Qatar/QA-WCMQ_FD18216518/2020 | EPI_ISL_1714663 |
| hCoV-19/Qatar/QA-WCMQ_FD18222868/2020 | EPI_ISL_1714666 |
| hCoV-19/Qatar/QA-WCMQ_FD18222877/2020 | EPI_ISL_1714667 |

| | |
|---|---|
| hCoV-19/Qatar/QA-QU_05-B4/2020 | EPI_ISL_1712837 |
| hCoV-19/Qatar/QA-QU_05-C7/2020 | EPI_ISL_1712846 |
| hCoV-19/Qatar/QA-QU_05-D9/2020 | EPI_ISL_1712851 |
| hCoV-19/Qatar/QA-QU_05-E10/2020 | EPI_ISL_1712853 |
| hCoV-19/Qatar/QA-WCMQ_FD18216498/2020 | EPI_ISL_1714662 |
| hCoV-19/Qatar/QA-WCMQ_FD18216524/2020 | EPI_ISL_1714664 |
| hCoV-19/Qatar/QA-WCMQ_FD18364037/2020 | EPI_ISL_1714743 |
| hCoV-19/Qatar/QA-WCMQ_FD18366226/2020 | EPI_ISL_1714755 |
| hCoV-19/Qatar/QA-QU_05-B8/2020 | EPI_ISL_1712841 |
| hCoV-19/Qatar/QA-QU_05-F7/2020 | EPI_ISL_1712862 |
| hCoV-19/Qatar/QA-QU_05-F8/2020 | EPI_ISL_1712863 |
| hCoV-19/Qatar/QA-WCMQ_FD15674720/2020 | EPI_ISL_1714156 |
| hCoV-19/Qatar/QA-WCMQ_FD18364075/2020 | EPI_ISL_1714744 |
| hCoV-19/Qatar/QA-QU_04-E8/2020 | EPI_ISL_1712813 |
| hCoV-19/Qatar/QA-QU_05-B5/2020 | EPI_ISL_1712838 |
| hCoV-19/Qatar/QA-QU_05-B6/2020 | EPI_ISL_1712839 |
| hCoV-19/Qatar/QA-QU_05-B7/2020 | EPI_ISL_1712840 |
| hCoV-19/Qatar/QA-QU_05-E3/2020 | EPI_ISL_1712855 |
| hCoV-19/Qatar/QA-WCMQ_FD15650635/2020 | EPI_ISL_1714132 |
| hCoV-19/Qatar/QA-WCMQ_FD15651331/2020 | EPI_ISL_1714135 |
| hCoV-19/Qatar/QA-WCMQ_FD15651333/2020 | EPI_ISL_1714136 |
| hCoV-19/Qatar/QA-WCMQ_FD18215300/2020 | EPI_ISL_1714653 |
| hCoV-19/Qatar/QA-WCMQ_FD18215301/2020 | EPI_ISL_1714654 |
| hCoV-19/Qatar/QA-WCMQ_FD18215304/2020 | EPI_ISL_1714655 |
| hCoV-19/Qatar/QA-WCMQ_FD18215306/2020 | EPI_ISL_1714656 |
| hCoV-19/Qatar/QA-QU_04-A5/2020 | EPI_ISL_1712795 |
| hCoV-19/Qatar/QA-QU_04-A6/2020 | EPI_ISL_1712796 |
| hCoV-19/Qatar/QA-QU_04-J8/2020 | EPI_ISL_1712830 |
| hCoV-19/Qatar/QA-QU_05-G8/2020 | EPI_ISL_1712872 |
| hCoV-19/Qatar/QA-QU_05-I8/2020 | EPI_ISL_1712888 |
| hCoV-19/Qatar/QA-WCMQ_FD15651569/2020 | EPI_ISL_1714139 |
| hCoV-19/Qatar/QA-WCMQ_FD15671484/2020 | EPI_ISL_1714143 |
| hCoV-19/Qatar/QA-WCMQ_FD18193903/2020 | EPI_ISL_1714630 |
| hCoV-19/Qatar/QA-WCMQ_FD18215266/2020 | EPI_ISL_1714650 |
| hCoV-19/Qatar/QA-WCMQ_FD18215269/2020 | EPI_ISL_1714651 |
| hCoV-19/Qatar/QA-QU_05-A1/2020 | EPI_ISL_1712831 |
| hCoV-19/Qatar/QA-QU_05-C6/2020 | EPI_ISL_1712845 |
| hCoV-19/Qatar/QA-QU_05-D2/2020 | EPI_ISL_1712849 |
| hCoV-19/Qatar/QA-QU_05-D4/2020 | EPI_ISL_1712850 |
| hCoV-19/Qatar/QA-QU_05-E1/2020 | EPI_ISL_1712852 |
| hCoV-19/Qatar/QA-QU_05-G10/2020 | EPI_ISL_1712866 |
| hCoV-19/Qatar/QA-QU_05-G3/2020 | EPI_ISL_1712868 |
| hCoV-19/Qatar/QA-QU_05-G4/2020 | EPI_ISL_1712869 |
| hCoV-19/Qatar/QA-QU_05-G7/2020 | EPI_ISL_1712871 |
| hCoV-19/Qatar/QA-QU_05-H1/2020 | EPI_ISL_1712873 |
| hCoV-19/Qatar/QA-QU_05-H2/2020 | EPI_ISL_1712875 |
| hCoV-19/Qatar/QA-QU_05-H3/2020 | EPI_ISL_1712876 |
| hCoV-19/Qatar/QA-QU_05-H8/2020 | EPI_ISL_1712880 |
| hCoV-19/Qatar/QA-QU_05-I2/2020 | EPI_ISL_1712882 |
| hCoV-19/Qatar/QA-QU_05-I3/2020 | EPI_ISL_1712883 |

| | |
|---|---|
| hCoV-19/Qatar/QA-QU_05-J5/2020 | EPI_ISL_1712892 |
| hCoV-19/Qatar/QA-WCMQ_FD15648572/2020 | EPI_ISL_1714126 |
| hCoV-19/Qatar/QA-WCMQ_FD15653230/2020 | EPI_ISL_1714141 |
| hCoV-19/Qatar/QA-WCMQ_FD15653243/2020 | EPI_ISL_1714142 |
| hCoV-19/Qatar/QA-WCMQ_FD15672876/2020 | EPI_ISL_1714145 |
| hCoV-19/Qatar/QA-WCMQ_FD15672879/2020 | EPI_ISL_1714146 |
| hCoV-19/Qatar/QA-WCMQ_FD15672880/2020 | EPI_ISL_1714147 |
| hCoV-19/Qatar/QA-WCMQ_FD15672882/2020 | EPI_ISL_1714148 |
| hCoV-19/Qatar/QA-WCMQ_FD15672898/2020 | EPI_ISL_1714150 |
| hCoV-19/Qatar/QA-WCMQ_FD18193856/2020 | EPI_ISL_1714584 |
| hCoV-19/Qatar/QA-QU_04-A8/2020 | EPI_ISL_1712797 |
| hCoV-19/Qatar/QA-QU_04-A9/2020 | EPI_ISL_1712798 |
| hCoV-19/Qatar/QA-QU_04-E1/2020 | EPI_ISL_1712809 |
| hCoV-19/Qatar/QA-QU_04-E2/2020 | EPI_ISL_1712811 |
| hCoV-19/Qatar/QA-QU_04-E3/2020 | EPI_ISL_1712812 |
| hCoV-19/Qatar/QA-QU_04-E9/2020 | EPI_ISL_1712814 |
| hCoV-19/Qatar/QA-QU_04-H1/2020 | EPI_ISL_1712822 |
| hCoV-19/Qatar/QA-QU_04-H2/2020 | EPI_ISL_1712824 |
| hCoV-19/Qatar/QA-QU_04-H4/2020 | EPI_ISL_1712825 |
| hCoV-19/Qatar/QA-QU_04-H6/2020 | EPI_ISL_1712826 |
| hCoV-19/Qatar/QA-QU_04-H7/2020 | EPI_ISL_1712827 |
| hCoV-19/Qatar/QA-QU_04-H8/2020 | EPI_ISL_1712828 |
| hCoV-19/Qatar/QA-QU_04-I6/2020 | EPI_ISL_1712829 |
| hCoV-19/Qatar/QA-QU_05-B10/2020 | EPI_ISL_1712835 |
| hCoV-19/Qatar/QA-QU_05-B2/2020 | EPI_ISL_1712836 |
| hCoV-19/Qatar/QA-QU_05-C2/2020 | EPI_ISL_1712843 |
| hCoV-19/Qatar/QA-QU_05-C5/2020 | EPI_ISL_1712844 |
| hCoV-19/Qatar/QA-QU_05-E8/2020 | EPI_ISL_1712859 |
| hCoV-19/Qatar/QA-QU_05-F10/2020 | EPI_ISL_1712860 |
| hCoV-19/Qatar/QA-QU_05-H10/2020 | EPI_ISL_1712874 |
| hCoV-19/Qatar/QA-QU_05-H4/2020 | EPI_ISL_1712877 |
| hCoV-19/Qatar/QA-QU_05-I6/2020 | EPI_ISL_1712886 |
| hCoV-19/Qatar/QA-QU_05-J6/2020 | EPI_ISL_1712893 |
| hCoV-19/Qatar/QA-QU_05-J7/2020 | EPI_ISL_1712894 |
| hCoV-19/Qatar/QA-WCMQ_FD15645971/2020 | EPI_ISL_1714125 |
| hCoV-19/Qatar/QA-WCMQ_FD15648575/2020 | EPI_ISL_1714127 |
| hCoV-19/Qatar/QA-WCMQ_FD15649701/2020 | EPI_ISL_1714128 |
| hCoV-19/Qatar/QA-WCMQ_FD15649738/2020 | EPI_ISL_1714130 |
| hCoV-19/Qatar/QA-WCMQ_FD15672845/2020 | EPI_ISL_1714144 |
| hCoV-19/Qatar/QA-WCMQ_FD15672883/2020 | EPI_ISL_1714149 |
| hCoV-19/Qatar/QA-WCMQ_FD15673744/2020 | EPI_ISL_1714152 |
| hCoV-19/Qatar/QA-WCMQ_FD15674674/2020 | EPI_ISL_1714153 |
| hCoV-19/Qatar/QA-WCMQ_FD15674698/2020 | EPI_ISL_1714154 |
| hCoV-19/Qatar/QA-WCMQ_FD15674717/2020 | EPI_ISL_1714155 |
| hCoV-19/Qatar/QA-WCMQ_FD15674928/2020 | EPI_ISL_1714157 |
| hCoV-19/Qatar/QA-WCMQ_FD15674947/2020 | EPI_ISL_1714158 |
| hCoV-19/Qatar/QA-WCMQ_FD15674989/2020 | EPI_ISL_1714159 |
| hCoV-19/Qatar/QA-WCMQ_FD15676536/2020 | EPI_ISL_1714161 |
| hCoV-19/Qatar/QA-WCMQ_FD18193822/2020 | EPI_ISL_1714550 |
| hCoV-19/Qatar/QA-WCMQ_FD18193910/2020 | EPI_ISL_1714637 |

| | |
|---|---|
| hCoV-19/Qatar/QA-WCMQ_FD18214894/2020 | EPI_ISL_1714645 |
| hCoV-19/Qatar/QA-WCMQ_FD18214900/2020 | EPI_ISL_1714646 |
| hCoV-19/Qatar/QA-WCMQ_FD18214947/2020 | EPI_ISL_1714647 |
| hCoV-19/Qatar/QA-WCMQ_FD18215283/2020 | EPI_ISL_1714652 |
| hCoV-19/Qatar/QA-WCMQ_FD18215309/2020 | EPI_ISL_1714657 |
| hCoV-19/Qatar/QA-WCMQ_FD18215325/2020 | EPI_ISL_1714659 |
| hCoV-19/Qatar/QA-WCMQ_FD18215529/2020 | EPI_ISL_1714661 |
| hCoV-19/Qatar/QA-QU_04-D1/2020 | EPI_ISL_1712804 |
| hCoV-19/Qatar/QA-QU_04-D2/2020 | EPI_ISL_1712806 |
| hCoV-19/Qatar/QA-QU_04-D3/2020 | EPI_ISL_1712807 |
| hCoV-19/Qatar/QA-QU_04-D4/2020 | EPI_ISL_1712808 |
| hCoV-19/Qatar/QA-QU_04-E10/2020 | EPI_ISL_1712810 |
| hCoV-19/Qatar/QA-QU_04-F3/2020 | EPI_ISL_1712815 |
| hCoV-19/Qatar/QA-QU_04-F4/2020 | EPI_ISL_1712816 |
| hCoV-19/Qatar/QA-QU_04-F6/2020 | EPI_ISL_1712817 |
| hCoV-19/Qatar/QA-QU_04-F7/2020 | EPI_ISL_1712818 |
| hCoV-19/Qatar/QA-QU_04-F8/2020 | EPI_ISL_1712819 |
| hCoV-19/Qatar/QA-QU_04-G1/2020 | EPI_ISL_1712820 |
| hCoV-19/Qatar/QA-QU_04-G9/2020 | EPI_ISL_1712821 |
| hCoV-19/Qatar/QA-QU_05-C10/2020 | EPI_ISL_1712842 |
| hCoV-19/Qatar/QA-QU_05-C9/2020 | EPI_ISL_1712848 |
| hCoV-19/Qatar/QA-QU_05-I4/2020 | EPI_ISL_1712884 |
| hCoV-19/Qatar/QA-WCMQ_FD15649726/2020 | EPI_ISL_1714129 |
| hCoV-19/Qatar/QA-WCMQ_FD15650061/2020 | EPI_ISL_1714131 |
| hCoV-19/Qatar/QA-WCMQ_FD15673719/2020 | EPI_ISL_1714151 |
| hCoV-19/Qatar/QA-WCMQ_FD15675017/2020 | EPI_ISL_1714160 |
| hCoV-19/Qatar/QA-WCMQ_FD18193847/2020 | EPI_ISL_1714575 |
| hCoV-19/Qatar/QA-WCMQ_FD18193878/2020 | EPI_ISL_1714605 |
| hCoV-19/Qatar/QA-WCMQ_FD18193899/2020 | EPI_ISL_1714626 |
| hCoV-19/Qatar/QA-WCMQ_FD18193914/2020 | EPI_ISL_1714641 |
| hCoV-19/Qatar/QA-WCMQ_FD18215505/2020 | EPI_ISL_1714660 |
| hCoV-19/Qatar/QA-WCMQ_FD18363620/2020 | EPI_ISL_1714741 |
| hCoV-19/Qatar/QA-WCMQ_FD18363627/2020 | EPI_ISL_1714742 |
| hCoV-19/Qatar/QA-WCMQ_FD18365446/2020 | EPI_ISL_1714750 |
| hCoV-19/Qatar/QA-WCMQ_FD18365465/2020 | EPI_ISL_1714751 |
| hCoV-19/Qatar/QA-WCMQ_FD18365515/2020 | EPI_ISL_1714752 |
| hCoV-19/Qatar/QA-WCMQ_FD18365535/2020 | EPI_ISL_1714753 |
| hCoV-19/Qatar/QA-WCMQ_FD18365537/2020 | EPI_ISL_1714754 |
| hCoV-19/Qatar/QA-QU_05-C8/2020 | EPI_ISL_1712847 |
| hCoV-19/Qatar/QA-QU_05-E6/2020 | EPI_ISL_1712857 |
| hCoV-19/Qatar/QA-QU_05-E7/2020 | EPI_ISL_1712858 |
| hCoV-19/Qatar/QA-WCMQ_FD15652594/2020 | EPI_ISL_1714140 |
| hCoV-19/Qatar/QA-WCMQ_FD18193826/2020 | EPI_ISL_1714554 |
| hCoV-19/Qatar/QA-WCMQ_FD18193854/2020 | EPI_ISL_1714582 |
| hCoV-19/Qatar/QA-WCMQ_FD18193855/2020 | EPI_ISL_1714583 |
| hCoV-19/Qatar/QA-WCMQ_FD18193859/2020 | EPI_ISL_1714586 |
| hCoV-19/Qatar/QA-WCMQ_FD18193900/2020 | EPI_ISL_1714627 |
| hCoV-19/Qatar/QA-WCMQ_FD18193909/2020 | EPI_ISL_1714636 |
| hCoV-19/Qatar/QA-WCMQ_FD18193912/2020 | EPI_ISL_1714639 |
| hCoV-19/Qatar/QA-QU_05-A3/2020 | EPI_ISL_1712832 |

| | |
|---|---|
| hCoV-19/Qatar/QA-QU_05-B1/2020 | EPI_ISL_1712834 |
| hCoV-19/Qatar/QA-QU_05-F6/2020 | EPI_ISL_1712861 |
| hCoV-19/Qatar/QA-QU_05-G5/2020 | EPI_ISL_1712870 |
| hCoV-19/Qatar/QA-QU_05-I5/2020 | EPI_ISL_1712885 |
| hCoV-19/Qatar/QA-WCMQ_FD18160907/2020 | EPI_ISL_1714362 |
| hCoV-19/Qatar/QA-WCMQ_FD18193823/2020 | EPI_ISL_1714551 |
| hCoV-19/Qatar/QA-WCMQ_FD18193824/2020 | EPI_ISL_1714552 |
| hCoV-19/Qatar/QA-WCMQ_FD18193827/2020 | EPI_ISL_1714555 |
| hCoV-19/Qatar/QA-WCMQ_FD18193828/2020 | EPI_ISL_1714556 |
| hCoV-19/Qatar/QA-WCMQ_FD18193829/2020 | EPI_ISL_1714557 |
| hCoV-19/Qatar/QA-WCMQ_FD18193830/2020 | EPI_ISL_1714558 |
| hCoV-19/Qatar/QA-WCMQ_FD18193831/2020 | EPI_ISL_1714559 |
| hCoV-19/Qatar/QA-WCMQ_FD18193832/2020 | EPI_ISL_1714560 |
| hCoV-19/Qatar/QA-WCMQ_FD18193833/2020 | EPI_ISL_1714561 |
| hCoV-19/Qatar/QA-WCMQ_FD18193836/2020 | EPI_ISL_1714564 |
| hCoV-19/Qatar/QA-WCMQ_FD18193871/2020 | EPI_ISL_1714598 |
| hCoV-19/Qatar/QA-WCMQ_FD18193825/2020 | EPI_ISL_1714553 |
| hCoV-19/Qatar/QA-WCMQ_FD18193860/2020 | EPI_ISL_1714587 |
| hCoV-19/Qatar/QA-WCMQ_FD18193868/2020 | EPI_ISL_1714595 |
| hCoV-19/Qatar/QA-WCMQ_FD18193885/2020 | EPI_ISL_1714612 |
| hCoV-19/Qatar/QA-WCMQ_FD18193886/2020 | EPI_ISL_1714613 |
| hCoV-19/Qatar/QA-WCMQ_FD18193917/2020 | EPI_ISL_1714644 |
| hCoV-19/Qatar/QA-WCMQ_FD18193861/2020 | EPI_ISL_1714588 |
| hCoV-19/Qatar/QA-WCMQ_FD18193863/2020 | EPI_ISL_1714590 |
| hCoV-19/Qatar/QA-WCMQ_FD18193879/2020 | EPI_ISL_1714606 |
| hCoV-19/Qatar/QA-WCMQ_FD18193880/2020 | EPI_ISL_1714607 |
| hCoV-19/Qatar/QA-WCMQ_FD18193888/2020 | EPI_ISL_1714615 |
| hCoV-19/Qatar/QA-WCMQ_FD18193906/2020 | EPI_ISL_1714633 |
| hCoV-19/Qatar/QA-WCMQ_FD15540700/2020 | EPI_ISL_1714057 |
| hCoV-19/Qatar/QA-WCMQ_FD15540728/2020 | EPI_ISL_1714069 |
| hCoV-19/Qatar/QA-WCMQ_FD17059325/2020 | EPI_ISL_1714178 |
| hCoV-19/Qatar/QA-WCMQ_FD18193838/2020 | EPI_ISL_1714566 |
| hCoV-19/Qatar/QA-WCMQ_FD18193846/2020 | EPI_ISL_1714574 |
| hCoV-19/Qatar/QA-WCMQ_FD18193892/2020 | EPI_ISL_1714619 |
| hCoV-19/Qatar/QA-WCMQ_FD18193893/2020 | EPI_ISL_1714620 |
| hCoV-19/Qatar/QA-WCMQ_FD18193894/2020 | EPI_ISL_1714621 |
| hCoV-19/Qatar/QA-WCMQ_FD18193895/2020 | EPI_ISL_1714622 |
| hCoV-19/Qatar/QA-WCMQ_FD18193896/2020 | EPI_ISL_1714623 |
| hCoV-19/Qatar/QA-WCMQ_FD18193897/2020 | EPI_ISL_1714624 |
| hCoV-19/Qatar/QA-WCMQ_FD18193905/2020 | EPI_ISL_1714632 |
| hCoV-19/Qatar/QA-WCMQ_FD18193840/2020 | EPI_ISL_1714568 |
| hCoV-19/Qatar/QA-WCMQ_FD18193841/2020 | EPI_ISL_1714569 |
| hCoV-19/Qatar/QA-WCMQ_FD18193842/2020 | EPI_ISL_1714570 |
| hCoV-19/Qatar/QA-WCMQ_FD18193852/2020 | EPI_ISL_1714580 |
| hCoV-19/Qatar/QA-WCMQ_FD18193857/2020 | EPI_ISL_1714585 |
| hCoV-19/Qatar/QA-WCMQ_FD18193862/2020 | EPI_ISL_1714589 |
| hCoV-19/Qatar/QA-WCMQ_FD18193865/2020 | EPI_ISL_1714592 |
| hCoV-19/Qatar/QA-WCMQ_FD18193866/2020 | EPI_ISL_1714593 |
| hCoV-19/Qatar/QA-WCMQ_FD18193875/2020 | EPI_ISL_1714602 |
| hCoV-19/Qatar/QA-WCMQ_FD18193877/2020 | EPI_ISL_1714604 |

| | |
|---|---|
| hCoV-19/Qatar/QA-WCMQ_FD18193898/2020 | EPI_ISL_1714625 |
| hCoV-19/Qatar/QA-WCMQ_FD18193904/2020 | EPI_ISL_1714631 |
| hCoV-19/Qatar/QA-WCMQ_FD18193913/2020 | EPI_ISL_1714640 |
| hCoV-19/Qatar/QA-WCMQ_FD17059315/2020 | EPI_ISL_1714172 |
| hCoV-19/Qatar/QA-WCMQ_FD18193834/2020 | EPI_ISL_1714562 |
| hCoV-19/Qatar/QA-WCMQ_FD18193837/2020 | EPI_ISL_1714565 |
| hCoV-19/Qatar/QA-WCMQ_FD18193843/2020 | EPI_ISL_1714571 |
| hCoV-19/Qatar/QA-WCMQ_FD18193850/2020 | EPI_ISL_1714578 |
| hCoV-19/Qatar/QA-WCMQ_FD18193853/2020 | EPI_ISL_1714581 |
| hCoV-19/Qatar/QA-WCMQ_FD18193869/2020 | EPI_ISL_1714596 |
| hCoV-19/Qatar/QA-WCMQ_FD18193881/2020 | EPI_ISL_1714608 |
| hCoV-19/Qatar/QA-WCMQ_FD18193887/2020 | EPI_ISL_1714614 |
| hCoV-19/Qatar/QA-WCMQ_FD18193889/2020 | EPI_ISL_1714616 |
| hCoV-19/Qatar/QA-WCMQ_FD18193890/2020 | EPI_ISL_1714617 |
| hCoV-19/Qatar/QA-WCMQ_FD18193891/2020 | EPI_ISL_1714618 |
| hCoV-19/Qatar/QA-WCMQ_FD18193915/2020 | EPI_ISL_1714642 |
| hCoV-19/Qatar/QA-WCMQ_FD18193916/2020 | EPI_ISL_1714643 |
| hCoV-19/Qatar/QA-WCMQ_FD15540731/2020 | EPI_ISL_1714071 |
| hCoV-19/Qatar/QA-WCMQ_FD18160881/2020 | EPI_ISL_1714355 |
| hCoV-19/Qatar/QA-WCMQ_FD18193839/2020 | EPI_ISL_1714567 |
| hCoV-19/Qatar/QA-WCMQ_FD18193864/2020 | EPI_ISL_1714591 |
| hCoV-19/Qatar/QA-WCMQ_FD18193882/2020 | EPI_ISL_1714609 |
| hCoV-19/Qatar/QA-WCMQ_FD18193901/2020 | EPI_ISL_1714628 |
| hCoV-19/Qatar/QA-QU_07-1-C6/2020 | EPI_ISL_1712924 |
| hCoV-19/Qatar/QA-QU_07-1-D9/2020 | EPI_ISL_1712939 |
| hCoV-19/Qatar/QA-QU_07-1-F5/2020 | EPI_ISL_1712949 |
| hCoV-19/Qatar/QA-QU_07-2-A4/2020 | EPI_ISL_1712980 |
| hCoV-19/Qatar/QA-QU_07-2-B4/2020 | EPI_ISL_1712989 |
| hCoV-19/Qatar/QA-QU_07-2-C1/2020 | EPI_ISL_1712995 |
| hCoV-19/Qatar/QA-QU_07-2-C4/2020 | EPI_ISL_1712999 |
| hCoV-19/Qatar/QA-QU_07-2-D2/2020 | EPI_ISL_1713005 |
| hCoV-19/Qatar/QA-QU_07-2-D4/2020 | EPI_ISL_1713007 |
| hCoV-19/Qatar/QA-QU_07-2-E12/2020 | EPI_ISL_1713014 |
| hCoV-19/Qatar/QA-QU_07-2-F11/2020 | EPI_ISL_1713022 |
| hCoV-19/Qatar/QA-WCMQ_FD17462447/2020 | EPI_ISL_1714266 |
| hCoV-19/Qatar/QA-WCMQ_FD18160880/2020 | EPI_ISL_1714354 |
| hCoV-19/Qatar/QA-WCMQ_FD18160897/2020 | EPI_ISL_1714361 |
| hCoV-19/Qatar/QA-WCMQ_FD18193851/2020 | EPI_ISL_1714579 |
| hCoV-19/Qatar/QA-WCMQ_FD18193874/2020 | EPI_ISL_1714601 |
| hCoV-19/Qatar/QA-WCMQ_FD18193876/2020 | EPI_ISL_1714603 |
| hCoV-19/Qatar/QA-WCMQ_FD18193902/2020 | EPI_ISL_1714629 |
| hCoV-19/Qatar/QA-WCMQ_FD18193911/2020 | EPI_ISL_1714638 |
| hCoV-19/Qatar/QA-QU_07-1-D1/2020 | EPI_ISL_1712928 |
| hCoV-19/Qatar/QA-QU_07-1-E12/2020 | EPI_ISL_1712943 |
| hCoV-19/Qatar/QA-QU_07-1-F1/2020 | EPI_ISL_1712946 |
| hCoV-19/Qatar/QA-QU_07-1-H6/2020 | EPI_ISL_1712972 |
| hCoV-19/Qatar/QA-QU_07-2-A1/2020 | EPI_ISL_1712975 |
| hCoV-19/Qatar/QA-QU_07-2-B11/2020 | EPI_ISL_1712985 |
| hCoV-19/Qatar/QA-QU_07-2-B6/2020 | EPI_ISL_1712991 |
| hCoV-19/Qatar/QA-QU_07-2-C7/2020 | EPI_ISL_1713001 |

| | |
|---|---|
| hCoV-19/Qatar/QA-QU_07-2-D7/2020 | EPI_ISL_1713009 |
| hCoV-19/Qatar/QA-WCMQ_FD17059314/2020 | EPI_ISL_1714171 |
| hCoV-19/Qatar/QA-WCMQ_FD17462364/2020 | EPI_ISL_1714190 |
| hCoV-19/Qatar/QA-WCMQ_FD17462366/2020 | EPI_ISL_1714192 |
| hCoV-19/Qatar/QA-WCMQ_FD17462372/2020 | EPI_ISL_1714197 |
| hCoV-19/Qatar/QA-WCMQ_FD17462408/2020 | EPI_ISL_1714230 |
| hCoV-19/Qatar/QA-WCMQ_FD17462432/2020 | EPI_ISL_1714251 |
| hCoV-19/Qatar/QA-WCMQ_FD17462450/2020 | EPI_ISL_1714269 |
| hCoV-19/Qatar/QA-QU_07-1-B2/2020 | EPI_ISL_1712910 |
| hCoV-19/Qatar/QA-QU_07-1-B3/2020 | EPI_ISL_1712911 |
| hCoV-19/Qatar/QA-QU_07-1-B8/2020 | EPI_ISL_1712916 |
| hCoV-19/Qatar/QA-QU_07-1-C1/2020 | EPI_ISL_1712918 |
| hCoV-19/Qatar/QA-QU_07-1-D10/2020 | EPI_ISL_1712929 |
| hCoV-19/Qatar/QA-QU_07-2-A2/2020 | EPI_ISL_1712978 |
| hCoV-19/Qatar/QA-QU_07-2-D6/2020 | EPI_ISL_1713008 |
| hCoV-19/Qatar/QA-QU_07-2-E7/2020 | EPI_ISL_1713019 |
| hCoV-19/Qatar/QA-WCMQ_FD15540730/2020 | EPI_ISL_1714070 |
| hCoV-19/Qatar/QA-WCMQ_FD17462362/2020 | EPI_ISL_1714188 |
| hCoV-19/Qatar/QA-WCMQ_FD17462386/2020 | EPI_ISL_1714211 |
| hCoV-19/Qatar/QA-WCMQ_FD17462394/2020 | EPI_ISL_1714217 |
| hCoV-19/Qatar/QA-WCMQ_FD17462401/2020 | EPI_ISL_1714223 |
| hCoV-19/Qatar/QA-WCMQ_FD17462456/2020 | EPI_ISL_1714274 |
| hCoV-19/Qatar/QA-QU_07-1-F6/2020 | EPI_ISL_1712950 |
| hCoV-19/Qatar/QA-QU_07-2-A3/2020 | EPI_ISL_1712979 |
| hCoV-19/Qatar/QA-QU_07-2-B1/2020 | EPI_ISL_1712983 |
| hCoV-19/Qatar/QA-QU_07-2-B12/2020 | EPI_ISL_1712986 |
| hCoV-19/Qatar/QA-QU_07-2-B7/2020 | EPI_ISL_1712992 |
| hCoV-19/Qatar/QA-QU_07-2-E6/2020 | EPI_ISL_1713018 |
| hCoV-19/Qatar/QA-QU_07-2-F6/2020 | EPI_ISL_1713028 |
| hCoV-19/Qatar/QA-QU_07-2-G5/2020 | EPI_ISL_1713032 |
| hCoV-19/Qatar/QA-QU_07-2-G8/2020 | EPI_ISL_1713034 |
| hCoV-19/Qatar/QA-QU_07-2-H6/2020 | EPI_ISL_1713038 |
| hCoV-19/Qatar/QA-WCMQ_FD15540588/2020 | EPI_ISL_1714050 |
| hCoV-19/Qatar/QA-WCMQ_FD17059318/2020 | EPI_ISL_1714175 |
| hCoV-19/Qatar/QA-WCMQ_FD17462393/2020 | EPI_ISL_1714216 |
| hCoV-19/Qatar/QA-WCMQ_FD17462426/2020 | EPI_ISL_1714247 |
| hCoV-19/Qatar/QA-WCMQ_FD17462440/2020 | EPI_ISL_1714259 |
| hCoV-19/Qatar/QA-QU_07-1-B11/2020 | EPI_ISL_1712908 |
| hCoV-19/Qatar/QA-QU_07-1-D11/2020 | EPI_ISL_1712930 |
| hCoV-19/Qatar/QA-QU_07-1-H5/2020 | EPI_ISL_1712971 |
| hCoV-19/Qatar/QA-QU_07-2-C5/2020 | EPI_ISL_1713000 |
| hCoV-19/Qatar/QA-QU_07-2-D8/2020 | EPI_ISL_1713010 |
| hCoV-19/Qatar/QA-QU_07-2-F5/2020 | EPI_ISL_1713027 |
| hCoV-19/Qatar/QA-QU_07-2-G7/2020 | EPI_ISL_1713033 |
| hCoV-19/Qatar/QA-WCMQ_FD17462395/2020 | EPI_ISL_1714218 |
| hCoV-19/Qatar/QA-WCMQ_FD17462425/2020 | EPI_ISL_1714246 |
| hCoV-19/Qatar/QA-WCMQ_FD17462433/2020 | EPI_ISL_1714252 |
| hCoV-19/Qatar/QA-QU_07-1-A1/2020 | EPI_ISL_1712896 |
| hCoV-19/Qatar/QA-QU_07-1-A9/2020 | EPI_ISL_1712906 |
| hCoV-19/Qatar/QA-QU_07-1-B1/2020 | EPI_ISL_1712907 |

| | |
|---|---|
| hCoV-19/Qatar/QA-QU_07-1-C2/2020 | EPI_ISL_1712921 |
| hCoV-19/Qatar/QA-QU_07-1-D2/2020 | EPI_ISL_1712932 |
| hCoV-19/Qatar/QA-QU_07-1-D4/2020 | EPI_ISL_1712934 |
| hCoV-19/Qatar/QA-QU_07-1-G10/2020 | EPI_ISL_1712955 |
| hCoV-19/Qatar/QA-QU_07-1-G6/2020 | EPI_ISL_1712962 |
| hCoV-19/Qatar/QA-QU_07-2-C3/2020 | EPI_ISL_1712998 |
| hCoV-19/Qatar/QA-QU_07-2-E2/2020 | EPI_ISL_1713015 |
| hCoV-19/Qatar/QA-QU_07-2-E8/2020 | EPI_ISL_1713020 |
| hCoV-19/Qatar/QA-QU_07-2-F2/2020 | EPI_ISL_1713024 |
| hCoV-19/Qatar/QA-QU_07-2-F7/2020 | EPI_ISL_1713029 |
| hCoV-19/Qatar/QA-QU_08-A4/2020 | EPI_ISL_1713042 |
| hCoV-19/Qatar/QA-QU_08-A5/2020 | EPI_ISL_1713043 |
| hCoV-19/Qatar/QA-QU_08-C4/2020 | EPI_ISL_1713051 |
| hCoV-19/Qatar/QA-WCMQ_FD15540687/2020 | EPI_ISL_1714051 |
| hCoV-19/Qatar/QA-WCMQ_FD17059317/2020 | EPI_ISL_1714174 |
| hCoV-19/Qatar/QA-WCMQ_FD17462402/2020 | EPI_ISL_1714224 |
| hCoV-19/Qatar/QA-WCMQ_FD17462403/2020 | EPI_ISL_1714225 |
| hCoV-19/Qatar/QA-WCMQ_FD17462411/2020 | EPI_ISL_1714233 |
| hCoV-19/Qatar/QA-WCMQ_FD17462415/2020 | EPI_ISL_1714236 |
| hCoV-19/Qatar/QA-WCMQ_FD17462431/2020 | EPI_ISL_1714250 |
| hCoV-19/Qatar/QA-QU_07-1-D12/2020 | EPI_ISL_1712931 |
| hCoV-19/Qatar/QA-QU_07-1-D3/2020 | EPI_ISL_1712933 |
| hCoV-19/Qatar/QA-QU_07-2-A6/2020 | EPI_ISL_1712981 |
| hCoV-19/Qatar/QA-QU_07-2-B3/2020 | EPI_ISL_1712988 |
| hCoV-19/Qatar/QA-QU_07-2-D3/2020 | EPI_ISL_1713006 |
| hCoV-19/Qatar/QA-QU_07-2-H10/2020 | EPI_ISL_1713035 |
| hCoV-19/Qatar/QA-WCMQ_FD17462439/2020 | EPI_ISL_1714258 |
| hCoV-19/Qatar/QA-WCMQ_FD17462441/2020 | EPI_ISL_1714260 |
| hCoV-19/Qatar/QA-QU_07-1-A5/2020 | EPI_ISL_1712902 |
| hCoV-19/Qatar/QA-QU_07-1-C5/2020 | EPI_ISL_1712923 |
| hCoV-19/Qatar/QA-QU_07-1-D5/2020 | EPI_ISL_1712935 |
| hCoV-19/Qatar/QA-QU_07-1-D6/2020 | EPI_ISL_1712936 |
| hCoV-19/Qatar/QA-QU_07-1-E1/2020 | EPI_ISL_1712940 |
| hCoV-19/Qatar/QA-QU_07-1-E10/2020 | EPI_ISL_1712941 |
| hCoV-19/Qatar/QA-QU_07-1-H9/2020 | EPI_ISL_1712974 |
| hCoV-19/Qatar/QA-QU_07-2-B8/2020 | EPI_ISL_1712993 |
| hCoV-19/Qatar/QA-QU_07-2-E1/2020 | EPI_ISL_1713012 |
| hCoV-19/Qatar/QA-QU_07-2-F8/2020 | EPI_ISL_1713030 |
| hCoV-19/Qatar/QA-QU_08-C2/2020 | EPI_ISL_1713049 |
| hCoV-19/Qatar/QA-QU_08-C3/2020 | EPI_ISL_1713050 |
| hCoV-19/Qatar/QA-QU_08-D1/2020 | EPI_ISL_1713053 |
| hCoV-19/Qatar/QA-QU_08-G1/2020 | EPI_ISL_1713064 |
| hCoV-19/Qatar/QA-WCMQ_FD17462368/2020 | EPI_ISL_1714194 |
| hCoV-19/Qatar/QA-WCMQ_FD17462436/2020 | EPI_ISL_1714255 |
| hCoV-19/Qatar/QA-QU_07-1-A3/2020 | EPI_ISL_1712900 |
| hCoV-19/Qatar/QA-QU_07-1-A4/2020 | EPI_ISL_1712901 |
| hCoV-19/Qatar/QA-QU_07-1-A6/2020 | EPI_ISL_1712903 |
| hCoV-19/Qatar/QA-QU_07-1-B7/2020 | EPI_ISL_1712915 |
| hCoV-19/Qatar/QA-QU_07-1-C9/2020 | EPI_ISL_1712927 |
| hCoV-19/Qatar/QA-QU_07-1-F3/2020 | EPI_ISL_1712947 |

| | |
|---|---|
| hCoV-19/Qatar/QA-QU_07-1-G5/2020 | EPI_ISL_1712961 |
| hCoV-19/Qatar/QA-QU_07-1-H1/2020 | EPI_ISL_1712965 |
| hCoV-19/Qatar/QA-QU_07-2-B2/2020 | EPI_ISL_1712987 |
| hCoV-19/Qatar/QA-QU_08-B1/2020 | EPI_ISL_1713044 |
| hCoV-19/Qatar/QA-QU_08-B3/2020 | EPI_ISL_1713046 |
| hCoV-19/Qatar/QA-QU_08-B4/2020 | EPI_ISL_1713047 |
| hCoV-19/Qatar/QA-QU_08-C5/2020 | EPI_ISL_1713052 |
| hCoV-19/Qatar/QA-QU_08-D5/2020 | EPI_ISL_1713054 |
| hCoV-19/Qatar/QA-QU_08-E1/2020 | EPI_ISL_1713055 |
| hCoV-19/Qatar/QA-QU_08-E2/2020 | EPI_ISL_1713056 |
| hCoV-19/Qatar/QA-QU_08-F2/2020 | EPI_ISL_1713060 |
| hCoV-19/Qatar/QA-QU_08-F3/2020 | EPI_ISL_1713061 |
| hCoV-19/Qatar/QA-QU_08-F5/2020 | EPI_ISL_1713063 |
| hCoV-19/Qatar/QA-QU_08-G4/2020 | EPI_ISL_1713067 |
| hCoV-19/Qatar/QA-QU_08-G5/2020 | EPI_ISL_1713068 |
| hCoV-19/Qatar/QA-WCMQ_FD17059306/2020 | EPI_ISL_1714163 |
| hCoV-19/Qatar/QA-WCMQ_FD17462419/2020 | EPI_ISL_1714240 |
| hCoV-19/Qatar/QA-WCMQ_FD17462429/2020 | EPI_ISL_1714249 |
| hCoV-19/Qatar/QA-WCMQ_FD17462438/2020 | EPI_ISL_1714257 |
| hCoV-19/Qatar/QA-QU_07-1-B5/2020 | EPI_ISL_1712913 |
| hCoV-19/Qatar/QA-QU_07-1-B9/2020 | EPI_ISL_1712917 |
| hCoV-19/Qatar/QA-QU_07-2-B5/2020 | EPI_ISL_1712990 |
| hCoV-19/Qatar/QA-QU_08-F4/2020 | EPI_ISL_1713062 |
| hCoV-19/Qatar/QA-WCMQ_FD17059312/2020 | EPI_ISL_1714169 |
| hCoV-19/Qatar/QA-WCMQ_FD17462374/2020 | EPI_ISL_1714199 |
| hCoV-19/Qatar/QA-WCMQ_FD17462452/2020 | EPI_ISL_1714271 |
| hCoV-19/Qatar/QA-QU_07-1-D8/2020 | EPI_ISL_1712938 |
| hCoV-19/Qatar/QA-WCMQ_FD17462442/2020 | EPI_ISL_1714261 |
| hCoV-19/Qatar/QA-WCMQ_FD17462444/2020 | EPI_ISL_1714263 |
| hCoV-19/Qatar/QA-QU_07-1-A11/2020 | EPI_ISL_1712897 |
| hCoV-19/Qatar/QA-QU_07-1-G9/2020 | EPI_ISL_1712964 |
| hCoV-19/Qatar/QA-QU_07-2-C11/2020 | EPI_ISL_1712996 |
| hCoV-19/Qatar/QA-QU_08-G2/2020 | EPI_ISL_1713065 |
| hCoV-19/Qatar/QA-QU_08-H2/2020 | EPI_ISL_1713069 |
| hCoV-19/Qatar/QA-QU_08-H4/2020 | EPI_ISL_1713071 |
| hCoV-19/Qatar/QA-WCMQ_FD15540740/2020 | EPI_ISL_1714075 |
| hCoV-19/Qatar/QA-WCMQ_FD17462376/2020 | EPI_ISL_1714201 |
| hCoV-19/Qatar/QA-WCMQ_FD17462405/2020 | EPI_ISL_1714227 |
| hCoV-19/Qatar/QA-QU_07-1-D7/2020 | EPI_ISL_1712937 |
| hCoV-19/Qatar/QA-QU_07-1-G1/2020 | EPI_ISL_1712954 |
| hCoV-19/Qatar/QA-QU_08-A1/2020 | EPI_ISL_1713040 |
| hCoV-19/Qatar/QA-QU_08-B2/2020 | EPI_ISL_1713045 |
| hCoV-19/Qatar/QA-QU_08-E4/2020 | EPI_ISL_1713057 |
| hCoV-19/Qatar/QA-QU_08-G3/2020 | EPI_ISL_1713066 |
| hCoV-19/Qatar/QA-QU_08-H3/2020 | EPI_ISL_1713070 |
| hCoV-19/Qatar/QA-WCMQ_FD15540690/2020 | EPI_ISL_1714052 |
| hCoV-19/Qatar/QA-WCMQ_FD17462361/2020 | EPI_ISL_1714187 |
| hCoV-19/Qatar/QA-WCMQ_FD17462363/2020 | EPI_ISL_1714189 |
| hCoV-19/Qatar/QA-WCMQ_FD17462379/2020 | EPI_ISL_1714204 |
| hCoV-19/Qatar/QA-WCMQ_FD17462381/2020 | EPI_ISL_1714206 |

| | |
|---|---|
| hCoV-19/Qatar/QA-WCMQ_FD17462413/2020 | EPI_ISL_1714234 |
| hCoV-19/Qatar/QA-WCMQ_FD17462414/2020 | EPI_ISL_1714235 |
| hCoV-19/Qatar/QA-WCMQ_FD17462417/2020 | EPI_ISL_1714238 |
| hCoV-19/Qatar/QA-WCMQ_FD17462455/2020 | EPI_ISL_1714273 |
| hCoV-19/Qatar/QA-QU_07-1-A8/2020 | EPI_ISL_1712905 |
| hCoV-19/Qatar/QA-QU_07-1-B4/2020 | EPI_ISL_1712912 |
| hCoV-19/Qatar/QA-QU_07-1-E11/2020 | EPI_ISL_1712942 |
| hCoV-19/Qatar/QA-QU_07-1-E9/2020 | EPI_ISL_1712945 |
| hCoV-19/Qatar/QA-QU_07-1-F9/2020 | EPI_ISL_1712953 |
| hCoV-19/Qatar/QA-QU_07-1-H3/2020 | EPI_ISL_1712969 |
| hCoV-19/Qatar/QA-QU_07-1-H4/2020 | EPI_ISL_1712970 |
| hCoV-19/Qatar/QA-QU_07-1-H8/2020 | EPI_ISL_1712973 |
| hCoV-19/Qatar/QA-QU_07-2-F3/2020 | EPI_ISL_1713025 |
| hCoV-19/Qatar/QA-QU_08-B5/2020 | EPI_ISL_1713048 |
| hCoV-19/Qatar/QA-QU_08-E5/2020 | EPI_ISL_1713058 |
| hCoV-19/Qatar/QA-WCMQ_FD17462383/2020 | EPI_ISL_1714208 |
| hCoV-19/Qatar/QA-WCMQ_FD17462384/2020 | EPI_ISL_1714209 |
| hCoV-19/Qatar/QA-WCMQ_FD17462399/2020 | EPI_ISL_1714221 |
| hCoV-19/Qatar/QA-WCMQ_FD17462446/2020 | EPI_ISL_1714265 |
| hCoV-19/Qatar/QA-WCMQ_FD17462448/2020 | EPI_ISL_1714267 |
| hCoV-19/Qatar/QA-QU_07-1-F8/2020 | EPI_ISL_1712952 |
| hCoV-19/Qatar/QA-QU_07-2-C12/2020 | EPI_ISL_1712997 |
| hCoV-19/Qatar/QA-QU_07-2-F9/2020 | EPI_ISL_1713031 |
| hCoV-19/Qatar/QA-QU_07-2-H3/2020 | EPI_ISL_1713036 |
| hCoV-19/Qatar/QA-QU_07-2-H9/2020 | EPI_ISL_1713039 |
| hCoV-19/Qatar/QA-QU_08-F1/2020 | EPI_ISL_1713059 |
| hCoV-19/Qatar/QA-WCMQ_FD17462390/2020 | EPI_ISL_1714214 |
| hCoV-19/Qatar/QA-WCMQ_FD17462423/2020 | EPI_ISL_1714244 |
| hCoV-19/Qatar/QA-WCMQ_FD17462424/2020 | EPI_ISL_1714245 |
| hCoV-19/Qatar/QA-WCMQ_FD17462434/2020 | EPI_ISL_1714253 |
| hCoV-19/Qatar/QA-QU_07-1-A12/2020 | EPI_ISL_1712898 |
| hCoV-19/Qatar/QA-QU_07-1-C11/2020 | EPI_ISL_1712920 |
| hCoV-19/Qatar/QA-QU_07-1-E2/2020 | EPI_ISL_1712944 |
| hCoV-19/Qatar/QA-QU_07-1-H10/2020 | EPI_ISL_1712966 |
| hCoV-19/Qatar/QA-QU_07-1-H12/2020 | EPI_ISL_1712967 |
| hCoV-19/Qatar/QA-QU_07-2-A11/2020 | EPI_ISL_1712977 |
| hCoV-19/Qatar/QA-QU_09-1-C5/2020 | EPI_ISL_1713087 |
| hCoV-19/Qatar/QA-WCMQ_FD15540732/2020 | EPI_ISL_1714072 |
| hCoV-19/Qatar/QA-WCMQ_FD15540733/2020 | EPI_ISL_1714073 |
| hCoV-19/Qatar/QA-WCMQ_FD17462367/2020 | EPI_ISL_1714193 |
| hCoV-19/Qatar/QA-WCMQ_FD17462418/2020 | EPI_ISL_1714239 |
| hCoV-19/Qatar/QA-QU_07-1-G8/2020 | EPI_ISL_1712963 |
| hCoV-19/Qatar/QA-QU_07-2-B10/2020 | EPI_ISL_1712984 |
| hCoV-19/Qatar/QA-QU_07-2-D10/2020 | EPI_ISL_1713002 |
| hCoV-19/Qatar/QA-QU_09-2-E6/2020 | EPI_ISL_1713132 |
| hCoV-19/Qatar/QA-WCMQ_FD17462392/2020 | EPI_ISL_1714215 |
| hCoV-19/Qatar/QA-WCMQ_FD17462398/2020 | EPI_ISL_1714220 |
| hCoV-19/Qatar/QA-WCMQ_FD17462435/2020 | EPI_ISL_1714254 |
| hCoV-19/Qatar/QA-QU_07-1-A7/2020 | EPI_ISL_1712904 |
| hCoV-19/Qatar/QA-QU_07-1-B12/2020 | EPI_ISL_1712909 |

| | |
|---|---|
| hCoV-19/Qatar/QA-QU_07-1-C8/2020 | EPI_ISL_1712926 |
| hCoV-19/Qatar/QA-QU_07-1-G2/2020 | EPI_ISL_1712958 |
| hCoV-19/Qatar/QA-QU_07-1-G3/2020 | EPI_ISL_1712959 |
| hCoV-19/Qatar/QA-QU_07-1-H2/2020 | EPI_ISL_1712968 |
| hCoV-19/Qatar/QA-QU_07-2-A10/2020 | EPI_ISL_1712976 |
| hCoV-19/Qatar/QA-QU_07-2-B9/2020 | EPI_ISL_1712994 |
| hCoV-19/Qatar/QA-QU_07-2-D12/2020 | EPI_ISL_1713004 |
| hCoV-19/Qatar/QA-QU_07-2-E3/2020 | EPI_ISL_1713016 |
| hCoV-19/Qatar/QA-QU_07-2-E9/2020 | EPI_ISL_1713021 |
| hCoV-19/Qatar/QA-QU_08-A2/2020 | EPI_ISL_1713041 |
| hCoV-19/Qatar/QA-WCMQ_FD17462369/2020 | EPI_ISL_1714195 |
| hCoV-19/Qatar/QA-WCMQ_FD17462373/2020 | EPI_ISL_1714198 |
| hCoV-19/Qatar/QA-WCMQ_FD17462375/2020 | EPI_ISL_1714200 |
| hCoV-19/Qatar/QA-WCMQ_FD17462380/2020 | EPI_ISL_1714205 |
| hCoV-19/Qatar/QA-WCMQ_FD17462409/2020 | EPI_ISL_1714231 |
| hCoV-19/Qatar/QA-WCMQ_FD17462420/2020 | EPI_ISL_1714241 |
| hCoV-19/Qatar/QA-WCMQ_FD17462422/2020 | EPI_ISL_1714243 |
| hCoV-19/Qatar/QA-WCMQ_FD18160923/2020 | EPI_ISL_1714365 |
| hCoV-19/Qatar/QA-QU_07-1-G4/2020 | EPI_ISL_1712960 |
| hCoV-19/Qatar/QA-QU_07-2-E10/2020 | EPI_ISL_1713013 |
| hCoV-19/Qatar/QA-QU_07-2-F4/2020 | EPI_ISL_1713026 |
| hCoV-19/Qatar/QA-WCMQ_FD17462421/2020 | EPI_ISL_1714242 |
| hCoV-19/Qatar/QA-WCMQ_FD17462445/2020 | EPI_ISL_1714264 |
| hCoV-19/Qatar/QA-QU_07-1-B6/2020 | EPI_ISL_1712914 |
| hCoV-19/Qatar/QA-QU_07-1-C10/2020 | EPI_ISL_1712919 |
| hCoV-19/Qatar/QA-QU_07-1-F4/2020 | EPI_ISL_1712948 |
| hCoV-19/Qatar/QA-QU_07-1-G12/2020 | EPI_ISL_1712957 |
| hCoV-19/Qatar/QA-QU_07-2-A9/2020 | EPI_ISL_1712982 |
| hCoV-19/Qatar/QA-WCMQ_FD17462406/2020 | EPI_ISL_1714228 |
| hCoV-19/Qatar/QA-WCMQ_FD17462410/2020 | EPI_ISL_1714232 |
| hCoV-19/Qatar/QA-WCMQ_FD17462449/2020 | EPI_ISL_1714268 |
| hCoV-19/Qatar/QA-WCMQ_FD17462454/2020 | EPI_ISL_1714272 |
| hCoV-19/Qatar/QA-QU_07-2-D11/2020 | EPI_ISL_1713003 |
| hCoV-19/Qatar/QA-WCMQ_FD17462371/2020 | EPI_ISL_1714196 |
| hCoV-19/Qatar/QA-WCMQ_FD17462382/2020 | EPI_ISL_1714207 |
| hCoV-19/Qatar/QA-WCMQ_FD17462389/2020 | EPI_ISL_1714213 |
| hCoV-19/Qatar/QA-WCMQ_FD17462416/2020 | EPI_ISL_1714237 |
| hCoV-19/Qatar/QA-QU_07-1-A2/2020 | EPI_ISL_1712899 |
| hCoV-19/Qatar/QA-QU_07-1-C3/2020 | EPI_ISL_1712922 |
| hCoV-19/Qatar/QA-QU_07-1-F7/2020 | EPI_ISL_1712951 |
| hCoV-19/Qatar/QA-QU_07-2-D9/2020 | EPI_ISL_1713011 |
| hCoV-19/Qatar/QA-QU_07-2-F12/2020 | EPI_ISL_1713023 |
| hCoV-19/Qatar/QA-WCMQ_FD17462365/2020 | EPI_ISL_1714191 |
| hCoV-19/Qatar/QA-WCMQ_FD17462437/2020 | EPI_ISL_1714256 |
| hCoV-19/Qatar/QA-WCMQ_FD17462451/2020 | EPI_ISL_1714270 |
| hCoV-19/Qatar/QA-QU_07-1-C7/2020 | EPI_ISL_1712925 |
| hCoV-19/Qatar/QA-QU_07-1-G11/2020 | EPI_ISL_1712956 |
| hCoV-19/Qatar/QA-QU_07-2-E4/2020 | EPI_ISL_1713017 |
| hCoV-19/Qatar/QA-QU_07-2-H4/2020 | EPI_ISL_1713037 |
| hCoV-19/Qatar/QA-WCMQ_FD17059316/2020 | EPI_ISL_1714173 |

| | |
|---|---|
| hCoV-19/Qatar/QA-WCMQ_FD17462387/2020 | EPI_ISL_1714212 |
| hCoV-19/Qatar/QA-QU_09-1-C6/2020 | EPI_ISL_1713088 |
| hCoV-19/Qatar/QA-WCMQ_FD17462404/2020 | EPI_ISL_1714226 |
| hCoV-19/Qatar/QA-QU_12-6-E11/2020 | EPI_ISL_1713292 |
| hCoV-19/Qatar/QA-WCMQ_FD17462397/2020 | EPI_ISL_1714219 |
| hCoV-19/Qatar/QA-QU_12-6-D1/2020 | EPI_ISL_1713290 |
| hCoV-19/Qatar/QA-QU_12-6-E4/2020 | EPI_ISL_1713294 |
| hCoV-19/Qatar/QA-WCMQ_FD17059321/2020 | EPI_ISL_1714177 |
| hCoV-19/Qatar/QA-WCMQ_FD17462400/2020 | EPI_ISL_1714222 |
| hCoV-19/Qatar/QA-WCMQ_FD17462407/2020 | EPI_ISL_1714229 |
| hCoV-19/Qatar/QA-QU_12-6-B2/2020 | EPI_ISL_1713289 |
| hCoV-19/Qatar/QA-QU_12-6-E1/2020 | EPI_ISL_1713291 |
| hCoV-19/Qatar/QA-QU_12-6-E12/2020 | EPI_ISL_1713293 |
| hCoV-19/Qatar/QA-QU_12-6-G10/2020 | EPI_ISL_1713295 |
| hCoV-19/Qatar/QA-WCMQ_FD17059309/2020 | EPI_ISL_1714166 |
| hCoV-19/Qatar/QA-WCMQ_FD17462377/2020 | EPI_ISL_1714202 |
| hCoV-19/Qatar/QA-WCMQ_FD17462378/2020 | EPI_ISL_1714203 |
| hCoV-19/Qatar/QA-WCMQ_FD17462428/2020 | EPI_ISL_1714248 |
| hCoV-19/Qatar/QA-WCMQ_FD17462443/2020 | EPI_ISL_1714262 |
| hCoV-19/Qatar/QA-QU_12-7-F5/2020 | EPI_ISL_1713316 |
| hCoV-19/Qatar/QA-QU_12-7-G8/2020 | EPI_ISL_1713320 |
| hCoV-19/Qatar/QA-WCMQ_FD17462385/2020 | EPI_ISL_1714210 |
| hCoV-19/Qatar/QA-WCMQ_FD18189985/2020 | EPI_ISL_1714464 |
| hCoV-19/Qatar/QA-WCMQ_FD18189988/2020 | EPI_ISL_1714467 |
| hCoV-19/Qatar/QA-WCMQ_FD18189996/2020 | EPI_ISL_1714475 |
| hCoV-19/Qatar/QA-WCMQ_FD18190008/2020 | EPI_ISL_1714487 |
| hCoV-19/Qatar/QA-WCMQ_FD18190020/2020 | EPI_ISL_1714498 |
| hCoV-19/Qatar/QA-WCMQ_FD18190029/2020 | EPI_ISL_1714507 |
| hCoV-19/Qatar/QA-WCMQ_FD18190068/2020 | EPI_ISL_1714542 |
| hCoV-19/Qatar/QA-WCMQ_FD18190072/2020 | EPI_ISL_1714546 |
| hCoV-19/Qatar/QA-QU_12-7-C9/2020 | EPI_ISL_1713305 |
| hCoV-19/Qatar/QA-QU_12-7-E11/2020 | EPI_ISL_1713311 |
| hCoV-19/Qatar/QA-QU_12-7-E7/2020 | EPI_ISL_1713314 |
| hCoV-19/Qatar/QA-QU_12-7-H1/2020 | EPI_ISL_1713321 |
| hCoV-19/Qatar/QA-QU_12-7-H12/2020 | EPI_ISL_1713322 |
| hCoV-19/Qatar/QA-QU_12-7-H9/2020 | EPI_ISL_1713326 |
| hCoV-19/Qatar/QA-WCMQ_FD17059308/2020 | EPI_ISL_1714165 |
| hCoV-19/Qatar/QA-WCMQ_FD17059311/2020 | EPI_ISL_1714168 |
| hCoV-19/Qatar/QA-WCMQ_FD18190005/2020 | EPI_ISL_1714484 |
| hCoV-19/Qatar/QA-WCMQ_FD18190010/2020 | EPI_ISL_1714489 |
| hCoV-19/Qatar/QA-WCMQ_FD18190032/2020 | EPI_ISL_1714510 |
| hCoV-19/Qatar/QA-WCMQ_FD18190040/2020 | EPI_ISL_1714518 |
| hCoV-19/Qatar/QA-WCMQ_FD18190047/2020 | EPI_ISL_1714523 |
| hCoV-19/Qatar/QA-WCMQ_FD18190054/2020 | EPI_ISL_1714529 |
| hCoV-19/Qatar/QA-WCMQ_FD18190064/2020 | EPI_ISL_1714538 |
| hCoV-19/Qatar/QA-WCMQ_FD18190073/2020 | EPI_ISL_1714547 |
| hCoV-19/Qatar/QA-QU_12-7-A11/2020 | EPI_ISL_1713297 |
| hCoV-19/Qatar/QA-QU_12-7-D5/2020 | EPI_ISL_1713307 |
| hCoV-19/Qatar/QA-QU_12-7-H6/2020 | EPI_ISL_1713324 |
| hCoV-19/Qatar/QA-WCMQ_FD18189986/2020 | EPI_ISL_1714465 |

| | |
|---|---|
| hCoV-19/Qatar/QA-WCMQ_FD18189993/2020 | EPI_ISL_1714472 |
| hCoV-19/Qatar/QA-WCMQ_FD18190009/2020 | EPI_ISL_1714488 |
| hCoV-19/Qatar/QA-WCMQ_FD18190066/2020 | EPI_ISL_1714540 |
| hCoV-19/Qatar/QA-QU_12-7-A10/2020 | EPI_ISL_1713296 |
| hCoV-19/Qatar/QA-QU_12-7-B11/2020 | EPI_ISL_1713301 |
| hCoV-19/Qatar/QA-QU_12-7-D7/2020 | EPI_ISL_1713308 |
| hCoV-19/Qatar/QA-QU_12-7-E3/2020 | EPI_ISL_1713312 |
| hCoV-19/Qatar/QA-QU_12-7-F3/2020 | EPI_ISL_1713315 |
| hCoV-19/Qatar/QA-QU_12-7-H4/2020 | EPI_ISL_1713323 |
| hCoV-19/Qatar/QA-WCMQ_FD17059310/2020 | EPI_ISL_1714167 |
| hCoV-19/Qatar/QA-WCMQ_FD18189994/2020 | EPI_ISL_1714473 |
| hCoV-19/Qatar/QA-WCMQ_FD18189995/2020 | EPI_ISL_1714474 |
| hCoV-19/Qatar/QA-WCMQ_FD18190007/2020 | EPI_ISL_1714486 |
| hCoV-19/Qatar/QA-WCMQ_FD18190017/2020 | EPI_ISL_1714495 |
| hCoV-19/Qatar/QA-WCMQ_FD18190058/2020 | EPI_ISL_1714533 |
| hCoV-19/Qatar/QA-WCMQ_FD18190065/2020 | EPI_ISL_1714539 |
| hCoV-19/Qatar/QA-WCMQ_FD18190069/2020 | EPI_ISL_1714543 |
| hCoV-19/Qatar/QA-QU_12-7-A5/2020 | EPI_ISL_1713299 |
| hCoV-19/Qatar/QA-QU_12-7-C10/2020 | EPI_ISL_1713304 |
| hCoV-19/Qatar/QA-QU_12-7-H7/2020 | EPI_ISL_1713325 |
| hCoV-19/Qatar/QA-WCMQ_FD18189989/2020 | EPI_ISL_1714468 |
| hCoV-19/Qatar/QA-WCMQ_FD18190002/2020 | EPI_ISL_1714481 |
| hCoV-19/Qatar/QA-WCMQ_FD18190003/2020 | EPI_ISL_1714482 |
| hCoV-19/Qatar/QA-WCMQ_FD18190034/2020 | EPI_ISL_1714512 |
| hCoV-19/Qatar/QA-WCMQ_FD18190038/2020 | EPI_ISL_1714516 |
| hCoV-19/Qatar/QA-WCMQ_FD18190052/2020 | EPI_ISL_1714527 |
| hCoV-19/Qatar/QA-WCMQ_FD18190055/2020 | EPI_ISL_1714530 |
| hCoV-19/Qatar/QA-WCMQ_FD18190056/2020 | EPI_ISL_1714531 |
| hCoV-19/Qatar/QA-WCMQ_FD18190057/2020 | EPI_ISL_1714532 |
| hCoV-19/Qatar/QA-QU_12-7-B1/2020 | EPI_ISL_1713300 |
| hCoV-19/Qatar/QA-WCMQ_FD17059320/2020 | EPI_ISL_1714176 |
| hCoV-19/Qatar/QA-QU_12-7-E6/2020 | EPI_ISL_1713313 |
| hCoV-19/Qatar/QA-QU_12-7-G1/2020 | EPI_ISL_1713318 |
| hCoV-19/Qatar/QA-WCMQ_FD18189992/2020 | EPI_ISL_1714471 |
| hCoV-19/Qatar/QA-WCMQ_FD18190004/2020 | EPI_ISL_1714483 |
| hCoV-19/Qatar/QA-WCMQ_FD18190011/2020 | EPI_ISL_1714490 |
| hCoV-19/Qatar/QA-WCMQ_FD18190021/2020 | EPI_ISL_1714499 |
| hCoV-19/Qatar/QA-WCMQ_FD18190027/2020 | EPI_ISL_1714505 |
| hCoV-19/Qatar/QA-WCMQ_FD18190042/2020 | EPI_ISL_1714519 |
| hCoV-19/Qatar/QA-WCMQ_FD18190053/2020 | EPI_ISL_1714528 |
| hCoV-19/Qatar/QA-WCMQ_FD18190062/2020 | EPI_ISL_1714536 |
| hCoV-19/Qatar/QA-WCMQ_FD18190063/2020 | EPI_ISL_1714537 |
| hCoV-19/Qatar/QA-WCMQ_FD18190071/2020 | EPI_ISL_1714545 |
| hCoV-19/Qatar/QA-QU_12-7-D12/2020 | EPI_ISL_1713306 |
| hCoV-19/Qatar/QA-QU_12-7-G7/2020 | EPI_ISL_1713319 |
| hCoV-19/Qatar/QA-WCMQ_FD18189997/2020 | EPI_ISL_1714476 |
| hCoV-19/Qatar/QA-WCMQ_FD18190019/2020 | EPI_ISL_1714497 |
| hCoV-19/Qatar/QA-WCMQ_FD18190028/2020 | EPI_ISL_1714506 |
| hCoV-19/Qatar/QA-WCMQ_FD18190030/2020 | EPI_ISL_1714508 |
| hCoV-19/Qatar/QA-WCMQ_FD18190049/2020 | EPI_ISL_1714524 |

| | |
|---|---|
| hCoV-19/Qatar/QA-QU_12-7-B3/2020 | EPI_ISL_1713302 |
| hCoV-19/Qatar/QA-WCMQ_FD18189987/2020 | EPI_ISL_1714466 |
| hCoV-19/Qatar/QA-WCMQ_FD18189991/2020 | EPI_ISL_1714470 |
| hCoV-19/Qatar/QA-WCMQ_FD18190000/2020 | EPI_ISL_1714479 |
| hCoV-19/Qatar/QA-WCMQ_FD18190001/2020 | EPI_ISL_1714480 |
| hCoV-19/Qatar/QA-WCMQ_FD18190006/2020 | EPI_ISL_1714485 |
| hCoV-19/Qatar/QA-WCMQ_FD18190022/2020 | EPI_ISL_1714500 |
| hCoV-19/Qatar/QA-WCMQ_FD18190025/2020 | EPI_ISL_1714503 |
| hCoV-19/Qatar/QA-WCMQ_FD18190031/2020 | EPI_ISL_1714509 |
| hCoV-19/Qatar/QA-WCMQ_FD18190037/2020 | EPI_ISL_1714515 |
| hCoV-19/Qatar/QA-WCMQ_FD18190044/2020 | EPI_ISL_1714520 |
| hCoV-19/Qatar/QA-WCMQ_FD18190074/2020 | EPI_ISL_1714548 |
| hCoV-19/Qatar/QA-QU_12-7-D9/2020 | EPI_ISL_1713310 |
| hCoV-19/Qatar/QA-QU_12-7-F8/2020 | EPI_ISL_1713317 |
| hCoV-19/Qatar/QA-WCMQ_FD18189990/2020 | EPI_ISL_1714469 |
| hCoV-19/Qatar/QA-WCMQ_FD18189998/2020 | EPI_ISL_1714477 |
| hCoV-19/Qatar/QA-WCMQ_FD18189999/2020 | EPI_ISL_1714478 |
| hCoV-19/Qatar/QA-WCMQ_FD18190018/2020 | EPI_ISL_1714496 |
| hCoV-19/Qatar/QA-WCMQ_FD18190026/2020 | EPI_ISL_1714504 |
| hCoV-19/Qatar/QA-WCMQ_FD18190035/2020 | EPI_ISL_1714513 |
| hCoV-19/Qatar/QA-WCMQ_FD18190039/2020 | EPI_ISL_1714517 |
| hCoV-19/Qatar/QA-WCMQ_FD18190045/2020 | EPI_ISL_1714521 |
| hCoV-19/Qatar/QA-WCMQ_FD18190046/2020 | EPI_ISL_1714522 |
| hCoV-19/Qatar/QA-WCMQ_FD18190050/2020 | EPI_ISL_1714525 |
| hCoV-19/Qatar/QA-WCMQ_FD18190051/2020 | EPI_ISL_1714526 |
| hCoV-19/Qatar/QA-WCMQ_FD18190070/2020 | EPI_ISL_1714544 |
| hCoV-19/Qatar/QA-WCMQ_FD18190023/2020 | EPI_ISL_1714501 |
| hCoV-19/Qatar/QA-WCMQ_FD18190033/2020 | EPI_ISL_1714511 |
| hCoV-19/Qatar/QA-WCMQ_FD18190036/2020 | EPI_ISL_1714514 |
| hCoV-19/Qatar/QA-WCMQ_FD18190024/2020 | EPI_ISL_1714502 |
| hCoV-19/Qatar/QA-WCMQ_FD18190060/2020 | EPI_ISL_1714535 |
| hCoV-19/Qatar/QA-QU_12-7-A12/2020 | EPI_ISL_1713298 |
| hCoV-19/Qatar/QA-WCMQ_FD17059313/2020 | EPI_ISL_1714170 |
| hCoV-19/Qatar/QA-WCMQ_FD18190075/2020 | EPI_ISL_1714549 |
| hCoV-19/Qatar/QA-WCMQ_FD18190067/2020 | EPI_ISL_1714541 |
| hCoV-19/Qatar/QA-WCMQ_FD18189981/2020 | EPI_ISL_1714461 |
| hCoV-19/Qatar/QA-WCMQ_FD18190013/2020 | EPI_ISL_1714492 |
| hCoV-19/Qatar/QA-WCMQ_FD18190012/2020 | EPI_ISL_1714491 |
| hCoV-19/Qatar/QA-WCMQ_FD18190016/2020 | EPI_ISL_1714494 |
| hCoV-19/Qatar/QA-WCMQ_FD18190059/2020 | EPI_ISL_1714534 |
| hCoV-19/Qatar/QA-QU_12-7-B8/2020 | EPI_ISL_1713303 |
| hCoV-19/Qatar/QA-WCMQ_FD18189980/2020 | EPI_ISL_1714460 |
| hCoV-19/Qatar/QA-WCMQ_FD18189983/2020 | EPI_ISL_1714462 |
| hCoV-19/Qatar/QA-WCMQ_FD18189984/2020 | EPI_ISL_1714463 |
| hCoV-19/Qatar/QA-WCMQ_FD18190014/2020 | EPI_ISL_1714493 |
| hCoV-19/Qatar/QA-QU_12-7-D8/2020 | EPI_ISL_1713309 |
| hCoV-19/Qatar/QA-QU_12-8-A2/2020 | EPI_ISL_1713329 |
| hCoV-19/Qatar/QA-WCMQ_FD18163117/2020 | EPI_ISL_1714387 |
| hCoV-19/Qatar/QA-WCMQ_FD18163158/2020 | EPI_ISL_1714426 |
| hCoV-19/Qatar/QA-WCMQ_FD18163188/2020 | EPI_ISL_1714456 |

| | |
|---|---|
| hCoV-19/Qatar/QA-WCMQ_FD17059329/2020 | EPI_ISL_1714181 |
| hCoV-19/Qatar/QA-WCMQ_FD18163112/2020 | EPI_ISL_1714382 |
| hCoV-19/Qatar/QA-WCMQ_FD18163113/2020 | EPI_ISL_1714383 |
| hCoV-19/Qatar/QA-WCMQ_FD18163114/2020 | EPI_ISL_1714384 |
| hCoV-19/Qatar/QA-WCMQ_FD18163115/2020 | EPI_ISL_1714385 |
| hCoV-19/Qatar/QA-WCMQ_FD18163116/2020 | EPI_ISL_1714386 |
| hCoV-19/Qatar/QA-WCMQ_FD18163145/2020 | EPI_ISL_1714414 |
| hCoV-19/Qatar/QA-WCMQ_FD18163168/2020 | EPI_ISL_1714436 |
| hCoV-19/Qatar/QA-WCMQ_FD18163171/2020 | EPI_ISL_1714439 |
| hCoV-19/Qatar/QA-QU_12-8-E10/2020 | EPI_ISL_1713345 |
| hCoV-19/Qatar/QA-QU_12-8-G1/2020 | EPI_ISL_1713355 |
| hCoV-19/Qatar/QA-WCMQ_FD18163173/2020 | EPI_ISL_1714441 |
| hCoV-19/Qatar/QA-WCMQ_FD18163184/2020 | EPI_ISL_1714452 |
| hCoV-19/Qatar/QA-QU_12-8-B4/2020 | EPI_ISL_1713334 |
| hCoV-19/Qatar/QA-QU_12-8-C11/2020 | EPI_ISL_1713338 |
| hCoV-19/Qatar/QA-WCMQ_FD18163122/2020 | EPI_ISL_1714392 |
| hCoV-19/Qatar/QA-QU_12-8-B9/2020 | EPI_ISL_1713336 |
| hCoV-19/Qatar/QA-QU_12-8-D10/2020 | EPI_ISL_1713342 |
| hCoV-19/Qatar/QA-QU_12-8-F9/2020 | EPI_ISL_1713354 |
| hCoV-19/Qatar/QA-WCMQ_FD18163162/2020 | EPI_ISL_1714430 |
| hCoV-19/Qatar/QA-QU_12-8-A12/2020 | EPI_ISL_1713328 |
| hCoV-19/Qatar/QA-QU_12-8-E6/2020 | EPI_ISL_1713347 |
| hCoV-19/Qatar/QA-WCMQ_FD18163119/2020 | EPI_ISL_1714389 |
| hCoV-19/Qatar/QA-WCMQ_FD18163143/2020 | EPI_ISL_1714412 |
| hCoV-19/Qatar/QA-WCMQ_FD18163147/2020 | EPI_ISL_1714416 |
| hCoV-19/Qatar/QA-WCMQ_FD18163150/2020 | EPI_ISL_1714419 |
| hCoV-19/Qatar/QA-WCMQ_FD18163190/2020 | EPI_ISL_1714458 |
| hCoV-19/Qatar/QA-QU_12-8-C8/2020 | EPI_ISL_1713341 |
| hCoV-19/Qatar/QA-QU_12-8-A4/2020 | EPI_ISL_1713330 |
| hCoV-19/Qatar/QA-QU_12-8-C10/2020 | EPI_ISL_1713337 |
| hCoV-19/Qatar/QA-QU_12-8-G11/2020 | EPI_ISL_1713356 |
| hCoV-19/Qatar/QA-WCMQ_FD18163106/2020 | EPI_ISL_1714376 |
| hCoV-19/Qatar/QA-WCMQ_FD18163109/2020 | EPI_ISL_1714379 |
| hCoV-19/Qatar/QA-WCMQ_FD18163159/2020 | EPI_ISL_1714427 |
| hCoV-19/Qatar/QA-WCMQ_FD18163187/2020 | EPI_ISL_1714455 |
| hCoV-19/Qatar/QA-QU_12-8-F10/2020 | EPI_ISL_1713349 |
| hCoV-19/Qatar/QA-WCMQ_FD18163105/2020 | EPI_ISL_1714375 |
| hCoV-19/Qatar/QA-WCMQ_FD18163124/2020 | EPI_ISL_1714394 |
| hCoV-19/Qatar/QA-WCMQ_FD18163139/2020 | EPI_ISL_1714408 |
| hCoV-19/Qatar/QA-WCMQ_FD18163140/2020 | EPI_ISL_1714409 |
| hCoV-19/Qatar/QA-WCMQ_FD18163153/2020 | EPI_ISL_1714421 |
| hCoV-19/Qatar/QA-WCMQ_FD18163156/2020 | EPI_ISL_1714424 |
| hCoV-19/Qatar/QA-WCMQ_FD18163172/2020 | EPI_ISL_1714440 |
| hCoV-19/Qatar/QA-QU_12-8-C7/2020 | EPI_ISL_1713340 |
| hCoV-19/Qatar/QA-QU_12-8-G6/2020 | EPI_ISL_1713358 |
| hCoV-19/Qatar/QA-WCMQ_FD15519042/2020 | EPI_ISL_1714046 |
| hCoV-19/Qatar/QA-WCMQ_FD18163104/2020 | EPI_ISL_1714374 |
| hCoV-19/Qatar/QA-WCMQ_FD18163110/2020 | EPI_ISL_1714380 |
| hCoV-19/Qatar/QA-WCMQ_FD18163121/2020 | EPI_ISL_1714391 |
| hCoV-19/Qatar/QA-WCMQ_FD18163126/2020 | EPI_ISL_1714396 |

| | |
|---|---|
| hCoV-19/Qatar/QA-WCMQ_FD18163128/2020 | EPI_ISL_1714397 |
| hCoV-19/Qatar/QA-WCMQ_FD18163155/2020 | EPI_ISL_1714423 |
| hCoV-19/Qatar/QA-WCMQ_FD18163163/2020 | EPI_ISL_1714431 |
| hCoV-19/Qatar/QA-WCMQ_FD18163166/2020 | EPI_ISL_1714434 |
| hCoV-19/Qatar/QA-WCMQ_FD18163167/2020 | EPI_ISL_1714435 |
| hCoV-19/Qatar/QA-WCMQ_FD18163185/2020 | EPI_ISL_1714453 |
| hCoV-19/Qatar/QA-WCMQ_FD18163186/2020 | EPI_ISL_1714454 |
| hCoV-19/Qatar/QA-QU_12-8-A7/2020 | EPI_ISL_1713331 |
| hCoV-19/Qatar/QA-QU_12-8-E9/2020 | EPI_ISL_1713348 |
| hCoV-19/Qatar/QA-QU_12-8-H2/2020 | EPI_ISL_1713360 |
| hCoV-19/Qatar/QA-WCMQ_FD18163123/2020 | EPI_ISL_1714393 |
| hCoV-19/Qatar/QA-WCMQ_FD18163125/2020 | EPI_ISL_1714395 |
| hCoV-19/Qatar/QA-WCMQ_FD18163131/2020 | EPI_ISL_1714400 |
| hCoV-19/Qatar/QA-WCMQ_FD18163136/2020 | EPI_ISL_1714405 |
| hCoV-19/Qatar/QA-WCMQ_FD18163157/2020 | EPI_ISL_1714425 |
| hCoV-19/Qatar/QA-WCMQ_FD18163160/2020 | EPI_ISL_1714428 |
| hCoV-19/Qatar/QA-WCMQ_FD18163174/2020 | EPI_ISL_1714442 |
| hCoV-19/Qatar/QA-WCMQ_FD18163182/2020 | EPI_ISL_1714450 |
| hCoV-19/Qatar/QA-QU_12-8-D5/2020 | EPI_ISL_1713343 |
| hCoV-19/Qatar/QA-WCMQ_FD18163107/2020 | EPI_ISL_1714377 |
| hCoV-19/Qatar/QA-WCMQ_FD18163135/2020 | EPI_ISL_1714404 |
| hCoV-19/Qatar/QA-WCMQ_FD18163178/2020 | EPI_ISL_1714446 |
| hCoV-19/Qatar/QA-QU_12-8-A9/2020 | EPI_ISL_1713332 |
| hCoV-19/Qatar/QA-QU_12-8-C3/2020 | EPI_ISL_1713339 |
| hCoV-19/Qatar/QA-WCMQ_FD18163144/2020 | EPI_ISL_1714413 |
| hCoV-19/Qatar/QA-WCMQ_FD18163148/2020 | EPI_ISL_1714417 |
| hCoV-19/Qatar/QA-WCMQ_FD18163154/2020 | EPI_ISL_1714422 |
| hCoV-19/Qatar/QA-WCMQ_FD18163170/2020 | EPI_ISL_1714438 |
| hCoV-19/Qatar/QA-WCMQ_FD18163176/2020 | EPI_ISL_1714444 |
| hCoV-19/Qatar/QA-WCMQ_FD18163177/2020 | EPI_ISL_1714445 |
| hCoV-19/Qatar/QA-WCMQ_FD18163180/2020 | EPI_ISL_1714448 |
| hCoV-19/Qatar/QA-QU_12-8-F12/2020 | EPI_ISL_1713350 |
| hCoV-19/Qatar/QA-QU_12-8-H10/2020 | EPI_ISL_1713359 |
| hCoV-19/Qatar/QA-WCMQ_FD18163108/2020 | EPI_ISL_1714378 |
| hCoV-19/Qatar/QA-WCMQ_FD18163137/2020 | EPI_ISL_1714406 |
| hCoV-19/Qatar/QA-WCMQ_FD18163149/2020 | EPI_ISL_1714418 |
| hCoV-19/Qatar/QA-WCMQ_FD18163169/2020 | EPI_ISL_1714437 |
| hCoV-19/Qatar/QA-WCMQ_FD18163179/2020 | EPI_ISL_1714447 |
| hCoV-19/Qatar/QA-QU_12-8-F5/2020 | EPI_ISL_1713351 |
| hCoV-19/Qatar/QA-QU_13-C2/2020 | EPI_ISL_1713371 |
| hCoV-19/Qatar/QA-WCMQ_FD18163102/2020 | EPI_ISL_1714372 |
| hCoV-19/Qatar/QA-WCMQ_FD18163111/2020 | EPI_ISL_1714381 |
| hCoV-19/Qatar/QA-WCMQ_FD18163120/2020 | EPI_ISL_1714390 |
| hCoV-19/Qatar/QA-WCMQ_FD18163129/2020 | EPI_ISL_1714398 |
| hCoV-19/Qatar/QA-WCMQ_FD18163130/2020 | EPI_ISL_1714399 |
| hCoV-19/Qatar/QA-WCMQ_FD18163189/2020 | EPI_ISL_1714457 |
| hCoV-19/Qatar/QA-QU_12-8-B7/2020 | EPI_ISL_1713335 |
| hCoV-19/Qatar/QA-WCMQ_FD17059328/2020 | EPI_ISL_1714180 |
| hCoV-19/Qatar/QA-WCMQ_FD18163097/2020 | EPI_ISL_1714367 |
| hCoV-19/Qatar/QA-WCMQ_FD18163101/2020 | EPI_ISL_1714371 |

| | |
|---|---|
| hCoV-19/Qatar/QA-WCMQ_FD18163132/2020 | EPI_ISL_1714401 |
| hCoV-19/Qatar/QA-WCMQ_FD18163146/2020 | EPI_ISL_1714415 |
| hCoV-19/Qatar/QA-WCMQ_FD18163191/2020 | EPI_ISL_1714459 |
| hCoV-19/Qatar/QA-QU_12-8-D6/2020 | EPI_ISL_1713344 |
| hCoV-19/Qatar/QA-QU_12-8-F6/2020 | EPI_ISL_1713352 |
| hCoV-19/Qatar/QA-QU_13-B2/2020 | EPI_ISL_1713367 |
| hCoV-19/Qatar/QA-WCMQ_FD18163098/2020 | EPI_ISL_1714368 |
| hCoV-19/Qatar/QA-WCMQ_FD18163099/2020 | EPI_ISL_1714369 |
| hCoV-19/Qatar/QA-WCMQ_FD18163161/2020 | EPI_ISL_1714429 |
| hCoV-19/Qatar/QA-WCMQ_FD18163181/2020 | EPI_ISL_1714449 |
| hCoV-19/Qatar/QA-QU_12-8-A11/2020 | EPI_ISL_1713327 |
| hCoV-19/Qatar/QA-QU_12-8-G3/2020 | EPI_ISL_1713357 |
| hCoV-19/Qatar/QA-WCMQ_FD18163096/2020 | EPI_ISL_1714366 |
| hCoV-19/Qatar/QA-WCMQ_FD18163100/2020 | EPI_ISL_1714370 |
| hCoV-19/Qatar/QA-WCMQ_FD18163103/2020 | EPI_ISL_1714373 |
| hCoV-19/Qatar/QA-WCMQ_FD18163118/2020 | EPI_ISL_1714388 |
| hCoV-19/Qatar/QA-WCMQ_FD18163133/2020 | EPI_ISL_1714402 |
| hCoV-19/Qatar/QA-WCMQ_FD18163138/2020 | EPI_ISL_1714407 |
| hCoV-19/Qatar/QA-WCMQ_FD18163142/2020 | EPI_ISL_1714411 |
| hCoV-19/Qatar/QA-WCMQ_FD18163151/2020 | EPI_ISL_1714420 |
| hCoV-19/Qatar/QA-WCMQ_FD18163164/2020 | EPI_ISL_1714432 |
| hCoV-19/Qatar/QA-WCMQ_FD18163165/2020 | EPI_ISL_1714433 |
| hCoV-19/Qatar/QA-WCMQ_FD18163183/2020 | EPI_ISL_1714451 |
| hCoV-19/Qatar/QA-QU_12-8-E5/2020 | EPI_ISL_1713346 |
| hCoV-19/Qatar/QA-WCMQ_FD18163134/2020 | EPI_ISL_1714403 |
| hCoV-19/Qatar/QA-WCMQ_FD18163141/2020 | EPI_ISL_1714410 |
| hCoV-19/Qatar/QA-WCMQ_FD18163175/2020 | EPI_ISL_1714443 |
| hCoV-19/Qatar/QA-QU_12-8-B3/2020 | EPI_ISL_1713333 |
| hCoV-19/Qatar/QA-QU_12-8-F8/2020 | EPI_ISL_1713353 |
| hCoV-19/Qatar/QA-WCMQ_FD17059307/2020 | EPI_ISL_1714164 |
| hCoV-19/Qatar/QA-QU_12-8-H8/2020 | EPI_ISL_1713361 |
| hCoV-19/Qatar/QA-WCMQ_FD15542781/2020 | EPI_ISL_1714077 |
| hCoV-19/Qatar/QA-WCMQ_FD15692498/2020 | EPI_ISL_1714162 |
| hCoV-19/Qatar/QA-WCMQ_FD17059334/2020 | EPI_ISL_1714184 |
| hCoV-19/Qatar/QA-WCMQ_FD18231586/2020 | EPI_ISL_1714668 |
| hCoV-19/Qatar/QA-QU_10-1-A8/2020 | EPI_ISL_1713145 |
| hCoV-19/Qatar/QA-WCMQ_FD17059336/2020 | EPI_ISL_1714185 |
| hCoV-19/Qatar/QA-QU_10-2-F8/2020 | EPI_ISL_1713178 |
| hCoV-19/Qatar/QA-QU_10-2-A1/2020 | EPI_ISL_1713146 |
| hCoV-19/Qatar/QA-QU_10-2-B5/2020 | EPI_ISL_1713154 |
| hCoV-19/Qatar/QA-QU_10-2-C6/2020 | EPI_ISL_1713157 |
| hCoV-19/Qatar/QA-QU_10-2-F4/2020 | EPI_ISL_1713174 |
| hCoV-19/Qatar/QA-QU_10-2-F5/2020 | EPI_ISL_1713175 |
| hCoV-19/Qatar/QA-QU_10-2-F6/2020 | EPI_ISL_1713176 |
| hCoV-19/Qatar/QA-QU_10-2-G2/2020 | EPI_ISL_1713180 |
| hCoV-19/Qatar/QA-QU_10-2-H6/2020 | EPI_ISL_1713192 |
| hCoV-19/Qatar/QA-QU_10-1-A4/2020 | EPI_ISL_1713141 |
| hCoV-19/Qatar/QA-QU_10-1-A5/2020 | EPI_ISL_1713142 |
| hCoV-19/Qatar/QA-QU_10-1-A7/2020 | EPI_ISL_1713144 |
| hCoV-19/Qatar/QA-QU_10-2-A3/2020 | EPI_ISL_1713147 |

| | |
|---|---|
| hCoV-19/Qatar/QA-QU_10-2-A4/2020 | EPI_ISL_1713148 |
| hCoV-19/Qatar/QA-QU_10-2-A6/2020 | EPI_ISL_1713149 |
| hCoV-19/Qatar/QA-QU_10-2-A7/2020 | EPI_ISL_1713150 |
| hCoV-19/Qatar/QA-QU_10-2-A9/2020 | EPI_ISL_1713151 |
| hCoV-19/Qatar/QA-QU_10-2-B1/2020 | EPI_ISL_1713152 |
| hCoV-19/Qatar/QA-QU_10-2-B2/2020 | EPI_ISL_1713153 |
| hCoV-19/Qatar/QA-QU_10-2-B7/2020 | EPI_ISL_1713155 |
| hCoV-19/Qatar/QA-QU_10-2-C1/2020 | EPI_ISL_1713156 |
| hCoV-19/Qatar/QA-QU_10-2-C9/2020 | EPI_ISL_1713158 |
| hCoV-19/Qatar/QA-QU_10-2-D1/2020 | EPI_ISL_1713159 |
| hCoV-19/Qatar/QA-QU_10-2-D3/2020 | EPI_ISL_1713160 |
| hCoV-19/Qatar/QA-QU_10-2-D5/2020 | EPI_ISL_1713161 |
| hCoV-19/Qatar/QA-QU_10-2-D9/2020 | EPI_ISL_1713164 |
| hCoV-19/Qatar/QA-QU_10-2-E2/2020 | EPI_ISL_1713165 |
| hCoV-19/Qatar/QA-QU_10-2-E3/2020 | EPI_ISL_1713166 |
| hCoV-19/Qatar/QA-QU_10-2-E4/2020 | EPI_ISL_1713167 |
| hCoV-19/Qatar/QA-QU_10-2-E5/2020 | EPI_ISL_1713168 |
| hCoV-19/Qatar/QA-QU_10-2-E6/2020 | EPI_ISL_1713169 |
| hCoV-19/Qatar/QA-QU_10-2-E7/2020 | EPI_ISL_1713170 |
| hCoV-19/Qatar/QA-QU_10-2-E8/2020 | EPI_ISL_1713171 |
| hCoV-19/Qatar/QA-QU_10-2-F1/2020 | EPI_ISL_1713172 |
| hCoV-19/Qatar/QA-QU_10-2-F7/2020 | EPI_ISL_1713177 |
| hCoV-19/Qatar/QA-QU_10-2-G1/2020 | EPI_ISL_1713179 |
| hCoV-19/Qatar/QA-QU_10-2-G3/2020 | EPI_ISL_1713181 |
| hCoV-19/Qatar/QA-QU_10-2-G4/2020 | EPI_ISL_1713182 |
| hCoV-19/Qatar/QA-QU_10-2-G5/2020 | EPI_ISL_1713183 |
| hCoV-19/Qatar/QA-QU_10-2-G6/2020 | EPI_ISL_1713184 |
| hCoV-19/Qatar/QA-QU_10-2-G8/2020 | EPI_ISL_1713186 |
| hCoV-19/Qatar/QA-QU_10-2-H3/2020 | EPI_ISL_1713189 |
| hCoV-19/Qatar/QA-QU_10-2-H5/2020 | EPI_ISL_1713191 |
| hCoV-19/Qatar/QA-QU_10-1-A1/2020 | EPI_ISL_1713139 |
| hCoV-19/Qatar/QA-QU_10-2-D6/2020 | EPI_ISL_1713162 |
| hCoV-19/Qatar/QA-QU_10-2-D7/2020 | EPI_ISL_1713163 |
| hCoV-19/Qatar/QA-QU_10-2-F2/2020 | EPI_ISL_1713173 |
| hCoV-19/Qatar/QA-QU_10-2-H1/2020 | EPI_ISL_1713187 |
| hCoV-19/Qatar/QA-QU_10-2-H2/2020 | EPI_ISL_1713188 |
| hCoV-19/Qatar/QA-QU_10-2-H8/2020 | EPI_ISL_1713193 |
| hCoV-19/Qatar/QA-QU_10-1-A6/2020 | EPI_ISL_1713143 |
| hCoV-19/Qatar/QA-QU_10-2-H4/2020 | EPI_ISL_1713190 |
| hCoV-19/Qatar/QA-QU_10-1-A2/2020 | EPI_ISL_1713140 |
| hCoV-19/Qatar/QA-WCMQ_FD17059326/2020 | EPI_ISL_1714179 |
| hCoV-19/Qatar/QA-QU_10-2-G7/2020 | EPI_ISL_1713185 |
| hCoV-19/Qatar/QA-WCMQ_FD17059332/2020 | EPI_ISL_1714183 |
| hCoV-19/Qatar/QA-QU_17-B1/2020 | EPI_ISL_1713473 |
| hCoV-19/Qatar/QA-QU_17-A6/2020 | EPI_ISL_1713471 |
| hCoV-19/Qatar/QA-QU_17-B6/2020 | EPI_ISL_1713475 |
| hCoV-19/Qatar/QA-QU_17-B7/2020 | EPI_ISL_1713476 |
| hCoV-19/Qatar/QA-QU_17-B10/2020 | EPI_ISL_1713474 |
| hCoV-19/Qatar/QA-QU_17-A3/2020 | EPI_ISL_1713469 |
| hCoV-19/Qatar/QA-QU_17-A7/2020 | EPI_ISL_1713472 |

| | |
|---|---|
| hCoV-19/Qatar/QA-QU_17-C1/2020 | EPI_ISL_1713477 |
| hCoV-19/Qatar/QA-QU_17-A2/2020 | EPI_ISL_1713468 |
| hCoV-19/Qatar/QA-QU_17-C4/2020 | EPI_ISL_1713478 |
| hCoV-19/Qatar/QA-QU_17-C5/2020 | EPI_ISL_1713479 |
| hCoV-19/Qatar/QA-QU_13-A2/2020 | EPI_ISL_1713363 |
| hCoV-19/Qatar/QA-QU_13-A3/2020 | EPI_ISL_1713364 |
| hCoV-19/Qatar/QA-QU_13-H1/2020 | EPI_ISL_1713377 |
| hCoV-19/Qatar/QA-QU_17-C6/2020 | EPI_ISL_1713480 |
| hCoV-19/Qatar/QA-QU_17-C7/2020 | EPI_ISL_1713481 |
| hCoV-19/Qatar/QA-QU_17-C8/2020 | EPI_ISL_1713482 |
| hCoV-19/Qatar/QA-QU_17-C9/2020 | EPI_ISL_1713483 |
| hCoV-19/Qatar/QA-QU_13-D1/2020 | EPI_ISL_1713373 |
| hCoV-19/Qatar/QA-QU_13-G1/2020 | EPI_ISL_1713376 |
| hCoV-19/Qatar/QA-QU_17-A4/2020 | EPI_ISL_1713470 |
| hCoV-19/Qatar/QA-WCMQ_FD17059331/2020 | EPI_ISL_1714182 |
| hCoV-19/Qatar/QA-QU_13-C1/2020 | EPI_ISL_1713370 |
| hCoV-19/Qatar/QA-QU_13-A1/2020 | EPI_ISL_1713362 |
| hCoV-19/Qatar/QA-QU_13-B1/2020 | EPI_ISL_1713366 |
| hCoV-19/Qatar/QA-QU_12-10-A8/2020 | EPI_ISL_1713201 |
| hCoV-19/Qatar/QA-QU_12-10-B4/2020 | EPI_ISL_1713205 |
| hCoV-19/Qatar/QA-QU_12-10-B10/2020 | EPI_ISL_1713203 |
| hCoV-19/Qatar/QA-QU_12-10-H10/2020 | EPI_ISL_1713233 |
| hCoV-19/Qatar/QA-QU_12-10-H6/2020 | EPI_ISL_1713236 |
| hCoV-19/Qatar/QA-QU_12-10-D8/2020 | EPI_ISL_1713219 |
| hCoV-19/Qatar/QA-QU_12-10-A2/2020 | EPI_ISL_1713197 |
| hCoV-19/Qatar/QA-QU_12-10-A3/2020 | EPI_ISL_1713198 |
| hCoV-19/Qatar/QA-QU_12-10-B1/2020 | EPI_ISL_1713202 |
| hCoV-19/Qatar/QA-QU_12-10-B2/2020 | EPI_ISL_1713204 |
| hCoV-19/Qatar/QA-QU_12-10-B6/2020 | EPI_ISL_1713206 |
| hCoV-19/Qatar/QA-QU_12-10-C2/2020 | EPI_ISL_1713208 |
| hCoV-19/Qatar/QA-QU_12-10-D1/2020 | EPI_ISL_1713213 |
| hCoV-19/Qatar/QA-QU_12-10-D6/2020 | EPI_ISL_1713217 |
| hCoV-19/Qatar/QA-QU_12-10-F2/2020 | EPI_ISL_1713227 |
| hCoV-19/Qatar/QA-QU_12-10-A7/2020 | EPI_ISL_1713200 |
| hCoV-19/Qatar/QA-QU_12-10-C6/2020 | EPI_ISL_1713210 |
| hCoV-19/Qatar/QA-QU_12-10-F6/2020 | EPI_ISL_1713229 |
| hCoV-19/Qatar/QA-QU_12-10-D12/2020 | EPI_ISL_1713216 |
| hCoV-19/Qatar/QA-QU_12-10-F10/2020 | EPI_ISL_1713225 |
| hCoV-19/Qatar/QA-QU_12-11-E2/2020 | EPI_ISL_1713263 |
| hCoV-19/Qatar/QA-QU_12-11-H2/2020 | EPI_ISL_1713281 |
| hCoV-19/Qatar/QA-QU_12-10-E3/2020 | EPI_ISL_1713222 |
| hCoV-19/Qatar/QA-QU_12-10-H7/2020 | EPI_ISL_1713237 |
| hCoV-19/Qatar/QA-QU_12-10-C3/2020 | EPI_ISL_1713209 |
| hCoV-19/Qatar/QA-QU_12-10-C8/2020 | EPI_ISL_1713211 |
| hCoV-19/Qatar/QA-QU_12-10-E9/2020 | EPI_ISL_1713224 |
| hCoV-19/Qatar/QA-QU_12-10-A6/2020 | EPI_ISL_1713199 |
| hCoV-19/Qatar/QA-QU_12-10-F4/2020 | EPI_ISL_1713228 |
| hCoV-19/Qatar/QA-QU_12-10-F8/2020 | EPI_ISL_1713230 |
| hCoV-19/Qatar/QA-QU_12-10-G12/2020 | EPI_ISL_1713232 |
| hCoV-19/Qatar/QA-QU_12-10-H3/2020 | EPI_ISL_1713235 |

| | |
|---|---|
| hCoV-19/Qatar/QA-QU_12-10-H8/2020 | EPI_ISL_1713238 |
| hCoV-19/Qatar/QA-QU_12-10-C9/2020 | EPI_ISL_1713212 |
| hCoV-19/Qatar/QA-QU_12-10-G10/2020 | EPI_ISL_1713231 |
| hCoV-19/Qatar/QA-QU_12-10-A10/2020 | EPI_ISL_1713194 |
| hCoV-19/Qatar/QA-QU_12-10-H11/2020 | EPI_ISL_1713234 |
| hCoV-19/Qatar/QA-QU_12-10-D9/2020 | EPI_ISL_1713220 |
| hCoV-19/Qatar/QA-QU_12-10-E7/2020 | EPI_ISL_1713223 |
| hCoV-19/Qatar/QA-QU_12-10-B8/2020 | EPI_ISL_1713207 |
| hCoV-19/Qatar/QA-QU_12-10-F11/2020 | EPI_ISL_1713226 |
| hCoV-19/Qatar/QA-QU_12-10-A11/2020 | EPI_ISL_1713195 |
| hCoV-19/Qatar/QA-QU_12-10-A12/2020 | EPI_ISL_1713196 |
| hCoV-19/Qatar/QA-QU_12-10-D10/2020 | EPI_ISL_1713214 |
| hCoV-19/Qatar/QA-QU_12-10-D11/2020 | EPI_ISL_1713215 |
| hCoV-19/Qatar/QA-QU_12-10-D7/2020 | EPI_ISL_1713218 |
| hCoV-19/Qatar/QA-QU_12-10-E10/2020 | EPI_ISL_1713221 |
| hCoV-19/Qatar/QA-QU_12-11-H11/2020 | EPI_ISL_1713279 |
| hCoV-19/Qatar/QA-QU_13-B3/2020 | EPI_ISL_1713368 |
| hCoV-19/Qatar/QA-QU_12-11-E11/2020 | EPI_ISL_1713261 |
| hCoV-19/Qatar/QA-QU_12-11-E9/2020 | EPI_ISL_1713267 |
| hCoV-19/Qatar/QA-QU_12-11-B3/2020 | EPI_ISL_1713250 |
| hCoV-19/Qatar/QA-QU_12-11-F11/2020 | EPI_ISL_1713268 |
| hCoV-19/Qatar/QA-QU_12-11-G1/2020 | EPI_ISL_1713275 |
| hCoV-19/Qatar/QA-QU_12-11-G10/2020 | EPI_ISL_1713276 |
| hCoV-19/Qatar/QA-QU_12-11-B2/2020 | EPI_ISL_1713249 |
| hCoV-19/Qatar/QA-QU_12-11-F9/2020 | EPI_ISL_1713274 |
| hCoV-19/Qatar/QA-QU_12-11-E12/2020 | EPI_ISL_1713262 |
| hCoV-19/Qatar/QA-QU_12-11-H10/2020 | EPI_ISL_1713278 |
| hCoV-19/Qatar/QA-QU_12-11-C9/2020 | EPI_ISL_1713256 |
| hCoV-19/Qatar/QA-QU_12-11-D3/2020 | EPI_ISL_1713258 |
| hCoV-19/Qatar/QA-QU_12-11-A12/2020 | EPI_ISL_1713240 |
| hCoV-19/Qatar/QA-QU_12-11-B4/2020 | EPI_ISL_1713251 |
| hCoV-19/Qatar/QA-QU_12-11-A2/2020 | EPI_ISL_1713241 |
| hCoV-19/Qatar/QA-QU_12-11-B10/2020 | EPI_ISL_1713247 |
| hCoV-19/Qatar/QA-QU_12-11-D1/2020 | EPI_ISL_1713257 |
| hCoV-19/Qatar/QA-QU_12-11-A10/2020 | EPI_ISL_1713239 |
| hCoV-19/Qatar/QA-QU_12-11-D4/2020 | EPI_ISL_1713259 |
| hCoV-19/Qatar/QA-QU_12-11-E3/2020 | EPI_ISL_1713264 |
| hCoV-19/Qatar/QA-QU_13-D2/2020 | EPI_ISL_1713374 |
| hCoV-19/Qatar/QA-QU_12-11-B1/2020 | EPI_ISL_1713246 |
| hCoV-19/Qatar/QA-QU_12-11-F12/2020 | EPI_ISL_1713269 |
| hCoV-19/Qatar/QA-QU_12-11-B12/2020 | EPI_ISL_1713248 |
| hCoV-19/Qatar/QA-QU_12-11-F8/2020 | EPI_ISL_1713273 |
| hCoV-19/Qatar/QA-QU_12-11-H9/2020 | EPI_ISL_1713285 |
| hCoV-19/Qatar/QA-QU_12-11-F3/2020 | EPI_ISL_1713270 |
| hCoV-19/Qatar/QA-QU_12-11-H12/2020 | EPI_ISL_1713280 |
| hCoV-19/Qatar/QA-QU_12-11-H3/2020 | EPI_ISL_1713282 |
| hCoV-19/Qatar/QA-QU_12-11-A5/2020 | EPI_ISL_1713242 |
| hCoV-19/Qatar/QA-QU_12-11-B5/2020 | EPI_ISL_1713252 |
| hCoV-19/Qatar/QA-QU_12-11-B9/2020 | EPI_ISL_1713254 |
| hCoV-19/Qatar/QA-QU_12-11-C6/2020 | EPI_ISL_1713255 |

| | |
|---|---|
| hCoV-19/Qatar/QA-QU_12-11-H4/2020 | EPI_ISL_1713283 |
| hCoV-19/Qatar/QA-QU_12-11-H7/2020 | EPI_ISL_1713284 |
| hCoV-19/Qatar/QA-QU_13-C3/2020 | EPI_ISL_1713372 |
| hCoV-19/Qatar/QA-QU_12-11-A6/2020 | EPI_ISL_1713243 |
| hCoV-19/Qatar/QA-QU_12-11-D8/2020 | EPI_ISL_1713260 |
| hCoV-19/Qatar/QA-QU_12-11-G6/2020 | EPI_ISL_1713277 |
| hCoV-19/Qatar/QA-QU_12-11-A7/2020 | EPI_ISL_1713244 |
| hCoV-19/Qatar/QA-QU_12-11-B6/2020 | EPI_ISL_1713253 |
| hCoV-19/Qatar/QA-QU_12-11-E8/2020 | EPI_ISL_1713266 |
| hCoV-19/Qatar/QA-QU_12-11-F5/2020 | EPI_ISL_1713271 |
| hCoV-19/Qatar/QA-QU_12-11-F6/2020 | EPI_ISL_1713272 |
| hCoV-19/Qatar/QA-QU_12-11-A8/2020 | EPI_ISL_1713245 |
| hCoV-19/Qatar/QA-QU_12-11-E7/2020 | EPI_ISL_1713265 |
| hCoV-19/Qatar/QA-QU_13-A4/2020 | EPI_ISL_1713365 |
| hCoV-19/Qatar/QA-QU_13-B4/2020 | EPI_ISL_1713369 |
| hCoV-19/Qatar/QA-QU_13-E3/2020 | EPI_ISL_1713375 |
| hCoV-19/Qatar/QA-QU_15-1-B4/2020 | EPI_ISL_1713382 |
| hCoV-19/Qatar/QA-QU_15-1-F3/2020 | EPI_ISL_1713386 |
| hCoV-19/Qatar/QA-QU_15-1-A8/2020 | EPI_ISL_1713381 |
| hCoV-19/Qatar/QA-QU_15-1-C8/2020 | EPI_ISL_1713383 |
| hCoV-19/Qatar/QA-QU_15-1-E8/2020 | EPI_ISL_1713385 |
| hCoV-19/Qatar/QA-QU_15-1-G8/2020 | EPI_ISL_1713388 |
| hCoV-19/Qatar/QA-QU_15-1-H7/2020 | EPI_ISL_1713389 |
| hCoV-19/Qatar/QA-QU_15-1-E7/2020 | EPI_ISL_1713384 |
| hCoV-19/Qatar/QA-QU_15-1-G12/2020 | EPI_ISL_1713387 |
| hCoV-19/Qatar/QA-QU_15-5-F2/2020 | EPI_ISL_1713417 |
| hCoV-19/Qatar/QA-QU_17-H9/2020 | EPI_ISL_1713517 |
| hCoV-19/Qatar/QA-WCMQ_FD17571280/2020 | EPI_ISL_1714279 |
| hCoV-19/Qatar/QA-WCMQ_FD17571337/2020 | EPI_ISL_1714328 |
| hCoV-19/Qatar/QA-WCMQ_FD17571338/2020 | EPI_ISL_1714329 |
| hCoV-19/Qatar/QA-WCMQ_FD17571339/2020 | EPI_ISL_1714330 |
| hCoV-19/Qatar/QA-WCMQ_FD17571341/2020 | EPI_ISL_1714332 |
| hCoV-19/Qatar/QA-WCMQ_FD17571342/2020 | EPI_ISL_1714333 |
| hCoV-19/Qatar/QA-WCMQ_FD17571361/2020 | EPI_ISL_1714351 |
| hCoV-19/Qatar/QA-WCMQ_FD17571364/2020 | EPI_ISL_1714352 |
| hCoV-19/Qatar/QA-WCMQ_FD17571367/2020 | EPI_ISL_1714353 |
| hCoV-19/Qatar/QA-QU_15-4-D11/2020 | EPI_ISL_1713412 |
| hCoV-19/Qatar/QA-QU_15-4-F12/2020 | EPI_ISL_1713413 |
| hCoV-19/Qatar/QA-WCMQ_FD17571340/2020 | EPI_ISL_1714331 |
| hCoV-19/Qatar/QA-QU_15-2-G8/2020 | EPI_ISL_1713398 |
| hCoV-19/Qatar/QA-QU_15-2-H8/2020 | EPI_ISL_1713402 |
| hCoV-19/Qatar/QA-QU_15-2-F12/2020 | EPI_ISL_1713397 |
| hCoV-19/Qatar/QA-QU_14-C2/2020 | EPI_ISL_1713378 |
| hCoV-19/Qatar/QA-QU_15-2-A10/2020 | EPI_ISL_1713390 |
| hCoV-19/Qatar/QA-QU_15-2-C11/2020 | EPI_ISL_1713392 |
| hCoV-19/Qatar/QA-QU_15-2-H10/2020 | EPI_ISL_1713399 |
| hCoV-19/Qatar/QA-QU_15-2-H6/2020 | EPI_ISL_1713401 |
| hCoV-19/Qatar/QA-QU_14-C3/2020 | EPI_ISL_1713379 |
| hCoV-19/Qatar/QA-QU_14-C5/2020 | EPI_ISL_1713380 |
| hCoV-19/Qatar/QA-QU_15-2-A6/2020 | EPI_ISL_1713391 |

| | |
|---|---|
| hCoV-19/Qatar/QA-QU_15-2-C2/2020 | EPI_ISL_1713393 |
| hCoV-19/Qatar/QA-QU_15-2-H5/2020 | EPI_ISL_1713400 |
| hCoV-19/Qatar/QA-QU_17-D1/2020 | EPI_ISL_1713484 |
| hCoV-19/Qatar/QA-QU_15-4-F5/2020 | EPI_ISL_1713414 |
| hCoV-19/Qatar/QA-QU_17-D2/2020 | EPI_ISL_1713487 |
| hCoV-19/Qatar/QA-WCMQ_FD17571288/2020 | EPI_ISL_1714285 |
| hCoV-19/Qatar/QA-WCMQ_FD17571300/2020 | EPI_ISL_1714297 |
| hCoV-19/Qatar/QA-WCMQ_FD17571303/2020 | EPI_ISL_1714300 |
| hCoV-19/Qatar/QA-WCMQ_FD17571343/2020 | EPI_ISL_1714334 |
| hCoV-19/Qatar/QA-QU_15-4-C7/2020 | EPI_ISL_1713411 |
| hCoV-19/Qatar/QA-QU_15-4-F6/2020 | EPI_ISL_1713415 |
| hCoV-19/Qatar/QA-QU_15-5-A12/2020 | EPI_ISL_1713416 |
| hCoV-19/Qatar/QA-QU_16-E10/2020 | EPI_ISL_1713456 |
| hCoV-19/Qatar/QA-QU_16-E11/2020 | EPI_ISL_1713457 |
| hCoV-19/Qatar/QA-QU_16-E12/2020 | EPI_ISL_1713458 |
| hCoV-19/Qatar/QA-QU_16-E6/2020 | EPI_ISL_1713459 |
| hCoV-19/Qatar/QA-QU_16-E7/2020 | EPI_ISL_1713460 |
| hCoV-19/Qatar/QA-QU_16-E8/2020 | EPI_ISL_1713461 |
| hCoV-19/Qatar/QA-QU_16-E9/2020 | EPI_ISL_1713462 |
| hCoV-19/Qatar/QA-QU_16-F1/2020 | EPI_ISL_1713463 |
| hCoV-19/Qatar/QA-QU_16-F2/2020 | EPI_ISL_1713466 |
| hCoV-19/Qatar/QA-QU_16-F3/2020 | EPI_ISL_1713467 |
| hCoV-19/Qatar/QA-QU_17-D5/2020 | EPI_ISL_1713488 |
| hCoV-19/Qatar/QA-WCMQ_FD17571274/2020 | EPI_ISL_1714275 |
| hCoV-19/Qatar/QA-WCMQ_FD17571275/2020 | EPI_ISL_1714276 |
| hCoV-19/Qatar/QA-WCMQ_FD17571276/2020 | EPI_ISL_1714277 |
| hCoV-19/Qatar/QA-WCMQ_FD17571279/2020 | EPI_ISL_1714278 |
| hCoV-19/Qatar/QA-WCMQ_FD17571281/2020 | EPI_ISL_1714280 |
| hCoV-19/Qatar/QA-WCMQ_FD17571282/2020 | EPI_ISL_1714281 |
| hCoV-19/Qatar/QA-WCMQ_FD17571283/2020 | EPI_ISL_1714282 |
| hCoV-19/Qatar/QA-WCMQ_FD17571286/2020 | EPI_ISL_1714283 |
| hCoV-19/Qatar/QA-WCMQ_FD17571287/2020 | EPI_ISL_1714284 |
| hCoV-19/Qatar/QA-WCMQ_FD17571289/2020 | EPI_ISL_1714286 |
| hCoV-19/Qatar/QA-WCMQ_FD17571290/2020 | EPI_ISL_1714287 |
| hCoV-19/Qatar/QA-WCMQ_FD17571291/2020 | EPI_ISL_1714288 |
| hCoV-19/Qatar/QA-WCMQ_FD17571292/2020 | EPI_ISL_1714289 |
| hCoV-19/Qatar/QA-WCMQ_FD17571293/2020 | EPI_ISL_1714290 |
| hCoV-19/Qatar/QA-WCMQ_FD17571294/2020 | EPI_ISL_1714291 |
| hCoV-19/Qatar/QA-WCMQ_FD17571295/2020 | EPI_ISL_1714292 |
| hCoV-19/Qatar/QA-WCMQ_FD17571296/2020 | EPI_ISL_1714293 |
| hCoV-19/Qatar/QA-WCMQ_FD17571297/2020 | EPI_ISL_1714294 |
| hCoV-19/Qatar/QA-WCMQ_FD17571298/2020 | EPI_ISL_1714295 |
| hCoV-19/Qatar/QA-WCMQ_FD17571299/2020 | EPI_ISL_1714296 |
| hCoV-19/Qatar/QA-WCMQ_FD17571301/2020 | EPI_ISL_1714298 |
| hCoV-19/Qatar/QA-WCMQ_FD17571302/2020 | EPI_ISL_1714299 |
| hCoV-19/Qatar/QA-WCMQ_FD17571304/2020 | EPI_ISL_1714301 |
| hCoV-19/Qatar/QA-WCMQ_FD17571305/2020 | EPI_ISL_1714302 |
| hCoV-19/Qatar/QA-WCMQ_FD17571306/2020 | EPI_ISL_1714303 |
| hCoV-19/Qatar/QA-WCMQ_FD17571308/2020 | EPI_ISL_1714304 |
| hCoV-19/Qatar/QA-WCMQ_FD17571309/2020 | EPI_ISL_1714305 |

| | |
|---|---|
| hCoV-19/Qatar/QA-WCMQ_FD17571310/2020 | EPI_ISL_1714306 |
| hCoV-19/Qatar/QA-WCMQ_FD17571311/2020 | EPI_ISL_1714307 |
| hCoV-19/Qatar/QA-WCMQ_FD17571312/2020 | EPI_ISL_1714308 |
| hCoV-19/Qatar/QA-WCMQ_FD17571313/2020 | EPI_ISL_1714309 |
| hCoV-19/Qatar/QA-WCMQ_FD17571314/2020 | EPI_ISL_1714310 |
| hCoV-19/Qatar/QA-WCMQ_FD17571315/2020 | EPI_ISL_1714311 |
| hCoV-19/Qatar/QA-WCMQ_FD17571316/2020 | EPI_ISL_1714312 |
| hCoV-19/Qatar/QA-WCMQ_FD17571317/2020 | EPI_ISL_1714313 |
| hCoV-19/Qatar/QA-WCMQ_FD17571318/2020 | EPI_ISL_1714314 |
| hCoV-19/Qatar/QA-WCMQ_FD17571319/2020 | EPI_ISL_1714315 |
| hCoV-19/Qatar/QA-WCMQ_FD17571320/2020 | EPI_ISL_1714316 |
| hCoV-19/Qatar/QA-WCMQ_FD17571322/2020 | EPI_ISL_1714317 |
| hCoV-19/Qatar/QA-WCMQ_FD17571324/2020 | EPI_ISL_1714318 |
| hCoV-19/Qatar/QA-WCMQ_FD17571325/2020 | EPI_ISL_1714319 |
| hCoV-19/Qatar/QA-WCMQ_FD17571326/2020 | EPI_ISL_1714320 |
| hCoV-19/Qatar/QA-WCMQ_FD17571327/2020 | EPI_ISL_1714321 |
| hCoV-19/Qatar/QA-WCMQ_FD17571328/2020 | EPI_ISL_1714322 |
| hCoV-19/Qatar/QA-WCMQ_FD17571329/2020 | EPI_ISL_1714323 |
| hCoV-19/Qatar/QA-WCMQ_FD17571332/2020 | EPI_ISL_1714324 |
| hCoV-19/Qatar/QA-WCMQ_FD17571333/2020 | EPI_ISL_1714325 |
| hCoV-19/Qatar/QA-WCMQ_FD17571335/2020 | EPI_ISL_1714326 |
| hCoV-19/Qatar/QA-WCMQ_FD17571336/2020 | EPI_ISL_1714327 |
| hCoV-19/Qatar/QA-WCMQ_FD17571344/2020 | EPI_ISL_1714335 |
| hCoV-19/Qatar/QA-WCMQ_FD17571345/2020 | EPI_ISL_1714336 |
| hCoV-19/Qatar/QA-WCMQ_FD17571346/2020 | EPI_ISL_1714337 |
| hCoV-19/Qatar/QA-WCMQ_FD17571347/2020 | EPI_ISL_1714338 |
| hCoV-19/Qatar/QA-WCMQ_FD17571348/2020 | EPI_ISL_1714339 |
| hCoV-19/Qatar/QA-WCMQ_FD17571349/2020 | EPI_ISL_1714340 |
| hCoV-19/Qatar/QA-WCMQ_FD17571350/2020 | EPI_ISL_1714341 |
| hCoV-19/Qatar/QA-WCMQ_FD17571351/2020 | EPI_ISL_1714342 |
| hCoV-19/Qatar/QA-WCMQ_FD17571352/2020 | EPI_ISL_1714343 |
| hCoV-19/Qatar/QA-WCMQ_FD17571353/2020 | EPI_ISL_1714344 |
| hCoV-19/Qatar/QA-WCMQ_FD17571354/2020 | EPI_ISL_1714345 |
| hCoV-19/Qatar/QA-WCMQ_FD17571355/2020 | EPI_ISL_1714346 |
| hCoV-19/Qatar/QA-WCMQ_FD17571356/2020 | EPI_ISL_1714347 |
| hCoV-19/Qatar/QA-WCMQ_FD17571357/2020 | EPI_ISL_1714348 |
| hCoV-19/Qatar/QA-WCMQ_FD17571358/2020 | EPI_ISL_1714349 |
| hCoV-19/Qatar/QA-WCMQ_FD17571359/2020 | EPI_ISL_1714350 |
| hCoV-19/Qatar/QA-QU_15-2-D2/2020 | EPI_ISL_1713394 |
| hCoV-19/Qatar/QA-QU_15-2-D3/2020 | EPI_ISL_1713395 |
| hCoV-19/Qatar/QA-QU_15-2-E1/2020 | EPI_ISL_1713396 |
| hCoV-19/Qatar/QA-QU_16-C10/2020 | EPI_ISL_1713440 |
| hCoV-19/Qatar/QA-QU_16-C11/2020 | EPI_ISL_1713441 |
| hCoV-19/Qatar/QA-QU_16-C12/2020 | EPI_ISL_1713442 |
| hCoV-19/Qatar/QA-QU_16-D1/2020 | EPI_ISL_1713449 |
| hCoV-19/Qatar/QA-QU_16-D2/2020 | EPI_ISL_1713450 |
| hCoV-19/Qatar/QA-QU_16-D3/2020 | EPI_ISL_1713451 |
| hCoV-19/Qatar/QA-QU_16-D4/2020 | EPI_ISL_1713452 |
| hCoV-19/Qatar/QA-QU_16-D5/2020 | EPI_ISL_1713453 |
| hCoV-19/Qatar/QA-QU_16-D6/2020 | EPI_ISL_1713454 |

| | |
|---|---|
| hCoV-19/Qatar/QA-QU_16-D7/2020 | EPI_ISL_1713455 |
| hCoV-19/Qatar/QA-QU_15-3-B9/2020 | EPI_ISL_1713404 |
| hCoV-19/Qatar/QA-QU_15-3-G3/2020 | EPI_ISL_1713410 |
| hCoV-19/Qatar/QA-QU_17-D10/2020 | EPI_ISL_1713485 |
| hCoV-19/Qatar/QA-QU_17-D9/2020 | EPI_ISL_1713489 |
| hCoV-19/Qatar/QA-QU_15-3-D9/2021 | EPI_ISL_1713407 |
| hCoV-19/Qatar/QA-QU_15-3-E8/2021 | EPI_ISL_1713408 |
| hCoV-19/Qatar/QA-QU_15-3-F7/2021 | EPI_ISL_1713409 |
| hCoV-19/Qatar/QA-QU_16-C7/2021 | EPI_ISL_1713448 |
| hCoV-19/Qatar/QA-QU_15-3-B2/2021 | EPI_ISL_1713403 |
| hCoV-19/Qatar/QA-QU_15-3-C12/2021 | EPI_ISL_1713405 |
| hCoV-19/Qatar/QA-QU_15-3-D11/2021 | EPI_ISL_1713406 |
| hCoV-19/Qatar/QA-QU_17-D12/2021 | EPI_ISL_1713486 |
| hCoV-19/Qatar/QA-QU_17-E1/2021 | EPI_ISL_1713490 |
| hCoV-19/Qatar/QA-QU_17-E2/2021 | EPI_ISL_1713494 |
| hCoV-19/Qatar/QA-QU_17-H1/2021 | EPI_ISL_1713510 |
| hCoV-19/Qatar/QA-QU_17-H4/2021 | EPI_ISL_1713512 |
| hCoV-19/Qatar/QA-QU_17-H5/2021 | EPI_ISL_1713513 |
| hCoV-19/Qatar/QA-QU_17-H6/2021 | EPI_ISL_1713514 |
| hCoV-19/Qatar/QA-QU_17-H7/2021 | EPI_ISL_1713515 |
| hCoV-19/Qatar/QA-QU_17-E4/2021 | EPI_ISL_1713495 |
| hCoV-19/Qatar/QA-QU_17-E5/2021 | EPI_ISL_1713496 |
| hCoV-19/Qatar/QA-QU_17-H2/2021 | EPI_ISL_1713511 |
| hCoV-19/Qatar/QA-QU_17-E6/2021 | EPI_ISL_1713497 |
| hCoV-19/Qatar/QA-QU_17-G4/2021 | EPI_ISL_1713508 |
| hCoV-19/Qatar/QA-QU_17-E10/2021 | EPI_ISL_1713491 |
| hCoV-19/Qatar/QA-QU_17-H8/2021 | EPI_ISL_1713516 |
| hCoV-19/Qatar/QA-QU_16-F12/2021 | EPI_ISL_1713465 |
| hCoV-19/Qatar/QA-QU_17-E11/2021 | EPI_ISL_1713492 |
| hCoV-19/Qatar/QA-QU_16-A1/2021 | EPI_ISL_1713421 |
| hCoV-19/Qatar/QA-QU_16-A10/2021 | EPI_ISL_1713422 |
| hCoV-19/Qatar/QA-QU_16-A2/2021 | EPI_ISL_1713423 |
| hCoV-19/Qatar/QA-QU_16-A3/2021 | EPI_ISL_1713424 |
| hCoV-19/Qatar/QA-QU_16-A4/2021 | EPI_ISL_1713425 |
| hCoV-19/Qatar/QA-QU_16-A5/2021 | EPI_ISL_1713426 |
| hCoV-19/Qatar/QA-QU_16-A6/2021 | EPI_ISL_1713427 |
| hCoV-19/Qatar/QA-QU_16-A7/2021 | EPI_ISL_1713428 |
| hCoV-19/Qatar/QA-QU_16-A8/2021 | EPI_ISL_1713429 |
| hCoV-19/Qatar/QA-QU_16-A9/2021 | EPI_ISL_1713430 |
| hCoV-19/Qatar/QA-QU_16-F11/2021 | EPI_ISL_1713464 |
| hCoV-19/Qatar/QA-QU_17-E12/2021 | EPI_ISL_1713493 |
| hCoV-19/Qatar/QA-QU_17-F1/2021 | EPI_ISL_1713498 |
| hCoV-19/Qatar/QA-QU_17-F5/2021 | EPI_ISL_1713501 |
| hCoV-19/Qatar/QA-QU_17-F6/2021 | EPI_ISL_1713502 |
| hCoV-19/Qatar/QA-QU_17-F10/2021 | EPI_ISL_1713499 |
| hCoV-19/Qatar/QA-QU_17-F8/2021 | EPI_ISL_1713503 |
| hCoV-19/Qatar/QA-QU_17-F9/2021 | EPI_ISL_1713504 |
| hCoV-19/Qatar/QA-QU_17-G1/2021 | EPI_ISL_1713505 |
| hCoV-19/Qatar/QA-QU_17-G12/2021 | EPI_ISL_1713507 |
| hCoV-19/Qatar/QA-QU_17-F11/2021 | EPI_ISL_1713500 |

| | |
|---|---|
| hCoV-19/Qatar/QA-QU_17-G10/2021 | EPI_ISL_1713506 |
| hCoV-19/Qatar/QA-QU_17-G9/2021 | EPI_ISL_1713509 |
| hCoV-19/Qatar/QA-QU_18-H7/2021 | EPI_ISL_1713923 |
| hCoV-19/Qatar/QA-QU_18-D10/2021 | EPI_ISL_1713891 |
| hCoV-19/Qatar/QA-QU_18-F10/2021 | EPI_ISL_1713907 |
| hCoV-19/Qatar/QA-QU_19-1-B11/2021 | EPI_ISL_1713938 |
| hCoV-19/Qatar/QA-QU_18-A5/2021 | EPI_ISL_1713867 |
| hCoV-19/Qatar/QA-QU_18-A8/2021 | EPI_ISL_1713870 |
| hCoV-19/Qatar/QA-QU_18-B6/2021 | EPI_ISL_1713877 |
| hCoV-19/Qatar/QA-QU_18-C7/2021 | EPI_ISL_1713888 |
| hCoV-19/Qatar/QA-QU_18-D7/2021 | EPI_ISL_1713896 |
| hCoV-19/Qatar/QA-QU_18-E7/2021 | EPI_ISL_1713903 |
| hCoV-19/Qatar/QA-QU_18-F5/2021 | EPI_ISL_1713910 |
| hCoV-19/Qatar/QA-QU_18-G3/2021 | EPI_ISL_1713916 |
| hCoV-19/Qatar/QA-QU_18-G8/2021 | EPI_ISL_1713918 |
| hCoV-19/Qatar/QA-QU_18-A6/2021 | EPI_ISL_1713868 |
| hCoV-19/Qatar/QA-QU_18-B8/2021 | EPI_ISL_1713879 |
| hCoV-19/Qatar/QA-QU_18-C6/2021 | EPI_ISL_1713887 |
| hCoV-19/Qatar/QA-QU_18-C8/2021 | EPI_ISL_1713889 |
| hCoV-19/Qatar/QA-QU_18-D6/2021 | EPI_ISL_1713895 |
| hCoV-19/Qatar/QA-QU_18-E3/2021 | EPI_ISL_1713899 |
| hCoV-19/Qatar/QA-QU_18-E6/2021 | EPI_ISL_1713902 |
| hCoV-19/Qatar/QA-QU_18-E8/2021 | EPI_ISL_1713904 |
| hCoV-19/Qatar/QA-QU_18-F3/2021 | EPI_ISL_1713908 |
| hCoV-19/Qatar/QA-QU_18-F8/2021 | EPI_ISL_1713912 |
| hCoV-19/Qatar/QA-QU_18-H4/2021 | EPI_ISL_1713922 |
| hCoV-19/Qatar/QA-QU_19-1-A9/2021 | EPI_ISL_1713935 |
| hCoV-19/Qatar/QA-QU_19-1-B10/2020 | EPI_ISL_1713937 |
| hCoV-19/Qatar/QA-QU_18-A4/2021 | EPI_ISL_1713866 |
| hCoV-19/Qatar/QA-QU_18-A7/2021 | EPI_ISL_1713869 |
| hCoV-19/Qatar/QA-QU_18-A9/2021 | EPI_ISL_1713871 |
| hCoV-19/Qatar/QA-QU_18-B4/2021 | EPI_ISL_1713875 |
| hCoV-19/Qatar/QA-QU_18-B5/2021 | EPI_ISL_1713876 |
| hCoV-19/Qatar/QA-QU_18-B7/2021 | EPI_ISL_1713878 |
| hCoV-19/Qatar/QA-QU_18-B9/2021 | EPI_ISL_1713880 |
| hCoV-19/Qatar/QA-QU_18-C4/2021 | EPI_ISL_1713885 |
| hCoV-19/Qatar/QA-QU_18-C5/2021 | EPI_ISL_1713886 |
| hCoV-19/Qatar/QA-QU_18-C9/2021 | EPI_ISL_1713890 |
| hCoV-19/Qatar/QA-QU_18-D5/2021 | EPI_ISL_1713894 |
| hCoV-19/Qatar/QA-QU_18-E4/2021 | EPI_ISL_1713900 |
| hCoV-19/Qatar/QA-QU_18-E5/2021 | EPI_ISL_1713901 |
| hCoV-19/Qatar/QA-QU_18-F4/2021 | EPI_ISL_1713909 |
| hCoV-19/Qatar/QA-QU_18-F7/2021 | EPI_ISL_1713911 |
| hCoV-19/Qatar/QA-QU_18-G5/2021 | EPI_ISL_1713917 |
| hCoV-19/Qatar/QA-QU_19-1-A1/2021 | EPI_ISL_1713925 |
| hCoV-19/Qatar/QA-QU_18-B1/2021 | EPI_ISL_1713872 |
| hCoV-19/Qatar/QA-QU_18-C1/2021 | EPI_ISL_1713881 |
| hCoV-19/Qatar/QA-QU_18-D9/2021 | EPI_ISL_1713897 |
| hCoV-19/Qatar/QA-QU_18-F1/2021 | EPI_ISL_1713906 |
| hCoV-19/Qatar/QA-QU_18-H2/2021 | EPI_ISL_1713921 |

| | |
|---|---|
| hCoV-19/Qatar/QA-QU_19-1-A10/2021 | EPI_ISL_1713926 |
| hCoV-19/Qatar/QA-QU_18-A2/2021 | EPI_ISL_1713865 |
| hCoV-19/Qatar/QA-QU_18-C10/2021 | EPI_ISL_1713882 |
| hCoV-19/Qatar/QA-QU_18-E9/2021 | EPI_ISL_1713905 |
| hCoV-19/Qatar/QA-QU_18-F9/2021 | EPI_ISL_1713913 |
| hCoV-19/Qatar/QA-QU_18-G10/2021 | EPI_ISL_1713914 |
| hCoV-19/Qatar/QA-QU_18-H10/2021 | EPI_ISL_1713919 |
| hCoV-19/Qatar/QA-QU_18-A10/2021 | EPI_ISL_1713863 |
| hCoV-19/Qatar/QA-QU_18-A12/2021 | EPI_ISL_1713864 |
| hCoV-19/Qatar/QA-QU_18-B11/2021 | EPI_ISL_1713873 |
| hCoV-19/Qatar/QA-QU_18-B12/2021 | EPI_ISL_1713874 |
| hCoV-19/Qatar/QA-QU_18-C11/2021 | EPI_ISL_1713883 |
| hCoV-19/Qatar/QA-QU_18-C12/2021 | EPI_ISL_1713884 |
| hCoV-19/Qatar/QA-QU_18-D11/2021 | EPI_ISL_1713892 |
| hCoV-19/Qatar/QA-QU_18-D12/2021 | EPI_ISL_1713893 |
| hCoV-19/Qatar/QA-QU_18-E12/2021 | EPI_ISL_1713898 |
| hCoV-19/Qatar/QA-QU_18-G12/2021 | EPI_ISL_1713915 |
| hCoV-19/Qatar/QA-QU_18-H11/2021 | EPI_ISL_1713920 |
| hCoV-19/Qatar/QA-QU_18-H9/2021 | EPI_ISL_1713924 |
| hCoV-19/Qatar/QA-QU_19-1-A4/2021 | EPI_ISL_1713931 |
| hCoV-19/Qatar/QA-QU_19-1-B7/2021 | EPI_ISL_1713944 |
| hCoV-19/Qatar/QA-QU_19-1-B4/2021 | EPI_ISL_1713941 |
| hCoV-19/Qatar/QA-QU_19-1-B5/2021 | EPI_ISL_1713942 |
| hCoV-19/Qatar/QA-QU_18-2-B1/2021 | EPI_ISL_1713568 |
| hCoV-19/Qatar/QA-QU_18-2-E2/2021 | EPI_ISL_1713575 |
| hCoV-19/Qatar/QA-QU_18-2-E3/2021 | EPI_ISL_1713576 |
| hCoV-19/Qatar/QA-QU_18-2-F3/2021 | EPI_ISL_1713578 |
| hCoV-19/Qatar/QA-QU_18-2-B4/2021 | EPI_ISL_1713570 |
| hCoV-19/Qatar/QA-QU_18-2-B5/2021 | EPI_ISL_1713571 |
| hCoV-19/Qatar/QA-QU_18-2-E6/2021 | EPI_ISL_1713577 |
| hCoV-19/Qatar/QA-QU_18-2-F5/2021 | EPI_ISL_1713579 |
| hCoV-19/Qatar/QA-QU_18-2-H6/2021 | EPI_ISL_1713582 |
| hCoV-19/Qatar/QA-QU_19-1-B3/2021 | EPI_ISL_1713940 |
| hCoV-19/Qatar/QA-QU_19-1-B8/2021 | EPI_ISL_1713945 |
| hCoV-19/Qatar/QA-QU_19-1-B1/2021 | EPI_ISL_1713936 |
| hCoV-19/Qatar/QA-QU_18-2-2-B1/2021 | EPI_ISL_1713567 |
| hCoV-19/Qatar/QA-QU_18-2-B7/2021 | EPI_ISL_1713572 |
| hCoV-19/Qatar/QA-QU_18-2-G11/2021 | EPI_ISL_1713580 |
| hCoV-19/Qatar/QA-QU_18-2-H9/2021 | EPI_ISL_1713583 |
| hCoV-19/Qatar/QA-QU_19-1-A5/2021 | EPI_ISL_1713932 |
| hCoV-19/Qatar/QA-QU_19-1-A7/2021 | EPI_ISL_1713934 |
| hCoV-19/Qatar/QA-QU_19-1-B9/2021 | EPI_ISL_1713946 |
| hCoV-19/Qatar/QA-QU_18-2-B11/2021 | EPI_ISL_1713569 |
| hCoV-19/Qatar/QA-QU_18-2-C11/2021 | EPI_ISL_1713573 |
| hCoV-19/Qatar/QA-QU_18-2-C12/2021 | EPI_ISL_1713574 |
| hCoV-19/Qatar/QA-QU_18-2-G9/2021 | EPI_ISL_1713581 |
| hCoV-19/Qatar/QA-QU_19-1-A2/2021 | EPI_ISL_1713929 |
| hCoV-19/Qatar/QA-QU_18-3-A11/2021 | EPI_ISL_1713585 |
| hCoV-19/Qatar/QA-QU_18-3-A12/2021 | EPI_ISL_1713586 |
| hCoV-19/Qatar/QA-QU_18-3-B12/2021 | EPI_ISL_1713592 |

| | |
|---|---|
| hCoV-19/Qatar/QA-QU_18-3-D12/2021 | EPI_ISL_1713602 |
| hCoV-19/Qatar/QA-QU_18-3-F11/2021 | EPI_ISL_1713611 |
| hCoV-19/Qatar/QA-QU_18-3-G11/2021 | EPI_ISL_1713617 |
| hCoV-19/Qatar/QA-QU_18-3-H12/2021 | EPI_ISL_1713621 |
| hCoV-19/Qatar/QA-QU_18-3-A8/2021 | EPI_ISL_1713589 |
| hCoV-19/Qatar/QA-QU_18-3-A9/2021 | EPI_ISL_1713590 |
| hCoV-19/Qatar/QA-QU_18-3-B10/2021 | EPI_ISL_1713591 |
| hCoV-19/Qatar/QA-QU_18-3-B8/2021 | EPI_ISL_1713594 |
| hCoV-19/Qatar/QA-QU_18-3-B9/2021 | EPI_ISL_1713595 |
| hCoV-19/Qatar/QA-QU_18-3-C9/2021 | EPI_ISL_1713600 |
| hCoV-19/Qatar/QA-QU_18-3-E10/2021 | EPI_ISL_1713605 |
| hCoV-19/Qatar/QA-QU_18-3-E8/2021 | EPI_ISL_1713608 |
| hCoV-19/Qatar/QA-QU_18-3-F10/2021 | EPI_ISL_1713610 |
| hCoV-19/Qatar/QA-QU_18-3-F8/2021 | EPI_ISL_1713614 |
| hCoV-19/Qatar/QA-QU_18-3-F9/2021 | EPI_ISL_1713615 |
| hCoV-19/Qatar/QA-QU_18-3-G8/2021 | EPI_ISL_1713619 |
| hCoV-19/Qatar/QA-QU_18-3-G9/2021 | EPI_ISL_1713620 |
| hCoV-19/Qatar/QA-QU_18-3-A6/2021 | EPI_ISL_1713588 |
| hCoV-19/Qatar/QA-QU_18-3-C5/2021 | EPI_ISL_1713598 |
| hCoV-19/Qatar/QA-QU_18-3-C7/2021 | EPI_ISL_1713599 |
| hCoV-19/Qatar/QA-QU_18-3-D6/2021 | EPI_ISL_1713604 |
| hCoV-19/Qatar/QA-QU_18-3-E5/2021 | EPI_ISL_1713606 |
| hCoV-19/Qatar/QA-QU_18-3-E7/2021 | EPI_ISL_1713607 |
| hCoV-19/Qatar/QA-QU_18-3-F5/2021 | EPI_ISL_1713612 |
| hCoV-19/Qatar/QA-QU_18-3-F7/2021 | EPI_ISL_1713613 |
| hCoV-19/Qatar/QA-QU_19-1-A11/2021 | EPI_ISL_1713927 |
| hCoV-19/Qatar/QA-QU_19-1-A6/2021 | EPI_ISL_1713933 |
| hCoV-19/Qatar/QA-QU_18-3-A3/2021 | EPI_ISL_1713587 |
| hCoV-19/Qatar/QA-QU_18-3-B3/2021 | EPI_ISL_1713593 |
| hCoV-19/Qatar/QA-QU_18-3-C3/2021 | EPI_ISL_1713597 |
| hCoV-19/Qatar/QA-QU_18-3-D3/2021 | EPI_ISL_1713603 |
| hCoV-19/Qatar/QA-QU_18-3-G2/2021 | EPI_ISL_1713618 |
| hCoV-19/Qatar/QA-QU_18-3-H2/2021 | EPI_ISL_1713622 |
| hCoV-19/Qatar/QA-QU_18-3-H3/2021 | EPI_ISL_1713623 |
| hCoV-19/Qatar/QA-QU_18-3-H4/2021 | EPI_ISL_1713624 |
| hCoV-19/Qatar/QA-QU_18-3-C2/2021 | EPI_ISL_1713596 |
| hCoV-19/Qatar/QA-QU_18-3-F1/2021 | EPI_ISL_1713609 |
| hCoV-19/Qatar/QA-QU_18-3-G1/2021 | EPI_ISL_1713616 |
| hCoV-19/Qatar/QA-QU_18-4-A11/2021 | EPI_ISL_1713626 |
| hCoV-19/Qatar/QA-QU_18-4-D11/2021 | EPI_ISL_1713642 |
| hCoV-19/Qatar/QA-QU_18-4-E12/2021 | EPI_ISL_1713645 |
| hCoV-19/Qatar/QA-QU_18-4-F12/2021 | EPI_ISL_1713651 |
| hCoV-19/Qatar/QA-QU_18-4-H10/2021 | EPI_ISL_1713660 |
| hCoV-19/Qatar/QA-QU_18-4-H12/2021 | EPI_ISL_1713661 |
| hCoV-19/Qatar/QA-QU_18-3-A1/2021 | EPI_ISL_1713584 |
| hCoV-19/Qatar/QA-QU_18-3-D1/2021 | EPI_ISL_1713601 |
| hCoV-19/Qatar/QA-QU_18-4-A9/2021 | EPI_ISL_1713629 |
| hCoV-19/Qatar/QA-QU_18-4-B10/2021 | EPI_ISL_1713630 |
| hCoV-19/Qatar/QA-QU_18-4-C12/2021 | EPI_ISL_1713635 |
| hCoV-19/Qatar/QA-QU_18-4-C9/2021 | EPI_ISL_1713640 |

| | |
|---|---|
| hCoV-19/Qatar/QA-QU_18-4-D10/2021 | EPI_ISL_1713641 |
| hCoV-19/Qatar/QA-QU_18-4-E8/2021 | EPI_ISL_1713649 |
| hCoV-19/Qatar/QA-QU_18-4-E9/2021 | EPI_ISL_1713650 |
| hCoV-19/Qatar/QA-QU_18-4-G9/2021 | EPI_ISL_1713659 |
| hCoV-19/Qatar/QA-QU_18-4-H8/2021 | EPI_ISL_1713665 |
| hCoV-19/Qatar/QA-QU_18-4-A6/2021 | EPI_ISL_1713628 |
| hCoV-19/Qatar/QA-QU_18-4-B8/2021 | EPI_ISL_1713633 |
| hCoV-19/Qatar/QA-QU_18-4-C6/2021 | EPI_ISL_1713638 |
| hCoV-19/Qatar/QA-QU_18-4-C8/2021 | EPI_ISL_1713639 |
| hCoV-19/Qatar/QA-QU_18-4-E6/2021 | EPI_ISL_1713648 |
| hCoV-19/Qatar/QA-QU_18-4-F5/2021 | EPI_ISL_1713653 |
| hCoV-19/Qatar/QA-QU_18-4-F7/2021 | EPI_ISL_1713654 |
| hCoV-19/Qatar/QA-QU_18-4-G6/2021 | EPI_ISL_1713657 |
| hCoV-19/Qatar/QA-QU_18-4-G7/2021 | EPI_ISL_1713658 |
| hCoV-19/Qatar/QA-QU_18-4-H5/2021 | EPI_ISL_1713664 |
| hCoV-19/Qatar/QA-QU_18-4-A4/2021 | EPI_ISL_1713627 |
| hCoV-19/Qatar/QA-QU_18-4-B2/2021 | EPI_ISL_1713631 |
| hCoV-19/Qatar/QA-QU_18-4-B5/2021 | EPI_ISL_1713632 |
| hCoV-19/Qatar/QA-QU_18-4-C3/2021 | EPI_ISL_1713636 |
| hCoV-19/Qatar/QA-QU_18-4-C4/2021 | EPI_ISL_1713637 |
| hCoV-19/Qatar/QA-QU_18-4-D2/2021 | EPI_ISL_1713643 |
| hCoV-19/Qatar/QA-QU_18-4-E3/2021 | EPI_ISL_1713646 |
| hCoV-19/Qatar/QA-QU_18-4-E4/2021 | EPI_ISL_1713647 |
| hCoV-19/Qatar/QA-QU_18-4-F2/2021 | EPI_ISL_1713652 |
| hCoV-19/Qatar/QA-QU_18-4-G2/2021 | EPI_ISL_1713655 |
| hCoV-19/Qatar/QA-QU_18-4-G4/2021 | EPI_ISL_1713656 |
| hCoV-19/Qatar/QA-QU_18-4-H2/2021 | EPI_ISL_1713662 |
| hCoV-19/Qatar/QA-QU_18-4-H3/2021 | EPI_ISL_1713663 |
| hCoV-19/Qatar/QA-QU_18-4-A1/2021 | EPI_ISL_1713625 |
| hCoV-19/Qatar/QA-QU_18-4-C1/2021 | EPI_ISL_1713634 |
| hCoV-19/Qatar/QA-QU_18-4-E1/2021 | EPI_ISL_1713644 |
| hCoV-19/Qatar/QA-QU_18-5-2-A2/2021 | EPI_ISL_1713691 |
| hCoV-19/Qatar/QA-QU_18-5-2-A6/2021 | EPI_ISL_1713693 |
| hCoV-19/Qatar/QA-QU_18-5-2-A9/2021 | EPI_ISL_1713694 |
| hCoV-19/Qatar/QA-QU_18-5-2-B9/2021 | EPI_ISL_1713696 |
| hCoV-19/Qatar/QA-QU_18-5-2-C2/2021 | EPI_ISL_1713698 |
| hCoV-19/Qatar/QA-QU_18-5-2-H6/2021 | EPI_ISL_1713712 |
| hCoV-19/Qatar/QA-QU_18-5-2-C1/2021 | EPI_ISL_1713697 |
| hCoV-19/Qatar/QA-QU_18-5-2-E5/2021 | EPI_ISL_1713704 |
| hCoV-19/Qatar/QA-QU_18-5-2-E8/2021 | EPI_ISL_1713705 |
| hCoV-19/Qatar/QA-QU_18-5-2-F5/2021 | EPI_ISL_1713707 |
| hCoV-19/Qatar/QA-QU_18-5-2-F6/2021 | EPI_ISL_1713708 |
| hCoV-19/Qatar/QA-QU_18-5-2-A10/2021 | EPI_ISL_1713689 |
| hCoV-19/Qatar/QA-QU_18-5-2-A12/2021 | EPI_ISL_1713690 |
| hCoV-19/Qatar/QA-QU_18-5-2-A4/2021 | EPI_ISL_1713692 |
| hCoV-19/Qatar/QA-QU_18-5-2-D6/2021 | EPI_ISL_1713701 |
| hCoV-19/Qatar/QA-QU_18-5-2-H2/2021 | EPI_ISL_1713710 |
| hCoV-19/Qatar/QA-QU_18-5-2-H5/2021 | EPI_ISL_1713711 |
| hCoV-19/Qatar/QA-QU_18-5-2-B4/2021 | EPI_ISL_1713695 |
| hCoV-19/Qatar/QA-QU_18-5-2-D2/2021 | EPI_ISL_1713699 |

| | |
|---|---|
| hCoV-19/Qatar/QA-QU_18-5-2-D3/2021 | EPI_ISL_1713700 |
| hCoV-19/Qatar/QA-QU_18-5-2-E2/2021 | EPI_ISL_1713702 |
| hCoV-19/Qatar/QA-QU_18-5-2-E4/2021 | EPI_ISL_1713703 |
| hCoV-19/Qatar/QA-QU_18-5-2-G4/2021 | EPI_ISL_1713709 |
| hCoV-19/Qatar/QA-QU_18-5-3-A5/2021 | EPI_ISL_1713714 |
| hCoV-19/Qatar/QA-QU_18-5-3-A8/2021 | EPI_ISL_1713717 |
| hCoV-19/Qatar/QA-QU_18-5-3-B7/2021 | EPI_ISL_1713719 |
| hCoV-19/Qatar/QA-QU_18-5-3-B9/2021 | EPI_ISL_1713720 |
| hCoV-19/Qatar/QA-QU_18-5-3-C10/2021 | EPI_ISL_1713721 |
| hCoV-19/Qatar/QA-QU_18-5-3-C8/2021 | EPI_ISL_1713722 |
| hCoV-19/Qatar/QA-QU_18-5-3-A6/2021 | EPI_ISL_1713715 |
| hCoV-19/Qatar/QA-QU_18-5-3-A7/2021 | EPI_ISL_1713716 |
| hCoV-19/Qatar/QA-QU_18-5-3-D4/2021 | EPI_ISL_1713724 |
| hCoV-19/Qatar/QA-QU_18-5-3-E1/2021 | EPI_ISL_1713725 |
| hCoV-19/Qatar/QA-QU_18-5-3-E10/2021 | EPI_ISL_1713726 |
| hCoV-19/Qatar/QA-QU_18-5-3-G7/2021 | EPI_ISL_1713733 |
| hCoV-19/Qatar/QA-QU_18-5-3-E4/2021 | EPI_ISL_1713727 |
| hCoV-19/Qatar/QA-QU_18-5-3-E9/2021 | EPI_ISL_1713729 |
| hCoV-19/Qatar/QA-QU_18-5-3-G1/2021 | EPI_ISL_1713730 |
| hCoV-19/Qatar/QA-QU_18-5-3-G3/2021 | EPI_ISL_1713732 |
| hCoV-19/Qatar/QA-QU_18-5-2-F4/2021 | EPI_ISL_1713706 |
| hCoV-19/Qatar/QA-QU_18-5-3-A12/2021 | EPI_ISL_1713713 |
| hCoV-19/Qatar/QA-QU_18-5-3-B1/2021 | EPI_ISL_1713718 |
| hCoV-19/Qatar/QA-QU_18-5-3-D11/2021 | EPI_ISL_1713723 |
| hCoV-19/Qatar/QA-QU_18-5-3-E6/2021 | EPI_ISL_1713728 |
| hCoV-19/Qatar/QA-QU_18-5-3-G11/2021 | EPI_ISL_1713731 |
| hCoV-19/Qatar/QA-QU_18-5-1-C10/2021 | EPI_ISL_1713668 |
| hCoV-19/Qatar/QA-QU_18-5-1-D10/2021 | EPI_ISL_1713673 |
| hCoV-19/Qatar/QA-QU_18-5-1-D12/2021 | EPI_ISL_1713674 |
| hCoV-19/Qatar/QA-QU_18-5-1-E11/2021 | EPI_ISL_1713676 |
| hCoV-19/Qatar/QA-QU_18-5-1-E12/2021 | EPI_ISL_1713677 |
| hCoV-19/Qatar/QA-QU_18-5-1-G12/2021 | EPI_ISL_1713684 |
| hCoV-19/Qatar/QA-QU_18-5-1-E9/2021 | EPI_ISL_1713680 |
| hCoV-19/Qatar/QA-QU_18-5-1-F7/2021 | EPI_ISL_1713683 |
| hCoV-19/Qatar/QA-QU_18-5-1-G9/2021 | EPI_ISL_1713685 |
| hCoV-19/Qatar/QA-QU_18-5-1-H6/2021 | EPI_ISL_1713687 |
| hCoV-19/Qatar/QA-QU_18-5-1-H9/2021 | EPI_ISL_1713688 |
| hCoV-19/Qatar/QA-QU_18-5-1-C2/2021 | EPI_ISL_1713669 |
| hCoV-19/Qatar/QA-QU_18-5-1-C3/2021 | EPI_ISL_1713670 |
| hCoV-19/Qatar/QA-QU_18-5-1-C6/2021 | EPI_ISL_1713671 |
| hCoV-19/Qatar/QA-QU_18-5-1-D2/2021 | EPI_ISL_1713675 |
| hCoV-19/Qatar/QA-QU_18-5-1-E5/2021 | EPI_ISL_1713679 |
| hCoV-19/Qatar/QA-QU_18-5-1-F6/2021 | EPI_ISL_1713682 |
| hCoV-19/Qatar/QA-QU_18-5-1-A4/2021 | EPI_ISL_1713666 |
| hCoV-19/Qatar/QA-QU_18-5-1-B2/2021 | EPI_ISL_1713667 |
| hCoV-19/Qatar/QA-QU_18-5-1-D1/2021 | EPI_ISL_1713672 |
| hCoV-19/Qatar/QA-QU_18-5-1-E4/2021 | EPI_ISL_1713678 |
| hCoV-19/Qatar/QA-QU_18-5-1-F1/2021 | EPI_ISL_1713681 |
| hCoV-19/Qatar/QA-QU_18-5-1-H1/2021 | EPI_ISL_1713686 |
| hCoV-19/Qatar/QA-QU_18-6-C3/2021 | EPI_ISL_1713735 |

| | |
|---|---|
| hCoV-19/Qatar/QA-QU_18-6-C4/2021 | EPI_ISL_1713736 |
| hCoV-19/Qatar/QA-QU_18-6-F2/2021 | EPI_ISL_1713744 |
| hCoV-19/Qatar/QA-QU_18-6-F3/2021 | EPI_ISL_1713745 |
| hCoV-19/Qatar/QA-QU_18-6-F4/2021 | EPI_ISL_1713746 |
| hCoV-19/Qatar/QA-QU_18-6-H4/2021 | EPI_ISL_1713753 |
| hCoV-19/Qatar/QA-QU_18-6-C7/2021 | EPI_ISL_1713737 |
| hCoV-19/Qatar/QA-QU_18-6-D7/2021 | EPI_ISL_1713740 |
| hCoV-19/Qatar/QA-QU_18-6-G4/2021 | EPI_ISL_1713749 |
| hCoV-19/Qatar/QA-QU_18-6-G5/2021 | EPI_ISL_1713750 |
| hCoV-19/Qatar/QA-QU_18-6-H6/2021 | EPI_ISL_1713754 |
| hCoV-19/Qatar/QA-QU_18-6-C9/2021 | EPI_ISL_1713738 |
| hCoV-19/Qatar/QA-QU_18-6-D1/2021 | EPI_ISL_1713739 |
| hCoV-19/Qatar/QA-QU_18-6-D9/2021 | EPI_ISL_1713741 |
| hCoV-19/Qatar/QA-QU_18-6-F8/2021 | EPI_ISL_1713747 |
| hCoV-19/Qatar/QA-QU_18-6-G9/2021 | EPI_ISL_1713751 |
| hCoV-19/Qatar/QA-QU_18-6-H9/2021 | EPI_ISL_1713755 |
| hCoV-19/Qatar/QA-QU_18-6-B10/2021 | EPI_ISL_1713734 |
| hCoV-19/Qatar/QA-QU_18-6-E12/2021 | EPI_ISL_1713742 |
| hCoV-19/Qatar/QA-QU_18-6-F12/2021 | EPI_ISL_1713743 |
| hCoV-19/Qatar/QA-QU_18-6-G12/2021 | EPI_ISL_1713748 |
| hCoV-19/Qatar/QA-QU_18-6-H11/2021 | EPI_ISL_1713752 |
| hCoV-19/Qatar/QA-QU_18-7-H12/2021 | EPI_ISL_1713775 |
| hCoV-19/Qatar/QA-QU_18-9-A1/2021 | EPI_ISL_1713820 |
| hCoV-19/Qatar/QA-QU_18-7-C9/2021 | EPI_ISL_1713763 |
| hCoV-19/Qatar/QA-QU_18-7-D9/2021 | EPI_ISL_1713764 |
| hCoV-19/Qatar/QA-QU_18-7-F11/2021 | EPI_ISL_1713769 |
| hCoV-19/Qatar/QA-QU_18-7-G11/2021 | EPI_ISL_1713773 |
| hCoV-19/Qatar/QA-QU_18-7-H10/2021 | EPI_ISL_1713774 |
| hCoV-19/Qatar/QA-QU_18-7-A8/2021 | EPI_ISL_1713758 |
| hCoV-19/Qatar/QA-QU_18-7-B6/2021 | EPI_ISL_1713760 |
| hCoV-19/Qatar/QA-QU_18-7-B8/2021 | EPI_ISL_1713761 |
| hCoV-19/Qatar/QA-QU_18-7-E8/2021 | EPI_ISL_1713767 |
| hCoV-19/Qatar/QA-QU_18-7-F6/2021 | EPI_ISL_1713772 |
| hCoV-19/Qatar/QA-QU_18-7-H8/2021 | EPI_ISL_1713777 |
| hCoV-19/Qatar/QA-QU_18-7-B2/2021 | EPI_ISL_1713759 |
| hCoV-19/Qatar/QA-QU_18-7-E5/2021 | EPI_ISL_1713766 |
| hCoV-19/Qatar/QA-QU_18-7-F1/2021 | EPI_ISL_1713768 |
| hCoV-19/Qatar/QA-QU_18-7-F2/2021 | EPI_ISL_1713770 |
| hCoV-19/Qatar/QA-QU_18-7-H2/2021 | EPI_ISL_1713776 |
| hCoV-19/Qatar/QA-QU_18-7-A2/2021 | EPI_ISL_1713756 |
| hCoV-19/Qatar/QA-QU_18-7-A4/2021 | EPI_ISL_1713757 |
| hCoV-19/Qatar/QA-QU_18-7-C4/2021 | EPI_ISL_1713762 |
| hCoV-19/Qatar/QA-QU_18-7-E3/2021 | EPI_ISL_1713765 |
| hCoV-19/Qatar/QA-QU_18-7-F3/2021 | EPI_ISL_1713771 |
| hCoV-19/Qatar/QA-QU_18-8-B2/2021 | EPI_ISL_1713784 |
| hCoV-19/Qatar/QA-QU_18-8-E2/2021 | EPI_ISL_1713803 |
| hCoV-19/Qatar/QA-QU_18-8-G1/2021 | EPI_ISL_1713811 |
| hCoV-19/Qatar/QA-QU_18-8-H1/2021 | EPI_ISL_1713816 |
| hCoV-19/Qatar/QA-QU_18-8-A5/2021 | EPI_ISL_1713780 |
| hCoV-19/Qatar/QA-QU_18-8-A6/2021 | EPI_ISL_1713781 |

| | |
|---|---|
| hCoV-19/Qatar/QA-QU_18-8-B5/2021 | EPI_ISL_1713785 |
| hCoV-19/Qatar/QA-QU_18-8-C4/2021 | EPI_ISL_1713790 |
| hCoV-19/Qatar/QA-QU_18-8-D3/2021 | EPI_ISL_1713797 |
| hCoV-19/Qatar/QA-QU_18-8-E1/2021 | EPI_ISL_1713801 |
| hCoV-19/Qatar/QA-QU_18-8-E3/2021 | EPI_ISL_1713804 |
| hCoV-19/Qatar/QA-QU_18-8-F3/2021 | EPI_ISL_1713807 |
| hCoV-19/Qatar/QA-QU_18-8-F5/2021 | EPI_ISL_1713808 |
| hCoV-19/Qatar/QA-QU_18-8-A11/2021 | EPI_ISL_1713779 |
| hCoV-19/Qatar/QA-QU_18-8-A7/2021 | EPI_ISL_1713782 |
| hCoV-19/Qatar/QA-QU_18-8-B6/2021 | EPI_ISL_1713786 |
| hCoV-19/Qatar/QA-QU_18-8-C6/2021 | EPI_ISL_1713791 |
| hCoV-19/Qatar/QA-QU_18-8-C8/2021 | EPI_ISL_1713792 |
| hCoV-19/Qatar/QA-QU_18-8-D7/2021 | EPI_ISL_1713798 |
| hCoV-19/Qatar/QA-QU_18-8-D8/2021 | EPI_ISL_1713799 |
| hCoV-19/Qatar/QA-QU_18-8-E6/2021 | EPI_ISL_1713805 |
| hCoV-19/Qatar/QA-QU_18-8-F6/2021 | EPI_ISL_1713809 |
| hCoV-19/Qatar/QA-QU_18-8-F7/2021 | EPI_ISL_1713810 |
| hCoV-19/Qatar/QA-QU_18-8-G8/2021 | EPI_ISL_1713814 |
| hCoV-19/Qatar/QA-QU_18-8-A10/2021 | EPI_ISL_1713778 |
| hCoV-19/Qatar/QA-QU_18-8-B1/2021 | EPI_ISL_1713783 |
| hCoV-19/Qatar/QA-QU_18-8-B9/2021 | EPI_ISL_1713787 |
| hCoV-19/Qatar/QA-QU_18-8-C12/2021 | EPI_ISL_1713789 |
| hCoV-19/Qatar/QA-QU_18-8-D1/2021 | EPI_ISL_1713793 |
| hCoV-19/Qatar/QA-QU_18-8-D11/2021 | EPI_ISL_1713795 |
| hCoV-19/Qatar/QA-QU_18-8-D12/2021 | EPI_ISL_1713796 |
| hCoV-19/Qatar/QA-QU_18-8-D9/2021 | EPI_ISL_1713800 |
| hCoV-19/Qatar/QA-QU_18-8-E8/2021 | EPI_ISL_1713806 |
| hCoV-19/Qatar/QA-QU_18-8-G9/2021 | EPI_ISL_1713815 |
| hCoV-19/Qatar/QA-QU_18-8-H6/2021 | EPI_ISL_1713818 |
| hCoV-19/Qatar/QA-QU_18-8-C10/2021 | EPI_ISL_1713788 |
| hCoV-19/Qatar/QA-QU_18-8-D10/2021 | EPI_ISL_1713794 |
| hCoV-19/Qatar/QA-QU_18-8-E11/2021 | EPI_ISL_1713802 |
| hCoV-19/Qatar/QA-QU_18-8-G11/2021 | EPI_ISL_1713812 |
| hCoV-19/Qatar/QA-QU_18-8-G12/2021 | EPI_ISL_1713813 |
| hCoV-19/Qatar/QA-QU_18-8-H10/2021 | EPI_ISL_1713817 |
| hCoV-19/Qatar/QA-QU_18-8-H9/2021 | EPI_ISL_1713819 |
| hCoV-19/Qatar/QA-QU_18-9-A12/2021 | EPI_ISL_1713823 |
| hCoV-19/Qatar/QA-QU_18-9-D11/2021 | EPI_ISL_1713836 |
| hCoV-19/Qatar/QA-QU_18-9-D12/2021 | EPI_ISL_1713837 |
| hCoV-19/Qatar/QA-QU_18-9-E12/2021 | EPI_ISL_1713843 |
| hCoV-19/Qatar/QA-QU_18-9-F11/2021 | EPI_ISL_1713851 |
| hCoV-19/Qatar/QA-QU_18-9-G11/2021 | EPI_ISL_1713857 |
| hCoV-19/Qatar/QA-QU_18-9-A11/2021 | EPI_ISL_1713822 |
| hCoV-19/Qatar/QA-QU_18-9-B2/2021 | EPI_ISL_1713826 |
| hCoV-19/Qatar/QA-QU_18-9-D1/2021 | EPI_ISL_1713834 |
| hCoV-19/Qatar/QA-QU_18-9-D10/2021 | EPI_ISL_1713835 |
| hCoV-19/Qatar/QA-QU_18-9-D9/2021 | EPI_ISL_1713840 |
| hCoV-19/Qatar/QA-QU_18-9-E1/2021 | EPI_ISL_1713841 |
| hCoV-19/Qatar/QA-QU_18-9-E10/2021 | EPI_ISL_1713842 |
| hCoV-19/Qatar/QA-QU_18-9-F1/2021 | EPI_ISL_1713849 |

| | |
|---|---|
| hCoV-19/Qatar/QA-QU_18-9-F10/2021 | EPI_ISL_1713850 |
| hCoV-19/Qatar/QA-QU_18-9-F9/2021 | EPI_ISL_1713854 |
| hCoV-19/Qatar/QA-QU_18-9-G1/2021 | EPI_ISL_1713855 |
| hCoV-19/Qatar/QA-QU_18-9-G10/2021 | EPI_ISL_1713856 |
| hCoV-19/Qatar/QA-QU_18-9-H10/2021 | EPI_ISL_1713859 |
| hCoV-19/Qatar/QA-QU_18-9-H9/2021 | EPI_ISL_1713862 |
| hCoV-19/Qatar/QA-QU_18-9-A10/2021 | EPI_ISL_1713821 |
| hCoV-19/Qatar/QA-QU_18-9-A7/2021 | EPI_ISL_1713825 |
| hCoV-19/Qatar/QA-QU_18-9-B3/2021 | EPI_ISL_1713827 |
| hCoV-19/Qatar/QA-QU_18-9-B7/2021 | EPI_ISL_1713830 |
| hCoV-19/Qatar/QA-QU_18-9-B9/2021 | EPI_ISL_1713831 |
| hCoV-19/Qatar/QA-QU_18-9-C6/2021 | EPI_ISL_1713832 |
| hCoV-19/Qatar/QA-QU_18-9-C9/2021 | EPI_ISL_1713833 |
| hCoV-19/Qatar/QA-QU_18-9-D6/2021 | EPI_ISL_1713839 |
| hCoV-19/Qatar/QA-QU_18-9-E3/2021 | EPI_ISL_1713845 |
| hCoV-19/Qatar/QA-QU_18-9-E4/2021 | EPI_ISL_1713846 |
| hCoV-19/Qatar/QA-QU_18-9-H8/2021 | EPI_ISL_1713861 |
| hCoV-19/Qatar/QA-QU_18-9-B5/2021 | EPI_ISL_1713829 |
| hCoV-19/Qatar/QA-QU_18-9-D5/2021 | EPI_ISL_1713838 |
| hCoV-19/Qatar/QA-QU_18-9-E2/2021 | EPI_ISL_1713844 |
| hCoV-19/Qatar/QA-QU_18-9-E5/2021 | EPI_ISL_1713847 |
| hCoV-19/Qatar/QA-QU_18-9-E7/2021 | EPI_ISL_1713848 |
| hCoV-19/Qatar/QA-QU_18-9-F2/2021 | EPI_ISL_1713852 |
| hCoV-19/Qatar/QA-QU_18-9-F6/2021 | EPI_ISL_1713853 |
| hCoV-19/Qatar/QA-QU_18-9-G2/2021 | EPI_ISL_1713858 |
| hCoV-19/Qatar/QA-QU_18-9-H6/2021 | EPI_ISL_1713860 |
| hCoV-19/Qatar/QA-QU_18-10-A11/2021 | EPI_ISL_1713520 |
| hCoV-19/Qatar/QA-QU_18-10-A6/2021 | EPI_ISL_1713525 |
| hCoV-19/Qatar/QA-QU_18-10-B3/2021 | EPI_ISL_1713533 |
| hCoV-19/Qatar/QA-QU_18-9-A6/2021 | EPI_ISL_1713824 |
| hCoV-19/Qatar/QA-QU_18-10-B1/2021 | EPI_ISL_1713529 |
| hCoV-19/Qatar/QA-QU_18-10-B2/2021 | EPI_ISL_1713532 |
| hCoV-19/Qatar/QA-QU_18-10-C1/2021 | EPI_ISL_1713539 |
| hCoV-19/Qatar/QA-QU_18-10-C2/2021 | EPI_ISL_1713542 |
| hCoV-19/Qatar/QA-QU_18-10-C3/2021 | EPI_ISL_1713543 |
| hCoV-19/Qatar/QA-QU_18-10-C4/2021 | EPI_ISL_1713544 |
| hCoV-19/Qatar/QA-QU_18-10-C8/2021 | EPI_ISL_1713548 |
| hCoV-19/Qatar/QA-QU_18-9-B4/2021 | EPI_ISL_1713828 |
| hCoV-19/Qatar/QA-QU_18-10-A8/2021 | EPI_ISL_1713527 |
| hCoV-19/Qatar/QA-QU_18-10-A9/2021 | EPI_ISL_1713528 |
| hCoV-19/Qatar/QA-QU_18-10-C7/2021 | EPI_ISL_1713547 |
| hCoV-19/Qatar/QA-QU_18-10-A10/2021 | EPI_ISL_1713519 |
| hCoV-19/Qatar/QA-QU_18-10-A12/2021 | EPI_ISL_1713521 |
| hCoV-19/Qatar/QA-QU_18-10-A7/2021 | EPI_ISL_1713526 |
| hCoV-19/Qatar/QA-QU_18-10-B12/2020 | EPI_ISL_1713531 |
| hCoV-19/Qatar/QA-QU_18-10-C6/2021 | EPI_ISL_1713546 |
| hCoV-19/Qatar/QA-QU_18-10-C9/2021 | EPI_ISL_1713549 |
| hCoV-19/Qatar/QA-QU_18-10-A5/2021 | EPI_ISL_1713524 |
| hCoV-19/Qatar/QA-QU_18-10-B9/2021 | EPI_ISL_1713538 |
| hCoV-19/Qatar/QA-QU_18-10-B5/2021 | EPI_ISL_1713535 |

| | |
|---|---|
| hCoV-19/Qatar/QA-QU_18-10-B6/2021 | EPI_ISL_1713536 |
| hCoV-19/Qatar/QA-QU_18-10-B7/2021 | EPI_ISL_1713537 |
| hCoV-19/Qatar/QA-QU_18-10-C12/2021 | EPI_ISL_1713541 |
| hCoV-19/Qatar/QA-QU_18-10-C5/2021 | EPI_ISL_1713545 |
| hCoV-19/Qatar/QA-QU_18-10-B10/2021 | EPI_ISL_1713530 |
| hCoV-19/Qatar/QA-QU_18-10-B4/2021 | EPI_ISL_1713534 |
| hCoV-19/Qatar/QA-QU_18-10-D11/2021 | EPI_ISL_1713552 |
| hCoV-19/Qatar/QA-QU_18-10-D4/2021 | EPI_ISL_1713555 |
| hCoV-19/Qatar/QA-QU_18-10-D7/2021 | EPI_ISL_1713557 |
| hCoV-19/Qatar/QA-QU_18-10-D8/2021 | EPI_ISL_1713558 |
| hCoV-19/Qatar/QA-QU_18-10-D9/2021 | EPI_ISL_1713559 |
| hCoV-19/Qatar/QA-QU_18-10-D2/2021 | EPI_ISL_1713553 |
| hCoV-19/Qatar/QA-QU_18-10-D3/2021 | EPI_ISL_1713554 |
| hCoV-19/Qatar/QA-QU_18-10-E8/2021 | EPI_ISL_1713563 |
| hCoV-19/Qatar/QA-QU_18-10-F1/2021 | EPI_ISL_1713564 |
| hCoV-19/Qatar/QA-QU_18-10-F11/2021 | EPI_ISL_1713566 |
| hCoV-19/Qatar/QA-QU_16-2-A5/2020 | EPI_ISL_1713420 |
| hCoV-19/Qatar/QA-QU_16-B11/2020 | EPI_ISL_1713431 |
| hCoV-19/Qatar/QA-QU_16-B12/2020 | EPI_ISL_1713432 |
| hCoV-19/Qatar/QA-QU_16-B4/2020 | EPI_ISL_1713433 |
| hCoV-19/Qatar/QA-QU_16-B5/2020 | EPI_ISL_1713434 |
| hCoV-19/Qatar/QA-QU_16-B6/2020 | EPI_ISL_1713435 |
| hCoV-19/Qatar/QA-QU_16-B7/2020 | EPI_ISL_1713436 |
| hCoV-19/Qatar/QA-QU_16-B8/2020 | EPI_ISL_1713437 |
| hCoV-19/Qatar/QA-QU_16-B9/2020 | EPI_ISL_1713438 |
| hCoV-19/Qatar/QA-QU_16-C1/2020 | EPI_ISL_1713439 |
| hCoV-19/Qatar/QA-QU_16-C2/2020 | EPI_ISL_1713443 |
| hCoV-19/Qatar/QA-QU_16-C3/2020 | EPI_ISL_1713444 |
| hCoV-19/Qatar/QA-QU_16-C4/2020 | EPI_ISL_1713445 |
| hCoV-19/Qatar/QA-QU_16-C5/2020 | EPI_ISL_1713446 |
| hCoV-19/Qatar/QA-QU_16-C6/2020 | EPI_ISL_1713447 |
| hCoV-19/Qatar/QA-QU_19-1-A3/2020 | EPI_ISL_1713930 |
| hCoV-19/Qatar/QA-QU_19-1-B2/2020 | EPI_ISL_1713939 |
| hCoV-19/Qatar/QA-QU_19-1-B6/2020 | EPI_ISL_1713943 |
| hCoV-19/Qatar/QA-QU_18-10-A2/2020 | EPI_ISL_1713522 |
| hCoV-19/Qatar/QA-QU_18-10-A4/2020 | EPI_ISL_1713523 |
| hCoV-19/Qatar/QA-QU_18-10-C10/2020 | EPI_ISL_1713540 |
| hCoV-19/Qatar/QA-QU_18-10-D1/2020 | EPI_ISL_1713550 |
| hCoV-19/Qatar/QA-QU_18-10-D5/2020 | EPI_ISL_1713556 |
| hCoV-19/Qatar/QA-QU_16-2-A2/2020 | EPI_ISL_1713418 |
| hCoV-19/Qatar/QA-QU_16-2-A4/2021 | EPI_ISL_1713419 |
| hCoV-19/Qatar/QA-QU_19-1-A12/2020 | EPI_ISL_1713928 |
| hCoV-19/Qatar/QA-QU_18-10-A1/2020 | EPI_ISL_1713518 |
| hCoV-19/Qatar/QA-QU_18-10-D10/2020 | EPI_ISL_1713551 |
| hCoV-19/Qatar/QA-QU_18-10-E3/2021 | EPI_ISL_1713561 |
| hCoV-19/Qatar/QA-QU_18-10-F10/2021 | EPI_ISL_1713565 |
| hCoV-19/Qatar/QA-QU_18-10-E12/2021 | EPI_ISL_1713560 |
| hCoV-19/Qatar/QA-QU_18-10-E4/2021 | EPI_ISL_1713562 |
